# Continuous Diagnostic Indices from BanffNET Automated Lesion Scores for Kidney Transplant Pathology: Development and Evaluation of a Diagnostic Prediction Model

**DOI:** 10.64898/2026.09.11.26362821

**Authors:** Arthur Vranken, Giorgio Buzzanca, Thibaut Vaulet, Karolien Wellekens, Priyanka Koshy, Danny van der Helm, Soufian Meziyerh, Jan-Hendrik Venhuizen, Saskia Haitjema, Arjan D. van Zuilen, Azam S. Nurmohamed, Frederike J. Bemelman, Tri Q. Nguyen, LUMC Global Kidney Biopsy Reader Group, Sandrine Florquin, Aiko P.J. de Vries, Jesper Kers, Geert Verbeke, Maarten Naesens

## Abstract

**Background:** Histopathological diagnoses in kidney transplant biopsies, as described by the Banff classification, suffer from substantial inter-observer variability and misinterpretation, making them prone to diagnostic errors. Continuous indices based on BanffNET automated lesion scores are proposed to quantify the continuous phenotypic spectrum of kidney transplant biopsies, complementary to pathologist assessment of biopsies.

**Methods:** The BanffNET model was used to obtain continuous lesion scoring for a training cohort of 2544 biopsies, a large validation cohort of 3863 biopsies, and a multi-reader cohort of 36 biopsies scored by 67 pathologists. A penalized regression approach was used to condense 17 automated lesion scores into four indices, representing the most important dimensions of graft injury: a TCMR/TI Index, an AMR/MVI Index, an Activity Index and a Chronicity Index. The BanffNET Indices were evaluated against Banff diagnoses and histological indices in the training cohort and validation cohort. Since pathologist-assigned Banff lesions and diagnoses suffer from inter-observer variability, associations were also assessed with diagnoses in a multi-reader study, with time to kidney graft failure in the training and validation cohort and with biopsy-based molecular signatures in the validation cohort.

**Results:** The BanffNET TCMR/TI Index, AMR/MVI Index and Activity Index showed excellent discrimination of Banff TCMR, Banff MVI and Banff Any Diagnosis respectively, with validation AUCs ranging from 0.80 to 0.92. In addition, the BanffNET Indices explained large amounts of variability in multi-observer reference standards, time to graft failure and molecular biopsy-based diagnostics. In the large majority of cases, variability explained by BanffNET Indices matched or exceeded the variability explained by pathologist-assigned diagnoses and histological indices.

**Conclusions:** BanffNET Indices are reproducible descriptors of whole slide images that could complement and augment kidney transplant biopsy evaluation by pathologists. Associations of BanffNET Indices with multi-observer diagnoses and related outcomes suggest that they capture additional information on tissue injury type and severity on top of pathologist-assigned Banff diagnoses and histological indices.

## Introduction

Diagnostics for kidney transplants primarily consist of functional markers such as eGFR and proteinuria, which are highly prognostic for graft failure yet non-specific for disease type, and biopsy-based diagnostics, that offer insight into disease type and severity. The globally accepted framework for biopsy-based kidney transplant diagnostics is the Banff classification.^1^ It defines a set of 17 semi-quantitative lesion scores, from which 6 potentially co-existing diagnostic categories can be derived, which can be further refined into around 20 subdiagnoses in total. The two primary diagnostic categories are antibody-mediated rejection (AMR) and T-cell mediated rejection (TCMR), which are characterized histologically by microvascular and tubulointerstitial inflammation, respectively. Both the lesion scores and the diagnoses are based on thresholds, for example: the Banff i2 lesion is defined as interstitial inflammation in 26% to 50% of unscarred cortical parenchyma, and the Banff diagnosis of Acute TCMR Grade IA is defined as i ≥ 2 and t = 2. Even if these thresholds were completely optimized, they would still create situations where patients are treated differently based on falling just below or just above one of these thresholds, despite highly similar disease characteristics.^2^ To mitigate this, intermediate diagnoses have been added to the framework, increasing its complexity, requiring regular revisions, without fully resolving the underlying issue. A second shortcoming of renal allograft pathology is inter-observer variability: different pathologists assign different lesion scores to the same biopsies, which leads to reproducibility issues in research and diagnostic heterogeneity between centers in clinical practice.^3–5^

To avoid threshold effects, there is growing emphasis on continuous diagnostic indices, which take values on a granular continuous range, rather than merely differentiating between disease absence and presence. On the one hand, Banff lesions have been combined into histological continuous indices.^6,7^ On the other hand, molecular diagnostics have been developed based on transcriptomic signatures.^8–10^ While molecular indices can help reduce inter-observer variability, they come at a steep cost per sample.

Another emerging field is computational nephropathology. AI models for the analysis of whole slide images (WSIs) have been developed for image segmentation, diagnostic classification and clinical outcome prediction. So far, diagnostic applications have mainly focused on classifying biopsies into existing binary Banff diagnoses, rather than quantifying the full spectra of injury types and severity.^11^ BanffNET is a novel probabilistic deep learning framework, developed to analyze WSIs to automate the scoring of Banff lesions on a continuous scale rather than a semi-quantitative scale. Visual probability maps aid in explaining and verifying its output. The automated lesion scores do not require discretization of the severity of the lesions, are reproducible, and had stronger associations with consensus-based diagnoses, biological correlates of rejection, and clinical outcome.^12^

Where the BanffNET lesion scores provide a very detailed description of the whole slide image, derived indices are needed to reduce the complexity while retaining most of the relevant diagnostic information for use in clinical practice, trial design, and research. To this end, we introduce a set of continuous diagnostic indices derived from the BanffNET lesion scores and evaluate them against multiple reference standards, given the lack of an objective ground truth. The indices are projections of BanffNET lesion scores onto four clinically defined dimensions of disease variability in kidney transplant patients: TCMR-related and tubulointerstitial injury, AMR-related and microvascular injury, overall inflammatory activity, and overall chronic injury. The four indices provide an additional layer of information on top of biopsy evaluation by pathologists and molecular diagnostics.

## Results

### Training and validation cohorts and sample characteristics

The training cohort of the BanffNET Indices consisted of 2544 biopsies from three centers in the Netherlands (Amsterdam, Utrecht and Leiden). The validation cohort consisted of 4122 biopsies from the kidney transplant center of Leuven, Belgium, of which 3863 had complete histological data and were included in the analyses. The multi-reader study cohort consisted of 36 biopsies from the Leiden transplant center that were not overlapping with the training cohort and that were scored by 67 pathologists, resulting in a total of 2412 biopsy scorings. The biopsy WSIs were additionally scored by BanffNET. Patient and biopsy characteristics of the training cohort and validation cohort are reported in **Table 1**. Characteristics of the full validation cohort and of the complete-case subset are reported in **Supplementary Table 1**. For each biopsy, Banff diagnoses and derived categories were defined as described in the **Online Methods**. In addition, BanffNET automated lesion scores of t, i, g, ptc, v, tma, ati, ti, t-ifta, i-ifta, ifta, cg, mm, ah, cv, gs and fsgs were computed for all biopsies, using the algorithms described in Buzzanca et al.^12^ These lesion scores range from 0 to 1 on a continuous scale.

**Table 1.** Transplant and biopsy characteristics of training cohort and validation cohort.

| Characteristic | Training cohort |  |  | Validation cohort |
| --- | --- | --- | --- | --- |
|  | Amsterdam | Utrecht | Leiden | Leuven |
| **Transplant characteristics** | **n = 1048** | **n = 583** | **n = 913** | **n = 1340** |
| Male recipient sex, % | 60% | 60% | 64% | 63% |
| Recipient age, mean (SD) | 47 (16) | 50 (16) | 52 (14) | 54 (13) |
| Recipient BMI, mean (SD) | NA<sup>a</sup> | NA<sup>a</sup> | NA<sup>a</sup> | 25 (4) |
| White European recipient ethnicity, % | NA<sup>a</sup> | NA<sup>a</sup> | NA<sup>a</sup> | 97% |
| Repeated transplantation, % | 15% | 16% | 13% | 15% |
| Pre-transplant HLA-DSA | NA<sup>a</sup> | NA<sup>a</sup> | NA<sup>a</sup> | 10% |
| Male donor sex, % | 49% | 51% | 45% | 53% |
| Donor age, mean (SD) | 49 (14) | 52 (14) | 52 (14) | 49 (15) |
| Deceased donor (brain death), % | 39% | 29% | 16% | 75% |
| Deceased donor (cardiac death), % | 24% | 32% | 28% | 18% |
| Living donor, % | 37% | 39% | 56% | 7% |
| CIT in hours, mean (SD) | 13 (9) | 12 (9) | 8 (7) | 13 (6) |
| Total number of HLA A/B/DR mismatches, median (IQR) | 3 (2-4) | 3 (2-4) | 4 (3-5) | 3 (2 – 4) |
| Missing, % | 0% | 0% | 0% | 30% |
| Total follow-up time in years, median (IQR) among non-missing | 6.3 (3.2 – 10.3) | 5.0 (2.1 – 9.3) | 5.7 (2.8 – 9.5) | 7.8 (4.0 – 11.4) |
| Missing, % | 10% | 20% | 49% | 0% |
| Graft failure, % | 30% | 18% | 8% | 16% |
| TCMR at any time during follow-up, % | NA<sup>b</sup> | NA<sup>b</sup> | NA<sup>b</sup> | 20% |
| AMR at any time during follow-up, % | NA<sup>b</sup> | NA<sup>b</sup> | NA<sup>b</sup> | 11% |
| MVI<sub>DSA-/C4d-</sub> at any time during follow-up, % | NA<sup>b</sup> | NA<sup>b</sup> | NA<sup>b</sup> | 12% |
| BKPyVAN at any time during follow-up, % | NA<sup>b</sup> | NA<sup>b</sup> | NA<sup>b</sup> | 8% |
| HLA-DSA at any time during follow-up, % | NA<sup>b</sup> | NA<sup>b</sup> | NA<sup>b</sup> | 13% |
| **Biopsy characteristics** | **n = 1048** | **n = 583** | **n = 913** | **n = 3863** |
| Days from transplantation to biopsy, median (IQR) | 367 (42 – 886) | 225 (70 – 1487) | 177 (13 – 1028) | 363 (89 – 740) |
| Missing, % | 10% | 20% | 49% | 0% |
| Years from biopsy to end of follow-up, median (IQR) | 4.0 (1.2 – 7.9) | 2.7 (1.2 – 5.1) | 3.1 (1.0 – 6.4) | 7.9 (4.0 – 11.0) |
| Missing, % | 10% | 20% | 49% | 0% |
| Indication biopsy, % | NA<sup>a</sup> | NA<sup>a</sup> | NA<sup>a</sup> | 28% |
| HLA-DSA presence, % of non-missing | 20% | 14% | 27% | 13% |
| Missing, % | 39% | 55% | 69% | 0% |
| C4d (binary), % of non-missing | 14% | 13% | 19% | 5% |
| Missing, % | 34% | 31% | 60% | 0% |
| Banff 2022 categories |  |  |  |  |
| No Diagnosis, % of non-missing | 35% | 26% | 7% | 70% |

| **Biopsy characteristics** | **Amsterdam**<br>**n = 1048** | **Utrecht**<br>**n = 583** | **Leiden**<br>**n = 913** | **Leuven**<br>**n = 3863** |
| --- | --- | --- | --- | --- |
| Missing, % | 38% | 36% | 39% | 0% |
| TCMR, % of non-missing | 21% | 19% | 32% | 9% |
| Missing, % | 3% | 4% | 2% | 0% |
| Borderline Changes, % of non-missing | 8% | 24% | 18% | 8% |
| Missing, % | 3% | 4% | 3% | 0% |
| AMR/MVI, % of non-missing | 33% | 31% | 64% | 13% |
| Missing, % | 53%<sup>c</sup> | 57%<sup>c</sup> | 69%<sup>c</sup> | 0% |
| AMR, % of non-missing | 22% | 21% | 38% | 8% |
| Missing, % | 53%<sup>c</sup> | 57%<sup>c</sup> | 69%<sup>c</sup> | 0% |
| Probable AMR, % of non-missing | 1% | 2% | 4% | 2% |
| Missing, % | 55% | 59% | 73% | 0% |
| MVI, % of non-missing | 20% | 20% | 38% | 12% |
| Missing, % | 0% | 0% | 0% | 0% |
| MVI<sub>DSA-/C4d-</sub>, % of non-missing | 11% | 11% | 30% | 6% |
| Missing, % | 56% | 59% | 73% | 0% |
| BKPyVAN, % of non-missing | 5% | 3% | 2% | 3% |
| Missing, % | 10% | 20% | 49% | 0% |
| Mixed Rejection, % of non-missing | 5% | 6% | 10% | 3% |
| Missing, % | 14% | 13% | 21% | 0% |

Abbreviations: no., number; IQR, interquartile range; BMI, body mass index; HLA-DSA, anti-human leukocyte antigen donor specific antibodies; CIT, cold ischemia time; TCMR, T-cell mediated rejection; AMR, antibody-mediated rejection; MVI<sub>DSA-/C4d-</sub>, C4d negative DSA negative microvascular inflammation; BKPyVAN, BK polyomavirus nephropathy.
<sup>a</sup>Recipient BMI, recipient ethnicity, pre-transplant HLA-DSA and biopsy type were not available for the training cohort.
<sup>b</sup>The training cohort included only a single biopsy per transplant, hence the diagnoses per transplant are the same as the diagnoses per biopsy.
<sup>c</sup>Diagnosis of AMR requires availability of HLA-DSA and C4d, which was limited in the training cohort.

In the training cohort, living donation accounted for 37% of transplants in Amsterdam, 39% in Utrecht and 56% in Leiden, with the remaining transplants from donation after brain death (39%, 29% and 16% respectively) and donation after cardiac death (24%, 32% and 28% respectively). Repeat transplantation was performed in 15%, 16% and 13% of transplants respectively. Median time from transplantation to biopsy was 367 days (IQR 42 – 886) in Amsterdam, 225 days (IQR 70 – 1487) in Utrecht and 177 days (IQR 13 – 1028) in Leiden. Among biopsies with available data, TCMR was diagnosed in 21%, 19% and 32% of biopsies, Borderline Changes in 8%, 24% and 18%, AMR in 22%, 21% and 38%, Probable AMR in 1%, 2% and 4%, MVI_DSA-/C4d-_ in 11%, 11% and 30%, BKPyVAN in 5%, 3% and 2% and Mixed Rejection in 5%, 6% and 10%. Recipient BMI, recipient ethnicity and biopsy type were not available for the training cohort (**Table 1**).

Most transplants in the validation cohort were from donation after brain death (75%), followed by donation after cardiac death (18%), and living donation (7%). Repeat transplantation was performed in 15% of transplants, and 10% had pre-transplantation HLA-DSA. 97% of recipients were of White European ethnicity. Median time from transplantation to biopsy was 363 days (IQR 89 – 740) and median time from biopsy to end of follow-up was 7.9 years (IQR 4.0 – 11.0). 28% of the biopsies were indication biopsies. TCMR was diagnosed in 9% of biopsies, Borderline Changes in 8% of biopsies, AMR in 8% of biopsies, Probable AMR in 2% of biopsies, MVI_DSA-/C4d-_ in 6% of biopsies, BKPyVAN in 3% of biopsies and Mixed Rejection in 3% of biopsies (**Table 1**).

### Development of BanffNET continuous Indices from BanffNET lesion scores

From the 17 BanffNET lesion scores, four BanffNET continuous Indices were developed: a TCMR/TI Index to quantify TCMR-related and tubulointerstitial injury, an AMR/MVI Index to quantify AMR-related and microvascular injury, an Activity Index to quantify overall inflammatory activity and a Chronicity Index to quantify overall chronic injury (**Table 2**). By design, the indices have mean zero and a standard deviation of one in the training cohort and represent spectra of injury severity. The indices were constructed using Ridge-penalized regression. For the TCMR/TI Index, the target variable of the regression was the Banff diagnosis of TCMR. For the AMR/MVI Index, the target variable was diagnosis of AMR/MVI. For the Activity Index and Chronicity Index, the target variable was the hazard of kidney graft failure. Similarly to previously proposed continuous indices^6^, each Index was trained on an *a priori* selection of lesions that were considered clinically relevant for the disease dimension that is represented by the index. In addition, stepwise backward selection was performed. Additional details are provided in the **Online Methods**.

**Table 2.**
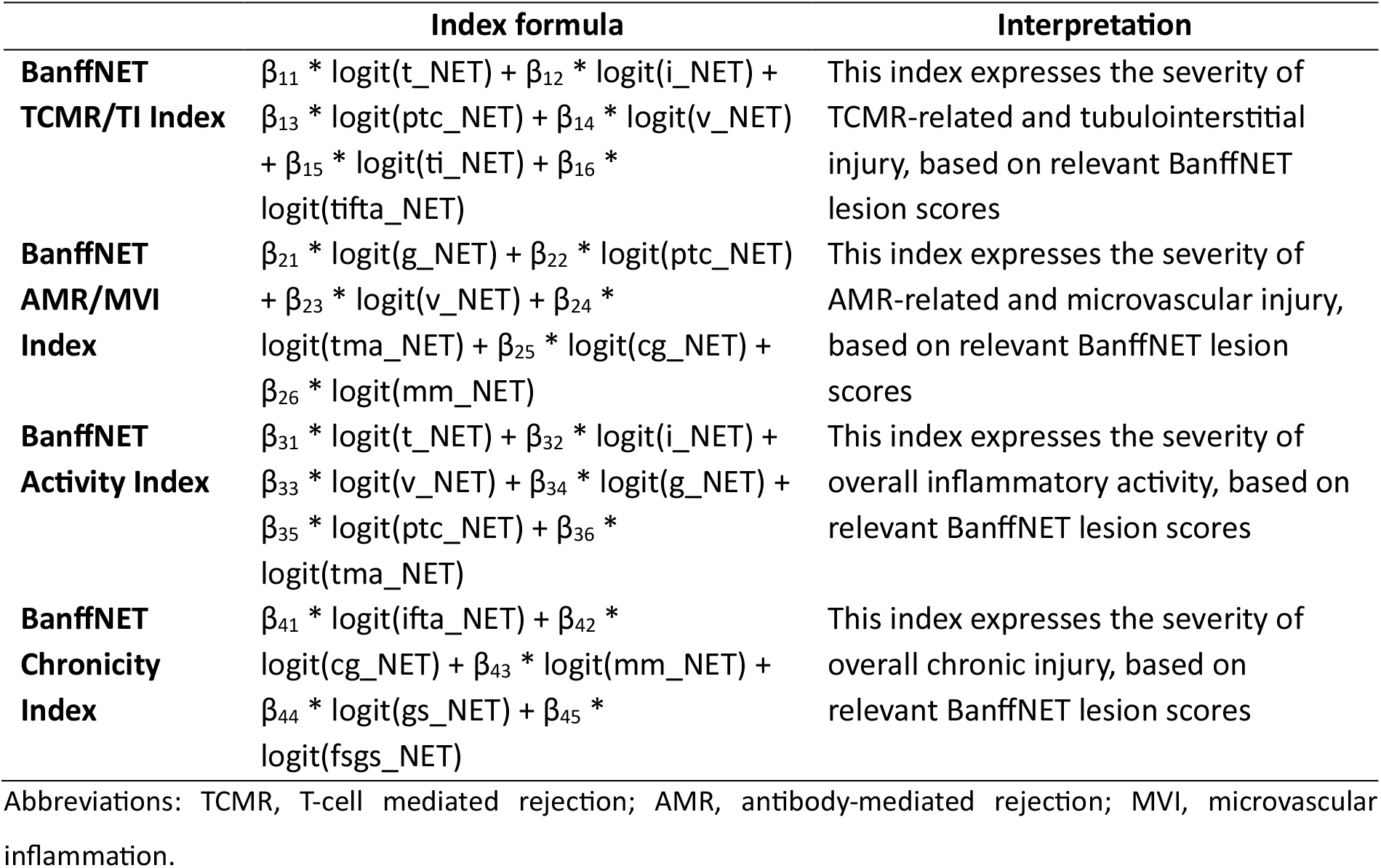
Definitions of the BanffNET Indices.

### The BanffNET Indices in relation to pathologist-assigned Banff diagnoses and histological indices

The Banff classification is considered the current gold standard for diagnosis of rejection, infection and scarring in kidney grafts. Nevertheless, it suffers from inter-observer variability.^5^ Therefore, comparisons of novel diagnostics with Banff reference standards can never reach perfect accuracy, and discrepancies may be caused by imperfections in the reference standard rather than in the novel diagnostic. Still, such comparisons are useful to detect extreme discrepancies, which are unlikely to be caused purely by limitations in the Banff reference standards.

#### Distribution of the indices

The distribution of the BanffNET Indices across Banff diagnostic groups in the training cohort is shown in **Figure 1**. The TCMR/TI Index increased from No Diagnosis to Borderline Changes to TCMR. Some BKPyVAN cases also had a moderate TCMR/TI Index value, which was expected since both TCMR and BKPyVAN are characterized by tubulointerstitial inflammation. The AMR/MVI Index gradually increased from No Diagnosis to Probable AMR to Mixed Rejection, MVI_DSA-/C4d-_ and AMR. The Activity Index gradually increased from No Diagnosis to fully inflamed categories (AMR, MVI_DSA-/C4d-_, TCMR and Mixed Rejection), with intermediate categories (Borderline Changes and Probable AMR) in between (**Figure 1**). The Chronicity Index was only slightly elevated in AMR and MVI_DSA-/C4d-_ biopsies. Distributions in the validation cohort were similar, except for the Chronicity Index, which was not elevated in any of the diagnostic categories (**Supplementary Figure 1**). The distribution of Banff categories in the 2-dimensional diagnostic space defined by the BanffNET TCMR/TI Index and the BanffNET AMR/MVI Index indicated substantial heterogeneity within Banff categories. The high proportion of protocol biopsies in the validation cohort produced less variability in the upper range of the BanffNET Indices compared to the training cohort (**Supplementary Figure 2**, **Supplementary Figure 3**).

**Figure 1.**
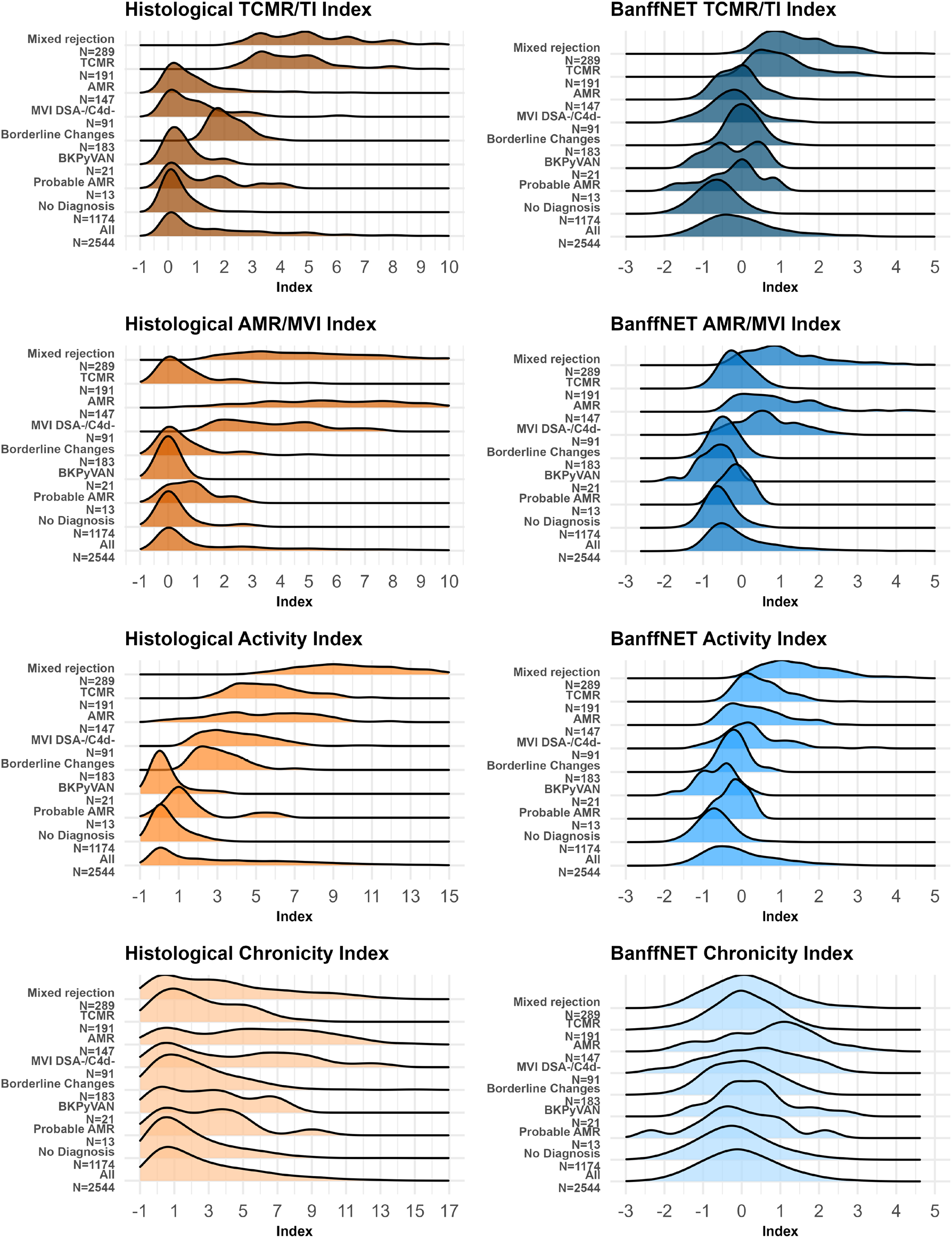
Distributions of histological indices and BanffNET Indices within Banff categories in the training cohort (n = 2544). Only exclusive diagnoses were included in the categories to prevent overlap.

#### Associations of the indices with each other and with Banff diagnoses

While the distributions of the indices showed intuitive patterns across the Banff categories, correlations provide additional insights. The correlation between the BanffNET TCMR/TI Index and BanffNET AMR/MVI Index was stronger than the correlation between the histological TCMR/TI index and histological AMR/MVI index, likely due to the denoising obtained by the BanffNET algorithms (**Supplementary Figure 4-5**). In the training data, correlations of the BanffNET Indices with their corresponding Banff diagnoses and histological indices were stronger than in the validation data (**Supplementary Figure 4-5**). When looking beyond correlation at linearity of the associations of BanffNET Indices with histological indices, any deviations from linearity were negligible (**Figure 2-3**). Further, associations of the BanffNET Indices with the log-odds of Banff diagnoses were slightly concave in the training cohort but close to linear in the validation cohort (**Supplementary Figure 6**).

**Figure 2.**
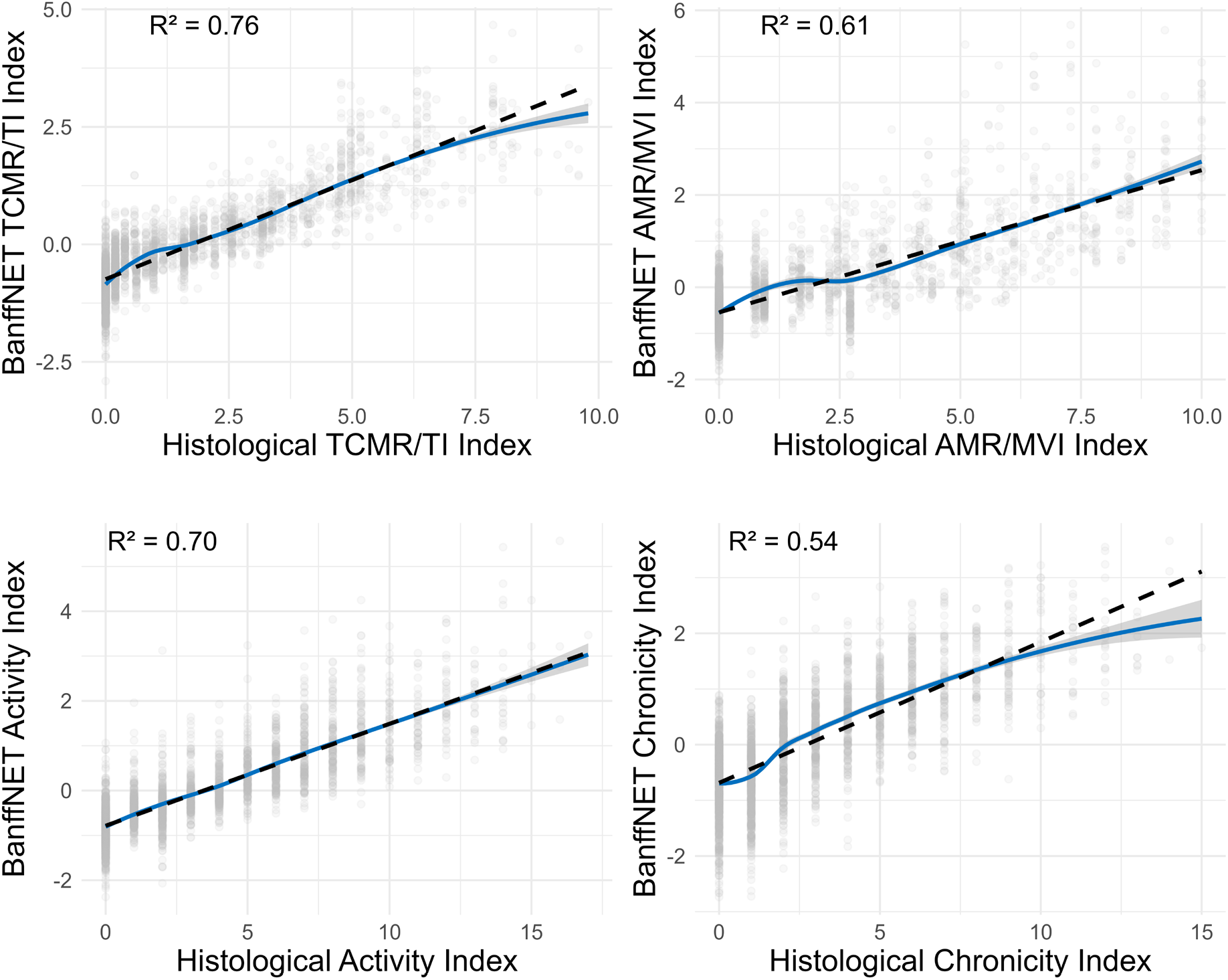
Linearity of associations with histological indices in the training cohort (n = 2544). Loess-smoothed scatter plots with pointwise 95% confidence intervals, to evaluate linearity in the associations of the BanffNET Indices with corresponding histological indices.

**Figure 3.**
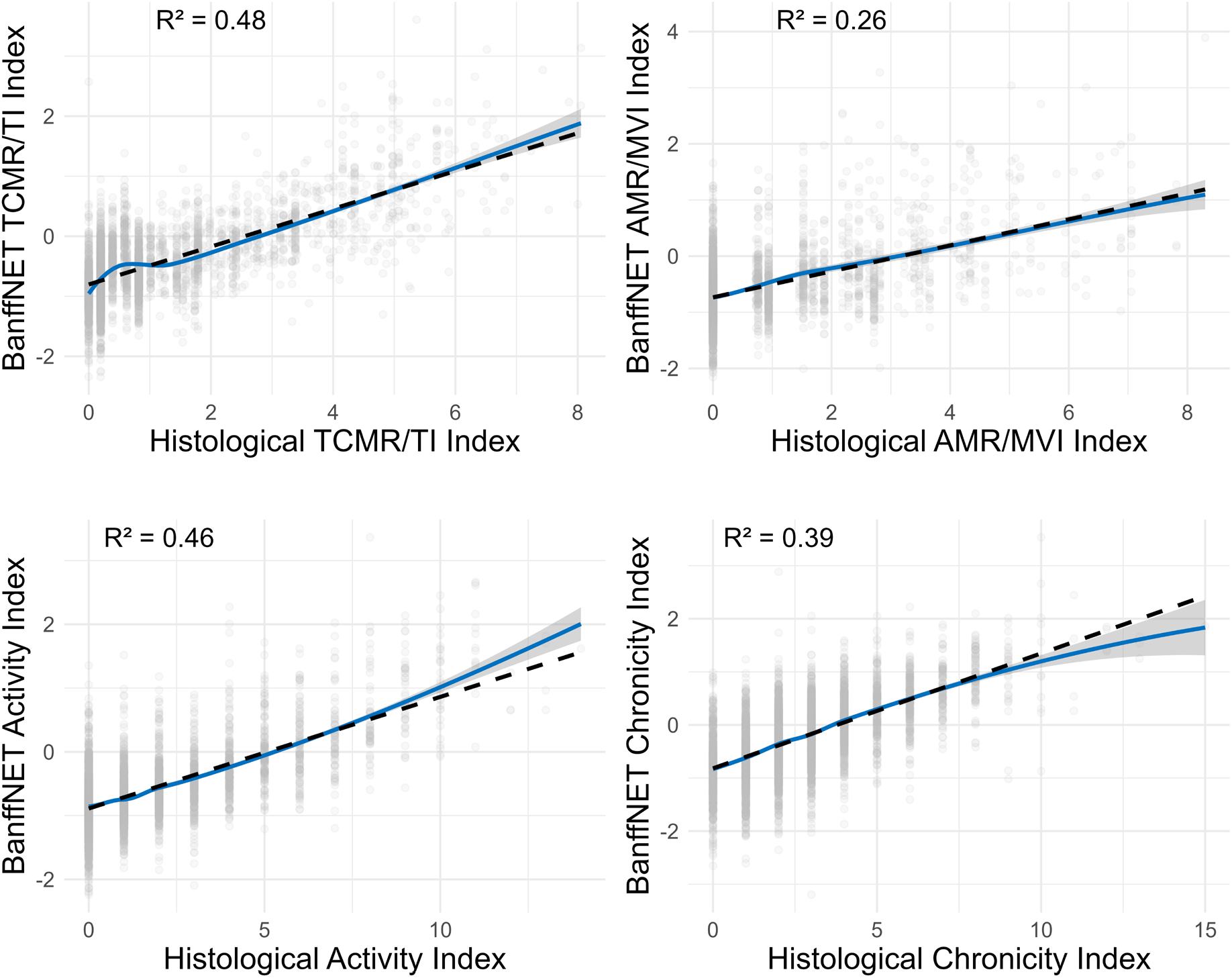
Linearity of associations with histological indices in the validation cohort (n = 3863). Loess-smoothed scatter plots with pointwise 95% confidence intervals to evaluate linearity in the associations of the BanffNET Indices with corresponding histological indices.

#### Discrimination

For a thorough evaluation of the diagnostic performance, we assessed discrimination through areas under the receiver operating characteristic curve (AUCs). The BanffNET TCMR/TI Index was tested against Banff TCMR, the BanffNET AMR/MVI Index was tested against diagnosis of MVI, and the BanffNET Activity Index was tested against the category ‘Any Diagnosis’. The BanffNET Chronicity Index was not included in this evaluation due to the lack of a binary Banff diagnosis for chronic injury. The AUCs of the BanffNET TCMR/TI Index for the Banff TCMR diagnosis were 0.96 (95% CI 0.95 – 0.97) and 0.92 (95% CI 0.91 – 0.94) in the training and validation cohort respectively. The BanffNET AMR/MVI Index had AUCs of 0.94 (95% CI 0.93 – 0.95) and 0.80 (95% CI 0.77 – 0.82) for the diagnosis of MVI in the respective cohorts. The BanffNET Activity Index had AUCs of 0.92 (95% CI 0.91 – 0.93) and 0.83 (95% CI 0.82 – 0.85) for Banff Any Diagnosis in the respective cohorts (**Figure 4**).

**Figure 4.**
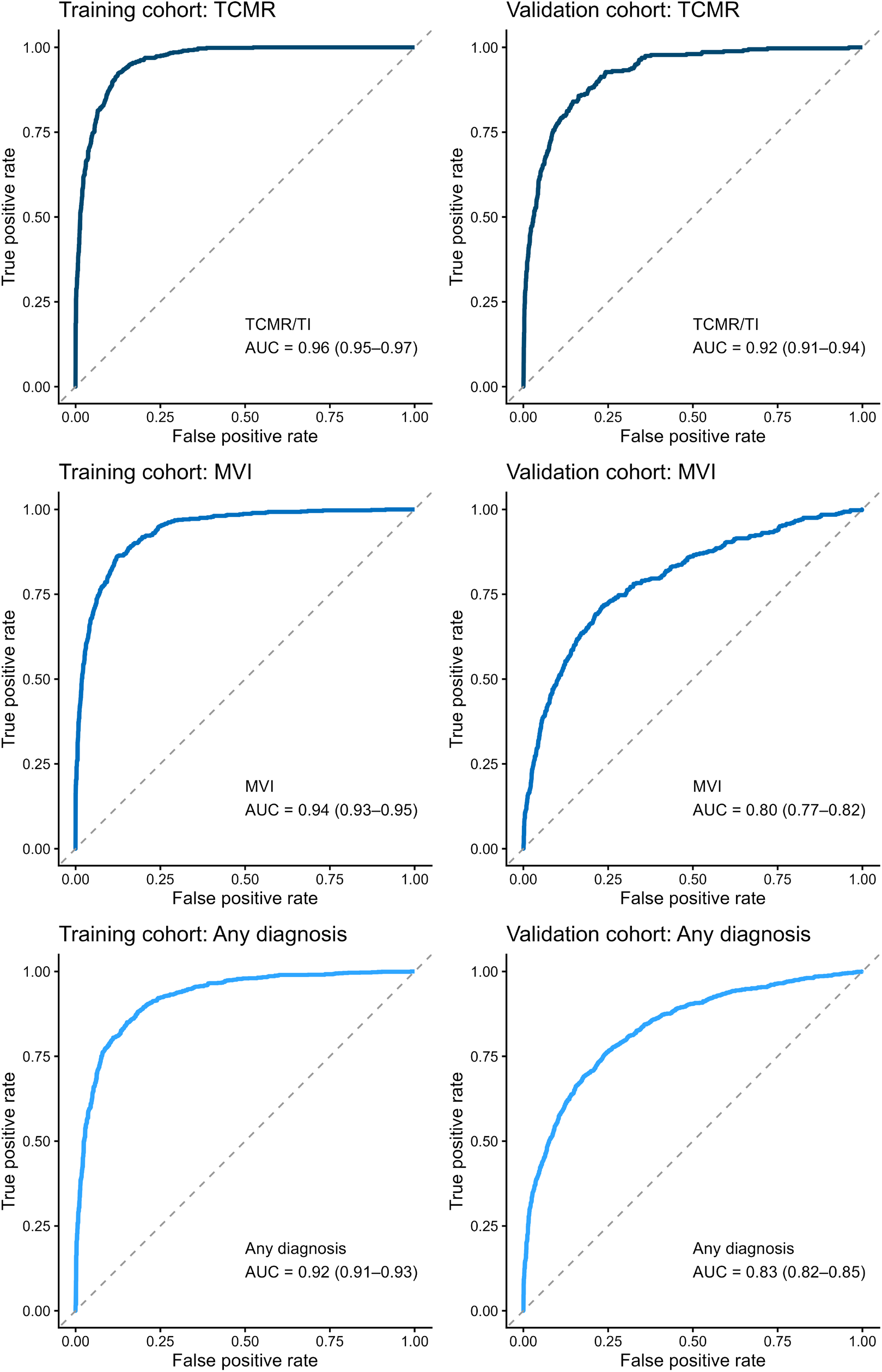
Discrimination of Banff diagnoses by BanffNET Indices. ROC curves of BanffNET TCMR/TI, AMR/MVI and Activity Indices for Banff TCMR, MVI and Any Diagnosis in the training cohort (n = 2544) and validation cohort (n = 3863).

#### Calibration

An external calibration analysis was performed to assess whether the BanffNET Indices remained aligned with the Banff diagnoses in the validation cohort. Calibration intercepts close to zero indicate low overall discrepancy and calibration slopes close to 1 indicate low over– or underfitting. The calibration intercepts were close to zero for MVI and Any Diagnosis (MVI calibration intercept 0.13, 95% CI –0.00 – 0.26; Any Diagnosis calibration intercept 0.06, 95% CI –0.03 – 0.15), while the TCMR intercept of 0.25 (95% CI 0.10 – 0.40) indicated a slight discrepancy between the BanffNET-predicted TCMR risk and the Banff TCMR risk, with the latter being higher than the former on average. Further, the calibration slopes for TCMR, MVI and Any Diagnosis were smaller than 1 (TCMR calibration slope 0.84, 95% CI 0.77 – 0.91; MVI calibration slope 0.50, 95% CI 0.45 – 0.55; Any Diagnosis calibration slope 0.78, 95% CI 0.72 – 0.84), indicating potential overfitting on the training data, especially of the BanffNET AMR/MVI Index. Flexible calibration curves showed that the largest discrepancy occurred in the upper range of the BanffNET AMR/MVI Index, where the observed probability of MVI was lower than the BanffNET-predicted probability (**Figure 5**).

**Figure 5.**
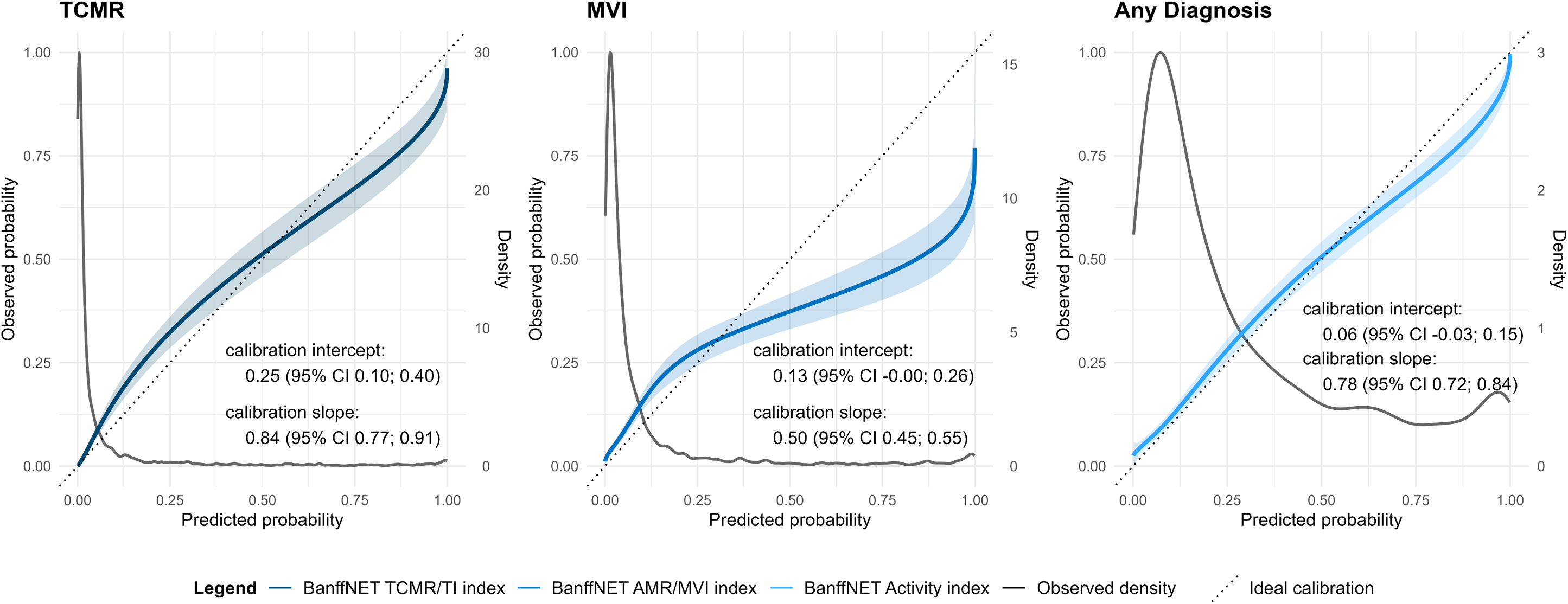
Calibration of BanffNET Indices in the validation cohort (n = 3863). Flexible calibration curves with pointwise 95% intervals for the observed event probability at a given predicted probability, as well as calibration intercepts and slopes, to quantify systematic discrepancies between BanffNET Indices and Banff diagnoses in the validation cohort. Density curves represent densities of the predicted probabilities. Since calibration analysis for binary outcomes requires risk predictions, the BanffNET Indices were transformed into risk predictions of Banff TCMR, Banff MVI and Banff Any Diagnosis.

#### Classification metrics after dichotomizations

Classification metrics were obtained after dichotomization of the BanffNET Indices. Dichotomization was necessary to calculate traditional diagnostic accuracy metrics, and dichotomized BanffNET Indices could be used in very specific applications such as inclusion/exclusion into studies or selection for further review. However, the Indices were developed primarily to provide more granular information than the existing binary diagnoses, and we recommend interpreting them on their original scale whenever possible. Three dichotomization thresholds were chosen for each Index, based on their sensitivity and specificity in the training cohort for the corresponding Banff diagnosis (Banff TCMR for the BanffNET TCMR/TI Index, Banff MVI for the BanffNET AMR/MVI Index, and Banff Any Diagnosis for the BanffNET Activity Index). The lowest threshold was based on 5% false negatives, the second threshold was balanced according to Youden’s J and the highest threshold was based on 5% false positives, which minimizes the number of false positives. The thresholds based on 5% false negatives were 0.20 for the BanffNET TCMR/TI Index, –0.18 for the BanffNET AMR/MVI Index and –0.55 for the BanffNET Activity Index in the training cohort. These thresholds resulted in specificities of 0.84, 0.75 and 0.66, in positive predictive values (PPV) of 0.65, 0.59 and 0.77, in negative predictive values (NPV) of 0.98, 0.98 and 0.92, and in percentages of biopsies under the threshold of 65%, 56%, and 33% respectively. In the validation cohort, these same thresholds resulted in sensitivities of 0.67, 0.52 and 0.73, in specificities of 0.95, 0.89 and 0.78, in PPVs of 0.52, 0.38 and 0.58, in NPVs of 0.97, 0.93 and 0.87, and in percentages of biopsies under the threshold of 88%, 84%, and 63% respectively (**Table 3**). Metrics for the dichotomizations according to Youden’s J and 5% false positives are shown in **Table 3**. While discrimination and calibration analyses for Banff diagnoses offer insight into discrepancies between BanffNET Indices and Banff diagnoses, they do not tell us which of the two is closer to the unobserved ground truth. Therefore, we next compared the BanffNET Indices with pathologist-assigned diagnoses and indices in terms of explained variability in four independent reference standards.

**Table 3.** Diagnostic accuracy of dichotomized BanffNET Indices for Banff diagnoses in the training cohort (n = 2544) and validation cohort (n = 3863).

|  |  | Training cohort (n = 2544) |  |  |  |  |  | Validation cohort (n = 3863) |  |  |  |  |  |
| --- | --- | --- | --- | --- | --- | --- | --- | --- | --- | --- | --- | --- | --- |
|  | Threshold | AUC (95% CI) | Sensitivity | Specificity | PPV | NPV | Biopsies under threshold (%) | AUC (95% CI) | Sensitivity | Specificity | PPV | NPV | Biopsies under threshold (%) |
| **BanffNET TCMR/TI Index versus Banff TCMR** |  |  |  |  |  |  |  |  |  |  |  |  |  |
| 5% false negatives | 0.20 | 0.96<br>(0.95-0.97) | 0.95 | 0.84 | 0.65 | 0.98 | 64.76 | 0.92<br>(0.91-0.94) | 0.67 | 0.95 | 0.52 | 0.97 | 88.06 |
| Balanced (Youden's J) | 0.28 | 0.96<br>(0.95-0.97) | 0.92 | 0.87 | 0.70 | 0.97 | 68.28 | 0.92<br>(0.91-0.94) | 0.64 | 0.95 | 0.56 | 0.96 | 89.41 |
| 5% false positives | 0.60 | 0.96<br>(0.95-0.97) | 0.75 | 0.95 | 0.83 | 0.92 | 78.26 | 0.92<br>(0.91-0.94) | 0.44 | 0.98 | 0.70 | 0.95 | 94.23 |
| **BanffNET AMR/MVI Index versus Banff MVI** |  |  |  |  |  |  |  |  |  |  |  |  |  |
| 5% false negatives | -0.18 | 0.94<br>(0.93-0.95) | 0.95 | 0.75 | 0.59 | 0.98 | 56.33 | 0.80<br>(0.77-0.82) | 0.52 | 0.89 | 0.38 | 0.93 | 84.13 |
| Balanced (Youden's J) | 0.09 | 0.94<br>(0.93-0.95) | 0.86 | 0.88 | 0.72 | 0.94 | 67.85 | 0.80<br>(0.77-0.82) | 0.38 | 0.95 | 0.47 | 0.92 | 90.76 |
| 5% false positives | 0.47 | 0.94<br>(0.93-0.95) | 0.69 | 0.95 | 0.84 | 0.89 | 77.83 | 0.80<br>(0.77-0.82) | 0.23 | 0.97 | 0.53 | 0.91 | 94.90 |
| **BanffNET Activity Index versus Banff Any Diagnosis** |  |  |  |  |  |  |  |  |  |  |  |  |  |
| 5% false negatives | -0.55 | 0.92<br>(0.91-0.93) | 0.95 | 0.66 | 0.77 | 0.92 | 32.73 | 0.83<br>(0.82-0.85) | 0.73 | 0.78 | 0.58 | 0.87 | 62.52 |
| Balanced (Youden's J) | -0.27 | 0.92<br>(0.91-0.93) | 0.85 | 0.85 | 0.87 | 0.83 | 46.95 | 0.83<br>(0.82-0.85) | 0.52 | 0.91 | 0.72 | 0.82 | 78.38 |
| 5% false positives | 0.08 | 0.92<br>(0.91-0.93) | 0.65 | 0.95 | 0.94 | 0.70 | 62.44 | 0.83<br>(0.82-0.85) | 0.32 | 0.98 | 0.86 | 0.77 | 88.83 |

Dichotomizations were performed based on 5% false negatives, Youden's J and 5% false positives in the training cohort. Abbreviations: AUC, area under the receiver operating characteristic curve; CI, confidence interval; PPV, positive predictive value; NPV, negative predictive value; TCMR, T-cell mediated rejection; MVI, microvascular inflammation.

### BanffNET Indices in relation to robust multi-observer reference standards

To address the inter-observer variability inherent to the Banff classification, robust Banff reference standards were constructed using data from the LUMC Global Reader Study^5^ where 67 pathologists scored the same 36 biopsies. Banff TCMR, MVI and Any Diagnosis were aggregated over all pathologists, resulting in more robust multi-observer Banff reference standards for TCMR, MVI and Any Diagnosis. The same 36 biopsies were analyzed by BanffNET, producing BanffNET lesion scores, which were transformed to the BanffNET Indices. Individual pathologists were then compared with BanffNET in terms of explained variability in the multi-observer reference standards, which is a measure of the association strength in linear models. Methodological details can be found in the **Online Methods**. The BanffNET TCMR/TI Index showed a stronger association with the multi-observer Banff TCMR reference standard than the average individual pathologist’s Banff TCMR diagnosis or histological TCMR/TI index. Combining the BanffNET TCMR/TI Index with the pathologist-assigned histological TCMR/TI index and Banff TCMR diagnosis increased the explained variability even further. Each of these differences was statistically significant. The BanffNET AMR/MVI Index explained a similar amount of variability in the multi-observer MVI reference standard as the average individual pathologist’s Banff MVI diagnosis and histological AMR/MVI index, and the combination of all three explained significantly more variability than any single diagnostic. The same was true for the explained variability in the multi-observer Banff Any Diagnosis reference standard: the BanffNET Activity Index performed similarly to the average individual pathologist’s Banff Any Diagnosis and histological Activity index, and the combination of all three produced a significantly higher explained variability (**Figure 6, Supplementary Table 2, Supplementary Figure 7**). The associations of the BanffNET Indices with this multi-observer reference standard were approximately linear, with small deviations in the extreme upper range, where an additional increase in the BanffNET Indices was no longer associated with an increase in the multi-observer Banff reference standard (**Supplementary Figure 8**). A second set of multi-observer reference standards was obtained by aggregating the histological indices over all pathologists. Here, BanffNET explained more variability in the multi-observer reference standards than pathologist-assigned Banff TCMR and Banff Any Diagnosis, but on average, it explained less variability than individual pathologists’ histological AMR/MVI and Activity indices, likely partly due to the role of C4d in these indices, which is not measured directly by BanffNET since it requires immunohistochemistry or immunofluorescence rather than mere histology (morphology) (**Figure 6, Supplementary Table 3, Supplementary Figure 9**).

**Figure 6.**
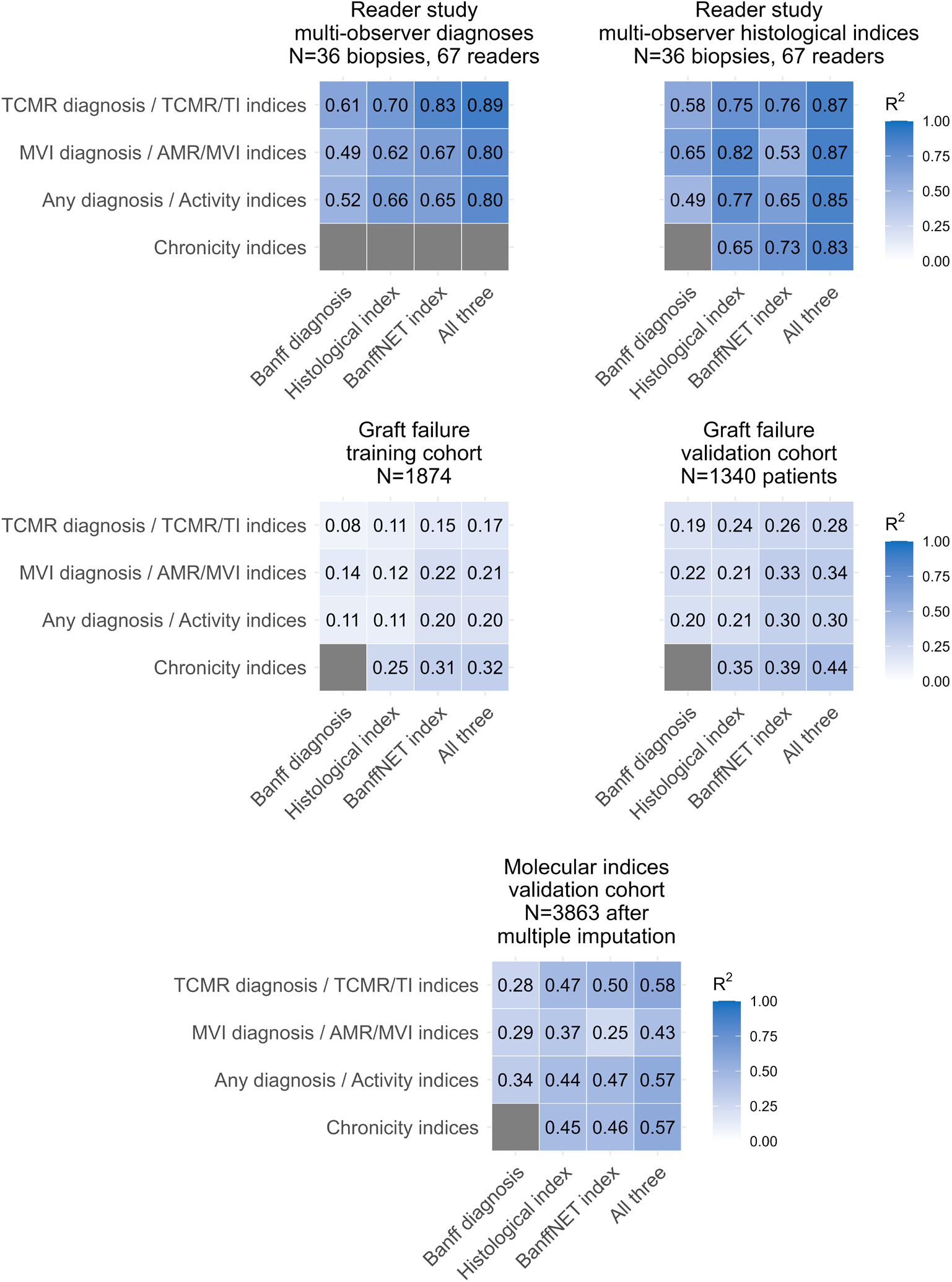
Comparison of Banff diagnoses, histological indices, BanffNET Indices and a combination of all three in terms of explained variability in various reference standards. Reference standards consisted of multi-observer diagnoses and multi-observer histological indices in the LUMC Global Reader Study, time to graft failure in the training and validation cohorts, and molecular indices in the validation cohort. Estimates denote R² values, except for graft failure, where Royston-Sauerbrei pseudo-R² was used. Grey cells indicate that no corresponding Banff diagnosis exists for the Chronicity indices.

### BanffNET Indices in relation to immunological factors

Where a Banff AMR diagnosis is based on histological lesions, C4d deposition and HLA-DSA presence, the histological AMR/MVI index is based on histological lesions and C4d, and the BanffNET AMR/MVI Index describes information in the WSI only. As a result, the association of the BanffNET AMR/MVI Index with C4d was weaker than that of Banff AMR and the histological AMR/MVI index in both cohorts, and its association with HLA-DSA was weaker in the validation cohort, while in the training cohort it was comparable to that of Banff AMR (**Supplementary Table 4**), highlighting the importance of using complementary sources of information.

### BanffNET Indices in relation to kidney graft failure

In the training cohort, all four BanffNET Indices explained more variability in time to kidney graft failure than their individual pathologist-assigned counterparts. The combination of BanffNET Indices with pathologist-assigned diagnoses and indices did not explain more variability than the BanffNET Indices by themselves. In the validation cohort, there was no significant difference between the explained variability by the BanffNET TCMR/TI Index, histological TCMR/TI index and Banff TCMR, but the BanffNET AMR/MVI Index and BanffNET Activity Index did outperform their pathologist-assigned counterparts in this respect. The BanffNET Chronicity Index did not explain more variability than the histological Chronicity index, though the combination of both did explain more variability than each index separately (**Figure 6**, **Supplementary Table 5-6, Supplementary Figure 10-11**). The BanffNET Indices also explained significant residual variability in time to graft failure in biopsies without any Banff diagnoses specifically (**Supplementary Table 5-6, Supplementary Figure 12-13**). It should be noted that patients were treated according to the Banff diagnoses, and not according to the BanffNET Indices, possibly skewing the comparison in terms of explained variability in graft failure in favor of the BanffNET Indices. Also, while the estimates for the BanffNET Activity Index and BanffNET Chronicity Index in the training cohort may be optimistic since these indices were trained directly on time to graft failure in this cohort, the associations generalized well to the validation cohort. The associations of the indices with the hazard of kidney graft failure were close to linear, except in the extreme upper range of the BanffNET AMR/MVI Index, where further increases in the index were not associated with differences in the hazard (**Supplementary Figure 14-15**).

### BanffNET Indices in relation to molecular signatures

Next, BanffNET Indices were compared to Banff diagnoses and histological indices in terms of their associations with recently proposed molecular indices constructed on biopsy gene expression data from the BHOT panel in a subset of the validation cohort, as described in Naesens et al.^10^ Multiple imputation was used to make results generalizable to the entire cohort and by extension the clinical population of interest. Because the molecular indices were trained on this exact data, the histological indices are advantaged in this comparison. Nevertheless, BanffNET TCMR/TI Index, Activity Index and Chronicity Index R² values closely resemble those of the histological indices. Only the BanffNET AMR/MVI Index explains less variability in the molecular AMR/MVI index than the histological AMR/MVI index (**Figure 6, Supplementary Table 7, Supplementary Figure 16**). The associations between the BanffNET Indices and the corresponding molecular indices were approximately linear, except in the extreme upper range of the BanffNET AMR/MVI Index, where higher values no longer corresponded to increases in the molecular AMR/MVI Index (**Supplementary Figure 17**).

## Discussion

In this paper, we presented four diagnostic indices for kidney transplant patients, derived from BanffNET automated lesion scores. The BanffNET TCMR/TI Index quantifies the severity of injury characteristic of TCMR and TI. The BanffNET AMR/MVI Index quantifies the severity of injury characteristic of AMR and MVI. The BanffNET Activity Index quantifies the severity of acute injury in kidney transplant biopsies and the BanffNET Chronicity Index quantifies the severity of chronic injury in kidney transplant biopsies. These indices provide a description of the WSI, which is independent of and complementary to the context-aware assessment of the pathologist.

The BanffNET TCMR/TI Index and BanffNET AMR/MVI Index were trained on the Banff diagnoses of TCMR and AMR/MVI respectively. The BanffNET Activity Index and the BanffNET Chronicity Index were both trained on the hazard of graft failure, while being restricted to active lesion scores and chronic lesion scores respectively, to capture the overall active and chronic histological injury that actually associates with increased risk of graft failure.

When dichotomized, the BanffNET Indices become close approximations of their corresponding Banff diagnoses. However, when the indices are used as continuous scores, they retain more relevant information. When 67 pathologists scored the same biopsies, the addition of BanffNET Indices provided a significant increase in the explained variability of the multi-pathologist reference standard, compensating for the large inter-observer variability between pathologists. Moreover, the BanffNET Indices often explained more variability in time to kidney graft failure than single-pathologist Banff diagnoses and histological indices. The BanffNET TCMR/TI, Activity and Chronicity Indices also correlated well with molecular indices derived from the BHOT panel, with R² values comparable to the histological indices.

Similar to molecular diagnostics for kidney transplants, BanffNET offers an objective, reproducible summary of acute and chronic kidney injury. However, where molecular diagnostics are relatively opaque and expensive, BanffNET is inexpensive to run and can provide visualizations of relevant areas on the WSI, ensuring transparency and explainability.

The implementation of AI-based diagnostic tools in clinical practice requires careful consideration. While we demonstrated that BanffNET Indices explain as much or more variability in various reference standards compared to pathologist-assigned diagnoses and indices, the goal of this study was not to assess clinical utility. Future studies should investigate how exactly BanffNET can be implemented in clinical trial contexts and in routine clinical practice, in a way that leverages the respective strengths of pathologists and BanffNET. It will be key to strike a balance between overreliance and underreliance on these AI diagnostics, while also preventing deskilling of pathologists. Therefore, training materials for pathologists and nephrologists may need to be developed to support optimal use of BanffNET.

BanffNET Indices are an extraction of relevant information from WSIs. To distinguish cases with similar histology but different underlying causes, ancillary testing will still be required. Similarly, BanffNET Indices are not meant to be used as a standalone prognostic test, since graft survival prognosis is best estimated with combined functional, immunological and histological data.

Our findings should be understood in the context of a number of limitations. First, metrics of clinical utility, while essential for safe and cost-effective clinical implementation, could not be obtained due to the lack of a gold standard against which to compare the BanffNET Indices, histological indices and Banff diagnoses. While a multi-reader study is currently the closest possible approximation to the histological ground truth, the number of biopsies in our multi-reader study was too small to measure calibration and clinical benefit with any meaningful degree of certainty. A large representative benchmark dataset would need to be constructed in order to objectively quantify and compare performance of kidney transplant WSI-based diagnostics. Second, patients’ treatment was adapted to their Banff diagnosis at the time of biopsy, which may differ from the Banff 2022 diagnoses which were re-derived from the original lesion scores for this study. Third, associations of BanffNET Indices, histological indices or Banff diagnoses with clinical outcomes may depend on the specific treatment decisions made in these cohorts. While data on treatment were not available, patients were treated according to the standard of care for transplant nephrology, and we deem it unlikely that common variations in treatment decisions would affect the conclusions. Fourth, no multiple testing adjustments were made and significance tests for differences in R² values should be interpreted as exploratory, since the family-wise error rate of 5% is likely not preserved given the large number of tests. Fifth, while BanffNET Indices performed well in the Dutch and Belgian cohorts, which were representative of the target population in these areas, further validation is needed in other European and non-European settings to extend geographic generalizability.

In this study, we developed and evaluated the BanffNET TCMR/TI Index, BanffNET AMR/MVI Index, BanffNET Activity Index and BanffNET Chronicity Index. These indices were based on BanffNET automated lesion scores and quantify TCMR-related and tubulointerstitial injury, AMR-related and microvascular injury, overall inflammatory activity and overall chronic injury in kidney transplant biopsies. These indices associate strongly with multi-observer aggregated diagnoses, single pathologist diagnoses, time to graft failure and molecular indices, suggesting high diagnostic value given the low cost per biopsy. To further validate the clinical utility of the indices, future studies should focus on implementation in the histopathology workflow.

## Supporting information

TRIPOD+AI checklist

## Online Methods

### LUMC Global Kidney Biopsy Reader Group

Adeyemi Adefidipe Abiola^1^, Sam Albadri^2^, Zahabia Ali^3^, Ahmed Altaleb^4^, Andrea Angioi^5^, Sultana G. Banu^6^, Marc Barry^7^, Ami R. Bhalodia^8^, Paola Bianco^9^, Verena Broecker^10^, Roman Bülow^11^, Bertrand Chauveau^12^, Guoli Chen^13^, Boonyarit Cheunsuchon^14^, Giovanna M. Crisi^15^, Simin Daneshvar^16^, Amelie Dendooven^17,18^, Pouneh Dokouhaki^19,20^, Cinthia B. Drachenberg^21^, Alton B. Farris^22^, Sophie Ferlicot^23^, Sandrine Florquin^24^, Francesco Fontana^25^, Jean-Baptiste Gibier^26^, Ian W. Gibson^27^, Saurabh Gujarathi^28^, Allen R. Hendricks^29^, Sufia Husain^30^, Jaynul Islam^31^, Wesam Ismail^32^, Geetha Jagannathan^22^, Jesper Kers^33,34,35,24^, Johannes Klager^35^, Nicolas Kozakowski^35^, Adriana Krizova^36^, Anila A. Kurien^37^, Boomi Kwon^38^, Vincenzo L’Imperio^39^, Felipe L. Ledesma^40^, Julia P. Low^41^, Joanne Martin^42^, Shweta S. Mehta^43^, Nidia Messias^44^, Salvatore E. Mignano^45^, Monik E. Miranda^46^, Gilbert Moeckel^47^, Chiara Pala^24,48^, Brendon Price^49^, Marc A. Ranson^27^, Andrea N. Rodriguez^50,51^, Joris J.T.H. Roelofs^24^, Avi Rosenberg^52^, Alireza Sadeghipour^53^, Miroslav Sekulic^54^, Suman Setty^55^, Michael Sheaff^3^, Maria F.S. Soares^56^, Jerasit Surintrspanont^57^, George Terinte-Balcan^58^, Francesca Testa^25^, David O. Henriquez Ticas^59^, Maria F. Toniolo^60^, Dominique van Midden^61^, Ramya K. Velagapudi^62^, Seethalakshmi Viswanathan^63^, Saskia von Stillfried^11^, Chih-Ying Wu^64,65^

^1^Department of Histopathology, African Medical Center of Excellence Hospital, Abuja, Nigeria

^2^Department of Pathology, Mayo Clinic, Mayo Clinic School of Medicine, Rochester, United States

^3^Department of Cellular Pathology, Barts NHS Trust, London, United Kingdom

^4^Histopathology Unit, Mubarak Alkabeer Hospital, Jabriya, Kuwait

^5^S.C.D.U. Nefrologia, Dialisi e Trapianto, ARNAS Brotzu Hospital, Cagliari, Italy

^6^Department of Pathology, Bangladesh Medical University, Dhaka, Bangladesh

^7^Department of Pathology, University of Utah, Salt Lake City, United States

^8^The Pathology Laboratory, Lake Charles, United States

^9^S.C. Anatomia Patologia, ARNAS Brotzu Hospital, Cagliari, Italy

^10^Department of Pathology, Sahlgrenska University Hospital, Gothenburg University, Gothenburg, Sweden

^11^Institute for Pathology, RWTH Aachen University Hospital, Aachen, Germany

^12^Department of Pathology, Bordeaux University Hospital, Bordeaux, France

^13^Department of Pathology, Penn State Health, Penn State College of Medicine, Hershey, United States

^14^Department of Pathology, Faculty of Medicine, Siriraj Hospital, Mahidol University, Bangkok, Thailand

^15^Department of Pathology, UMASS-Chan School of Medical School – Baystate, Springfield, United States

^16^Department of Anatomical Pathology, St. Vincent’s Hospital, University of Melbourne, Melbourne, Australia

^17^Department of Pathology, University Hospital of Ghent, Ghent University, Ghent, Belgium

^18^Department of Pathology, Faculty of Medicine and Health Sciences, University of Antwerp, Wilrijk, Belgium

^19^Department of Pathology and Laboratory Medicine, University of British Columbia, Vancouver, Canada

^20^Department of Pathology, St. Paul Hospital, Providence Health Care, Vancouver, Canada

^21^Department of Pathology, University of Maryland School of Medicine, Baltimore, United States

^22^Department of Pathology and laboratory Medicine, Emory University Hospital, Emory University, Atlanta, United States

^23^Department of Pathology, Le Kremlin-Bicêtre Hospital, Paris-Saclay University, Paris, France

^24^Department of Pathology, Amsterdam University Medical Center, University of Amsterdam, Amsterdam, Netherlands

^25^Nephrology, Dialysis and Kidney Transplant Unit, University Hospital of Modena, Modena, Italy

^26^Institute of Pathology, Centre Hospitalier Universitaire de Lille, Lille, France

^27^Department of Pathology, University of Manitoba, Winnipeg, Canada

^28^Department of Pathology, Manipal Hospital, Pune, India

^29^Department of Pathology, University of Texas Southwestern Medical Center, Dallas, United States

^30^Department of Pathology, College of Medicine, King Saud University Medical City, King Saud University, Riyadh, Saudi Arabia

^31^Department of Pathology, BIRDEM General Hospital, Dhaka, Bangladesh

^32^Department of Pathology, Beni-Suef University, Beni-Suef, Egypt

^33^Systems Pathology Research Group, Department of Pathology, Leiden University Medical Center, Leiden University, Leiden, Netherlands

^34^Leiden Transplant Center, Leiden University Medical Center, Leiden, Netherlands

^35^Department of Pathology and Clinical Bioinformatics, Erasmus Medical Center, Rotterdam, Netherlands

^35^Department of Pathology, General Hospital of Vienna, Medical University of Vienna, Vienna, Austria

^36^Department of Laboratory Medicine, St. Michael’s Hospital, University of Toronto, Toronto, Canada

^37^Department of Pathology, Renopath, Center for Renal and Urological Pathology, Chennai, India

^38^Department of Pathology, Mater Hospital, Brisbane, Australia

^39^Department of Medicine and Surgery, Pathology, Fondazione IRCCS San Gerardo dei Tintori, University of Milano-Bicocca, Monza, Italy

^40^Department of Pathology, University of São Paulo Medicine School, São Paulo, Brazil

^41^Department of Anatomical Pathology, Sydpath, St. Vincent’s Hospital, Sydney, University of South Wales, Australia

^42^Department of Cellular Pathology, The Royal London Hospital, Queen Mary University of London, London, United Kingdom

^43^Department of Pathology, Kauvery Hospital, Bengaluru, India

^44^Department of Pathology and Immunology, Washington University School of Medicine, Saint Louis, United States

^45^Department of Pathology & Genomic Medicine, Thomas Jefferson University Hospital, Philadelphia, United States

^46^Department of Anatomical Pathology, Cipto Mangunkusumo Hospital, Universitas Indonesia, Jakarta, Indonesia

^47^Department of Pathology, Yale New Haven Hospital, Yale University, New Haven, United States

^48^Department of Medicine and Surgery, Nephrology Unit, Parma University Hospital, Parma, Italy

^49^Division of Anatomical Pathology, Department of Pathology, NHLS Groote Schuur Hospital, University of Cape Town, Cape Town, South Africa

^50^Department of Pathology, Hospital Nacional Profesor Alejandro Posadas, Buenos Aires, Argentina

^51^Department of Pathology, The Favaloro Foundation University Hospital, Buenos Aires, Argentina

^52^Department of Pathology, Johns Hopkins University School of Medicine, Baltimore, United States

^53^Department of Pathology, Rasoul-Akram Hospital, Iran University of Medical Sciences, Tehran, Iran

^54^Department of Pathology and Cell Biology, Columbia University Medical Center, Columbia University, New York, United States

^55^Department of Pathology, University of Illinois Hospital and Health Sciences System, University of Illinois at Chicago, Chicago, United States

^56^Department of Cellular Pathology, John Radcliffe Hospital, Oxford University Hospitals NHS Foundation Trust, Oxford, United Kingdom

^57^Department of Pathology, King Chulalongkorn Memorial Hospital, The Thai Red Cross Society, Bangkok, Thailand

^58^Department of Pathology, Necker-Enfants Malades Hospital, Assistance Publique-Hôpitaux de Paris, Paris, France

^59^Department of Pathology, University Health Network, University of Toronto, Toronto, Canada

^60^Department of Pathology, Instituto de Trasplantes y Alta Complejidad, Buenos Aires, Argentina

^61^Department of Pathology, Radboud University Medical Center, Radboud University, Nijmegen, Netherlands

^62^Department of Pathology, University of Mississippi Medical Center, Jackson, United States

^63^Department of Tissue Pathology, Westmead Hospital, Westmead Clinical School, University of Sydney, Sydney, Australia

^64^Department of Pathology and laboratory Medicine, Taichung Veterans General Hospital, Taichung, Taiwan

^65^Institute of Biomedical Sciences, National Chung Hsing University, Taichung, Taiwan

### Study population and data collection

The training cohort consisted of kidney transplant biopsies taken at the University Medical Centers of Amsterdam, Leiden and Utrecht in the Netherlands between 2000 and 2024 and scored with the BanffNET algorithm. Biopsies consisted of indication biopsies conducted at the time of graft dysfunction and a minority of protocol biopsies conducted primarily at 6, 12 or 24 months post-transplantation in the context of clinical trials. The validation cohort consisted of kidney transplant biopsies taken at the University Hospitals Leuven, Belgium between March 2004 and March 2022 that were retrospectively scored with the BanffNET algorithm for the purpose of this study. Biopsies consisted of indication biopsies conducted at the time of graft dysfunction and protocol biopsies conducted primarily at 3, 12, and 24 months post-transplantation as part of an established follow-up protocol. The multi-reader study consisted of 36 biopsies scored by 67 pathologists and by BanffNET. Details are provided in Buzzanca et al.^12^ Biopsies of the training cohort and validation cohort were included if they were of adequate quality and taken between time of transplantation and time of graft failure or patient death. Biopsies from combined organ transplants or kidney transplants following non-renal organ transplants were excluded. The study was approved by the Research Ethics Committee of the University Hospitals Leuven (S64006), the institutional review board of Amsterdam University Medical Center (registration number 19.260), the institutional review board of University Medical Center Utrecht (registration number 19.482) and the institutional review board of Leiden University Medical Center (registration number W2020.031). Informed consent was waived for all patients. Data transfer agreements were set up between the contributing centers. No protocol was prepared for this retrospective study, and the study was not registered. There was no patient or public involvement in any aspect of the study.

### Kidney transplant biopsy evaluation

In all cohorts, biopsies were performed and scored as part of routine clinical practice by expert pathologists, following the Banff classification in force at the time of the biopsy. Banff diagnoses were subsequently re-derived from these lesion scores according to the Banff 2022 criteria, without rescoring the lesions. This included semi-quantitative grading of the lesions t, i, g, tma, ptc, v, ti, i-ifta, t-ifta, ci, ct, cg, cv, ah and mm. In the validation cohort, Banff diagnoses were derived from Banff lesion scores by expert pathologists. In the remaining cohorts, this conversion was done in an automated way, using the BanffClass tool.^13^ The Banff diagnoses of interest were Borderline Changes, TCMR, Probable AMR, AMR, MVI_DSA-/C4d-_ and BKPyVAN. For this study, isolated v lesions were not considered as TCMR, following recent evidence showing that in these cases, the presence of v-lesions did not significantly associate with graft survival.^14^ Further, MVI was defined as (g+ptc)≥2, with g≥1 if TCMR or Borderline Changes were present. AMR/MVI was defined as the presence of AMR or MVI_DSA-/C4d-_. Mixed Rejection was defined as TCMR in combination with either AMR or MVI_DSA-/C4d-_. Any Diagnosis was defined as Borderline Changes, TCMR, Probable AMR, AMR, MVI_DSA-/C4d-_, BKPyVAN, or any combination of these phenotypes. No Diagnosis was defined as the absence of Any Diagnosis. In addition, automated continuous lesion scores were retrospectively assigned with the BanffNET algorithm. These scores range from 0 to 1 and consist of the following: t, i, g, tma, ati, ptc, v, ti, i-ifta, t-ifta, ifta, cg, cv, ah, mm, gs, fsgs. BanffNET reports a single combined ifta score rather than separate ci/ct scores, and additionally scores ati, gs and fsgs, which are not part of routine semi-quantitative Banff grading at all centers. In addition, a subset of the validation cohort underwent transcriptomic analysis to assign recently proposed molecular indices based on the BHOT gene panel.^10^

### LUMC Global Reader Study

The LUMC Global Reader Study^5^ is a fully online digital reader study that included 36 cases (108 WSIs; an H&E, PAS and silver staining for each case), not overlapping with the LUMC training cohort. Slides were prospectively scanned with the Philips Ultrafast WSI scanner at a resolution of 0.25 micrometer per pixel for primary clinical diagnostics. Digitized WSIs were uploaded to the online platform Slidescore (https://www.slidescore.com) and fully anonymized. Included were a total of 67 pathologists from 50 cities in 24 countries: Argentina (2), Australia (4), Austria (3), Bangladesh (2), Belgium (1), Brazil (1), Canada (4), Egypt (1), France (4), Germany (2), India (3), Indonesia (1), Iran (1), Italy (6), Kuwait (1), Nigeria (1), Saudi Arabia (1), South Africa (1), Sweden (1), Taiwan (1), Thailand (2), The Netherlands (3), United Kingdom (4), United States of America (17). The study spans all permanently inhabited continents (Europe, Americas, Africa, Asia, Australia) and 13 of 22 (59%) geographical subregions as defined by the UNSD. All participants were renal pathologists experienced with scoring the Banff classification system with an average of 10 years (range 1 – 30 years) of experience. For each of the 36 cases, a single integrated electronic case report form (eCRF) was created in Slidescore and cases were presented to the pathologists in a per-person randomized order. Pathologists were asked to assess the following scores: total number of glomeruli, total number of globally sclerosed glomeruli, g (0-3), cg (0-3), mm (0-3), i (0-3), t (0-3), ptc (0-3), v (0-3 or absent), ti (0-100% with 10% increments), ifta (0-100% with 10% increments), i-ifta (0-3), t-ifta (0-3), cv (0-3 or absent), ah (0-3), acute tubular injury (present/absent), thrombotic microangiopathy (present/absent), and focal and segmental glomerulosclerosis (present/absent). HLA-DSA and C4d were available for these cases and were used to derive Banff diagnoses and histological indices.

### Development of BanffNET-based continuous indices

Four continuous indices were calculated, named the BanffNET TCMR/TI Index, BanffNET AMR/MVI Index, BanffNET Activity Index and BanffNET Chronicity Index. The indices were constructed as functions of BanffNET lesion scores and were standardized to have a mean of zero and a standard deviation of one in the training cohort.

For the TCMR/TI Index, the following BanffNET lesions were considered as candidate predictors: t, i, v, ptc, ati, ti, t-ifta, ifta, cv. For the AMR/MVI Index, the following BanffNET lesions were considered as candidate predictors: g, ptc, v, tma, cg, cv, ah, mm, fsgs. For the Activity Index, lesions representative of (exclusively) active injury were considered as candidate predictors: t, i, g, tma, ati, ptc, v. The lesions i-ifta, t-ifta and ti were not included because of their chronic component. For the Chronicity Index, lesions representative of chronic injury were considered as candidate predictors: ifta, cg, cv, ah, mm, gs, fsgs. The indices were computed for each individual using the following formula:

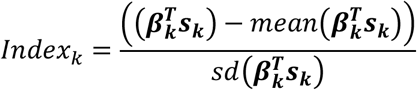

Here, *s_k_* is the vector of subsets of logit-transformed BanffNET lesion scores for BanffNET Index *k*, and 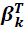 is the corresponding transposed vector of coefficients. For the TCMR/TI Index and AMR/MVI Index, these coefficients are derived from Ridge-penalized logistic regression models for Banff TCMR and Banff AMR/MVI respectively. For the Activity Index and Chronicity Index, the coefficients are derived from Ridge-penalized inverse-probability-of-censoring-weighted (IPCW) Cox models for the hazard of graft failure. Graft failure was defined as re-transplantation or return to dialysis. Cox models were censored for the competing event of death with a functioning graft.

Ridge penalization and subsequent linear recalibration on the training data were utilized to dampen extreme coefficients due to the strong multicollinearity of the BanffNET lesion scores, without compromising the internal calibration of the predictions. The penalization parameter was chosen such that all multivariable estimates had the same sign as the univariable estimates. The final subset of relevant lesion scores for each Index was obtained through backwards elimination. Specifically, lesion scores with a coefficient that contributed less than 6% of the sum of all coefficients were excluded in a stepwise manner. This produced model equations that balanced sparsity and clinical interpretability with internal predictive performance.

We deliberately chose not to train the Activity Index on the actual Any Diagnosis indicator, since training on the hazard of graft failure likely produces a score that more closely approximates the true, clinically relevant disease activity.

Clustering by center in the training cohort was not explicitly taken into account in development of the BanffNET Indices. Given the retrospective cohorts, post-hoc sample size calculations for model training and model evaluation were conducted.^15,16^ These calculations were based on the observed AUCs for Banff diagnoses, the observed prevalences of the target Banff diagnoses, the number of parameters in the BanffNET Indices development models, and a shrinkage factor of 0.90. Required training cohort sample sizes were 317, 323 and 1306 observations for prediction of TCMR, AMR/MVI and time to graft failure respectively. For model evaluation, calculated sample sizes were 4444, 3676 and 1826 for TCMR, MVI and Any Diagnosis respectively. For model evaluation, large required sample sizes were driven by the criterion for a narrow confidence interval for the calibration slope. However, the observed confidence intervals for the calibration slopes did in fact meet the criterion (narrower than 0.2), hence the validation was not underpowered.

## Missing data

An overview of all analyses with missing data strategy, number of observations and number of events is reported in **Supplementary Table 8**. For the analyses where multiple imputation was performed, missingness-at-random (MAR) was assumed. Specifically, most of the missingness occurred in HLA-DSA, C4d and molecular indices and depended on observed histology and calendar time. Missingness in graft survival (treated as instantaneous right censoring) depended primarily on calendar time. Multiple imputation by chained equations (MICE)^17^ was used with predictive mean matching for ordinal lesion scores and logistic regression for binary C4d and HLA-DSA. Banff diagnoses were derived in an automated way from imputed lesion scores, C4d and HLA-DSA using BanffClass, an online tool built for this purpose.^13^ 30 imputed datasets were generated. The models to train the coefficients of the BanffNET Indices were fitted on the concatenation of the 30 imputed datasets for simplicity and since only point estimates were needed, not accurate variances. In evaluation analyses, Rubin’s rule was applied if bootstrapping was not needed. If bootstrapping was applied, a two-stage resampling scheme was used where every bootstrap sample was drawn from a randomly chosen imputed dataset (each with equal probability). In the graft survival analyses, missing graft survival information was treated as censoring at time 0, which results in exclusion of those cases from the Cox models. Since censoring in the training cohort was not purely administrative, the assumption of non-informative censoring was likely violated. Hence, inverse probability of censoring weighting (IPCW) was applied to improve generalizability. Complete case analyses were performed for missingness in the histological indices in the validation cohort since the extent of the missingness was very limited and did not meaningfully change the representativeness of the cohort. The analyses involving BHOT molecular indices were also performed on validation cohort observations with complete histological data, but multiple imputation was used to impute the missingness in the BHOT molecular indices while taking into account the uncertainty in these imputations. Even though the degree of missingness of the molecular indices was very high, the MAR assumption is almost guaranteed to hold, since observation of these indices was the consequence of inclusion decisions which were explicitly based on the observed histology, with very low risk of selection bias as a result.

## Statistical analysis

### BanffNET Indices in relation to Banff diagnoses

In order to evaluate the TCMR/TI, AMR/MVI and Activity Indices, we first assessed their predictive performance for Banff TCMR, Banff MVI and Banff Any Diagnosis respectively. Areas under the receiver operating characteristic curve (AUC) were estimated for each pair of BanffNET Index and corresponding Banff diagnosis. Next, external calibration was evaluated on the validation cohort. Calibration intercepts and slopes were calculated with 95% confidence intervals using logistic regression models, as described in Van Calster and colleagues.^18^ Flexible calibration curves with point-wise 95% confidence intervals were fitted using generalized additive models with spline-based smooths and a logit link function.

Further, three thresholds were chosen for each Index, based on 5% false negatives, on Youden’s J and on 5% false positives in the training cohort. For each threshold, the sensitivity, specificity, negative predictive value (NPV), positive predictive value (PPV) and number of biopsies under the threshold were reported in both the training and the validation cohort. In addition, univariable logistic regression models were fitted to assess the strength and shape of the association of each Index with its corresponding Banff diagnosis. Linearity was explored with restricted cubic splines.

### BanffNET Indices in relation to robust multi-observer reference standards

Since Banff diagnoses are susceptible to inter-observer variability, a similar validation was done on the LUMC Global Reader Study^5^ dataset in which 67 pathologists each assigned lesion scores to the same set of 36 biopsies. Banff TCMR, MVI and Any Diagnosis were derived from the pathologist-assigned lesion scores and averaged across pathologists to obtain Banff multi-observer reference standards with continuous probability distributions ranging from 0 to 1. Analogously, pathologist-averaged histological indices were calculated. For example, the Banff TCMR multi-observer reference standard was calculated as follows:

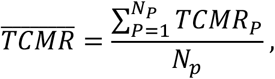

with *TCMR_P_* the TCMR diagnosis assigned by pathologist *P* and *N_P_* the total number of pathologists in the study. Next, R² values were calculated for each individual pathologist for each diagnosis or histological index from linear models with the reference standard as dependent variable and the corresponding Banff diagnosis, histological index, BanffNET Index or a combination of all three as independent variables. Reference standards from binary diagnoses were logit-transformed. R² values were then averaged across pathologists. For example, the explained variability in the Banff TCMR multi-observer reference standard by the average individual pathologist’s TCMR scoring is calculated as follows:

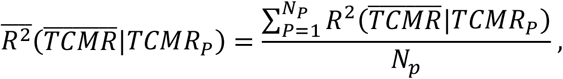

with *R*^2^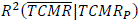 the variability in 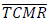 explained by *TCMR_P_*. The explained variability in each multi-observer reference standard by the corresponding BanffNET Indices was calculated from linear models without needing aggregation of R² values. R² values from the average individual pathologist’s diagnosis, the average individual pathologist’s histological index, the BanffNET Index and the combination of all three were all compared with each other. 95% bootstrap confidence intervals were calculated for the R² values and for the differences between them. The bias-correction and acceleration approach was applied to accommodate the boundedness and asymmetry of the R² and difference-in-R² bootstrap distributions.^19^ A difference was considered statistically significant if the confidence interval did not include zero. Loess-smoothed scatter plots were used to assess the shape of the association of each BanffNET Index with its corresponding multi-observer Banff reference standard.

### BanffNET Indices in relation to immunological factors

Associations of the BanffNET AMR/MVI Index and histological AMR/MVI index with HLA-DSA and C4d were compared in terms of the Nagelkerke R², derived from logistic regression models for HLA-DSA and C4d. 95% bootstrap confidence intervals were calculated using the bias-correction and acceleration approach.^19^

### BanffNET Indices in relation to kidney graft failure and molecular signatures

For the graft survival outcome, Royston-Sauerbrei R² was estimated from Cox models with time-dependent covariates, which enables the inclusion of multiple biopsies per patient without introducing bias, and reduces to regular Cox models in the absence of multiple biopsies per patient, which is the case in the training cohort. For molecular indices, regular R² was calculated from linear models. 95% bootstrap confidence intervals were calculated using the bias-correction and acceleration approach.^19^ Shapes of associations were explored with penalized smoothing splines for the graft survival outcome and with loess-smoothed scatter plots for the molecular indices.

All analyses were performed in R version 4.4.1 (R Foundation for Statistical Computing). The following packages were used: the MICE package^20^ for multiple imputation, glmnet^21^ for estimation of the BanffNET Indices weights, the survival package^22^ for survival analyses, pROC^23^ for estimation of ROCs and AUCs, boot^24^ for bootstrapping, mgcv^25^ for generalized additive models, and ggplot2^26^ and patchwork^27^ for plotting figures. Results were reported in accordance with the TRIPOD+AI recommendations, and the TRIPOD+AI checklist is available as online content linked to this article.^28^

## Data availability

The datasets analyzed in this study contain sensitive clinical information and cannot be made publicly available due to patient confidentiality. Validation data were obtained from participating centers under data transfer agreements. Deidentified data from the training and validation cohorts may be made available for academic research upon reasonable request to the corresponding author, subject to approval by the participating centers, institutional and ethical review, and completion of a data transfer agreement. For all requests, please contact the corresponding author, M.N., directly by email at.

## Code availability

The code for all analyses in this paper will be made available upon publication.

## Author contributions

AV, GB and MN designed the study and were responsible for study oversight; TQN, SF, DvdH, SM, JHV, SH, ADvZ, ASN, FJB, APJdV and JK were involved in training data collection, data curation and/or labeling; TV, PK, KW and MN curated and provided validation data; 67 pathologists scored the biopsies in the reader study; AV performed all analyses with input from GB, MN and JK; MN and JK were involved in supervision of the work; GV provided critical review of the statistical methods; AV and MN wrote the manuscript; AV, GB, MN, and JK provided critical review of the manuscript. All authors reviewed the content of the manuscript prior to submission.

## References

1. Naesens M, Roufosse C, Haas M, et al. The Banff 2022 Kidney Meeting Report: Reappraisal of microvascular inflammation and the role of biopsy-based transcript diagnostics. In: Am J Transplant. Vol 24. Elsevier B.V.; 2024:338–349. doi:10.1016/j.ajt.2023.10.016

2. Vickers AJ, Basch E, Kattan MW. Against Diagnosis. Ann Intern Med. 2008;149(3):200-203. doi:10.7326/0003-4819-149-3-200808050-00010

3. Furness PN, Taub N, Assmann KJM, et al. International Variation in Histologic Grading Is Large, and Persistent Feedback Does Not Improve Reproducibility. Am J Surg Pathol. 2003;27(6):805–810.

4. Becker JU, Mayerich D, Padmanabhan M, et al. Artificial intelligence and machine learning in nephropathology. Kidney Int. 2020;98(1):65–75. doi:10.1016/j.kint.2020.02.027

5. Hofstraat-Boersma R, du Long R, Buzzanca G, et al. Drivers of Diagnostic Variation in a Digital Global Kidney Transplant Reader Study. Preprint posted online July 13, 2026. doi:10.64898/2026.07.09.26357318

6. Vaulet T, Koshy P, Wellekens K, et al. Continuous indices to assess the phenotypic spectrum of kidney transplant rejection. Nat Commun. 2025;16(1):10417. doi:10.1038/s41467-025-65153-9

7. Haas M, Mirocha J, Huang E, et al. A Banff-based histologic chronicity index is associated with graft loss in patients with a kidney transplant and antibody-mediated rejection. Kidney Int. 2023;103(1):187–195. doi:10.1016/j.kint.2022.09.030

8. Halloran PF, Reeve J, Akalin E, et al. Real Time Central Assessment of Kidney Transplant Indication Biopsies by Microarrays: The INTERCOMEX Study. Am J Transplant. 2017;17(11):2851–2862. doi:10.1111/ajt.14329

9. Mengel M, Loupy A, Haas M, et al. Banff 2019 Meeting Report: Molecular diagnostics in solid organ transplantation–Consensus for the Banff Human Organ Transplant (B-HOT) gene panel and open source multicenter validation. Am J Transplant. 2020;20(9):2305–2317. doi:10.1111/ajt.16059

10. Naesens M, Wellekens K, Vaulet T, et al. Continuous Transcriptomic Indices for Assessing the Spectrum of Kidney Transplant Rejection: International Derivation and Validation Study. Preprint posted online March 12, 2026. doi:10.21203/rs.3.rs-9012525/v1

11. Hölscher DL, Bülow RD, Strauch M, Boor P. Advances in computational nephropathology. Kidney Int. 2025;108:1031–1044. doi:10.1016/j.kint.2025.06.029

12. Buzzanca G, Pala C, He J, et al. BanffNET, a Deep Learning System for Comprehensive Histological Lesion Quantification in Kidney Transplant Biopsies. Preprint posted online September 2, 2026. doi:10.64898/2026.08.28.26360029

13. Chauveau B, Naesens M. BanffClass – a web application for the classification of renal transplant biopsies. July 9, 2026. Accessed August 10, 2026. https://banffclass.anvil.app

14. Wellekens K, Coemans M, Koshy P, et al. The impact of the Banff v-lesion on rejection classification and outcomes: Insights from a multicenter study. American Journal of Transplantation. Published online 2025. doi:10.1016/j.ajt.2025.04.023

15. Riley RD, Ensor J, Snell KIE, et al. Calculating the sample size required for developing a clinical prediction model. BMJ. 2020;368:m441. doi:10.1136/bmj.m441

16. Riley RD, Snell KIE, Archer L, et al. Evaluation of clinical prediction models (part 3): calculating the sample size required for an external validation study. BMJ. 2024;384:e074821. doi:10.1136/bmj-2023-074821

17. van Buuren S. Flexible Imputation of Missing Data. Chapman & Hall/CRC; 2012.

18. Van Calster B, McLernon DJ, Van Smeden M, et al. Calibration: The Achilles heel of predictive analytics. BMC Med. 2019;17(1). doi:10.1186/s12916-019-1466-7

19. Efron B. Better Bootstrap Confidence Intervals. J Am Stat Assoc. 1987;82(397):171. doi:10.2307/2289144

20. van Buuren S, Groothuis-Oudshoorn K. mice: Multivariate Imputation by Chained Equations in R. J Stat Softw. 2011;45(3). doi:10.18637/jss.v045.i03

21. Friedman J, Hastie T, Tibshirani R. Regularization paths for generalized linear models via coordinate descent. J Stat Softw. 2010;33(1):1–22. doi:10.18637/jss.v033.i01

22. Therneau T. A Package for Survival Analysis in R. 2024.

23. Robin X, Turck N, Hainard A, et al. pROC: an open-source package for R and S+ to analyze and compare ROC curves. BMC Bioinformatics. 2011;12(1):77. doi:10.1186/1471-2105-12-77

24. Canty A, Ripley B. boot: Bootstrap Functions. CRAN: Contributed Packages. Preprint posted online April 8, 1999. doi:10.32614/CRAN.package.boot

25. Wood SN. Fast Stable Restricted Maximum Likelihood and Marginal Likelihood Estimation of Semiparametric Generalized Linear Models. J R Stat Soc Series B Stat Methodol. 2011;73(1):3–36. doi:10.1111/j.1467-9868.2010.00749.x

26. Wickham H. Ggplot2. Springer International Publishing; 2016. doi:10.1007/978-3-319-24277-4

27. Pedersen TL. patchwork: The Composer of Plots. CRAN: Contributed Packages. Preprint posted online December 1, 2019. doi:10.32614/CRAN.package.patchwork

28. Collins GS, Moons KGM, Dhiman P, et al. TRIPOD+AI statement: updated guidance for reporting clinical prediction models that use regression or machine learning methods. BMJ. 2024;385:e078378. doi:10.1136/bmj-2023-078378

