## Supplementary Material for "Continuous Diagnostic Indices from BanffNET Automated Lesion Scores for Kidney Transplant Pathology: Development and Evaluation of a Diagnostic Prediction Model"

**Supplementary tables**

**Supplementary Table 1. Transplant and biopsy characteristics of the full validation cohort and of the complete-case subset used in the analyses.**

| **Transplant characteristics** | **Full validation cohort (n = 1355 transplants)** | **Validation cohort complete-case subset (n = 1340 transplants)** |
|---|---|---|
| Male recipient sex, % | 64% | 63% |
| Recipient age, mean (SD) | 54 (13) | 54 (13) |
| Recipient BMI, mean (SD) | 26 (4) | 25 (4) |
| White European recipient ethnicity, % | 97% | 97% |
| Repeated transplantation, % | 15% | 15% |
| Pre-transplant HLA-DSA | 10% | 10% |
| Male donor sex, % | 53% | 53% |
| Donor age, mean (SD) | 49 (15) | 49 (15) |
| Deceased donor (brain death), % | 75% | 75% |
| Deceased donor (cardiac death), % | 18% | 18% |
| Living donor, % | 7% | 7% |
| CIT in hours, mean (SD) | 13 (6) | 13 (6) |
| Total number of HLA A/B/DR mismatches, median (IQR) | 3 (2 – 4) | 3 (2 – 4) |
| Missing, % | 30% | 30% |
| Total follow-up time in years, median (IQR) | 7.8 (4.0 – 11.4) | 7.8 (4.0 – 11.4) |
| Graft failure, % | 16% | 16% |
| TCMR at any time during follow-up, % | 20% | 20% |
| AMR at any time during follow-up, % | 11% | 11% |
| $MVI_{DSA-/C4d-}$ at any time during follow-up, % | 12% | 12% |

| Transplant characteristics | Full validation cohort (n = 1355 transplants) | Validation cohort complete-case subset (n = 1340 transplants) |
|---|---|---|
| BKPyVAN at any time during follow-up, % | 8% | 8% |
| DSA at any time during follow-up, % | 13% | 13% |
| **Biopsy characteristics** | **n = 4122 biopsies** | **n = 3863 biopsies** |
| Days from transplantation to biopsy, median (IQR) | 363 (89 – 742) | 363 (89 – 740) |
| Years from biopsy to end of follow-up, median (IQR) | 7.9 (4.0 – 11.0) | 7.9 (4.0 – 11.0) |
| Indication biopsy, % | 27% | 28% |
| HLA-DSA presence, % of non-missing | 13% | 13% |
| Missing, % | 0% | 0% |
| C4d (binary), % of non-missing | 5% | 5% |
| Missing, % | 1% | 0% |
| Banff 22 categories | | |
| No Diagnosis, % of non-missing | 71% | 70% |
| Missing, % | 0% | 0% |
| TCMR, % of non-missing | 9% | 9% |
| Missing, % | 0% | 0% |
| Borderline Changes, % of non-missing | 8% | 8% |
| Missing, % | 0% | 0% |
| AMR/MVI, % of non-missing | 13% | 13% |
| Missing, % | 0% | 0% |
| AMR, % of non-missing | 7% | 8% |
| Missing, % | 0% | 0% |
| Probable AMR, % of non-missing | 2% | 2% |
| Missing, % | 0% | 0% |
| MVI, % of non-missing | 11% | 12% |

| Biopsy characteristics | n = 4122 biopsies | n = 3863 biopsies |
|---|---|---|
| Missing, % | 0% | 0% |
| $MVI_{DSA-/C4d-}$, % of non-missing | 5% | 6% |
| Missing, % | 0% | 0% |
| BKPyVAN, % of non-missing | 3% | 3% |
| Missing, % | 0% | 0% |
| Mixed Rejection, % of non-missing | 3% | 3% |
| Missing, % | 0% | 0% |

Abbreviations: no., number; IQR, interquartile range; BMI, body mass index; HLA-DSA, anti-human leukocyte antigen donor specific antibodies; CIT, cold ischemia time; TCMR, T-cell mediated rejection; AMR, antibody-mediated rejection; $MVI_{DSA-/C4d-}$, C4d negative DSA negative microvascular inflammation; BKPyVAN, BK polyomavirus nephropathy. The complete-case subset comprises the biopsies with complete histological data that were used in all analyses, and is the validation cohort reported in Table 1.

**Supplementary Table 2. Differences in explained variability ($R^2$) of multi-observer aggregated diagnoses, by Banff diagnoses, histological indices, BanffNET Indices and the combination of all three in the multi-reader cohort (n = 36 biopsies scored by 67 pathologists).**

| Model 1 | Model 2 | $R^2$ difference | 95% CI |
|---|---|---|---|
| Combined TCMR/TI diagnostics | BanffNET TCMR/TI Index | 0.059 | (0.022; 0.117) |
| Combined TCMR/TI diagnostics | Histological TCMR/TI Index | 0.197 | (0.122; 0.279) |
| Combined TCMR/TI diagnostics | Banff TCMR diagnosis | 0.281 | (0.173; 0.369) |
| BanffNET TCMR/TI Index | Histological TCMR/TI Index | 0.138 | (0.023; 0.252) |
| BanffNET TCMR/TI Index | Banff TCMR diagnosis | 0.222 | (0.082; 0.328) |
| Histological TCMR/TI Index | Banff TCMR diagnosis | 0.084 | (0.004; 0.162) |
| Combined AMR/MVI diagnostics | BanffNET AMR/MVI Index | 0.131 | (0.068; 0.257) |
| Combined AMR/MVI diagnostics | Histological AMR/MVI Index | 0.179 | (0.080; 0.290) |
| Combined AMR/MVI diagnostics | Banff MVI diagnosis | 0.311 | (0.244; 0.373) |
| BanffNET AMR/MVI Index | Histological AMR/MVI Index | 0.049 | (-0.144; 0.198) |
| BanffNET AMR/MVI Index | Banff MVI diagnosis | 0.180 | (-0.012; 0.284) |
| Histological AMR/MVI Index | Banff MVI diagnosis | 0.132 | (0.028; 0.205) |
| Combined Activity diagnostics | BanffNET Activity Index | 0.144 | (0.066; 0.265) |
| Combined Activity diagnostics | Histological Activity Index | 0.136 | (0.071; 0.221) |
| Combined Activity diagnostics | Banff Any Diagnosis | 0.278 | (0.219; 0.329) |

| | | | |
|---|---|---|---|
| BanffNET Activity Index | Histological Activity Index | -0.008 | (-0.172; 0.136) |
| BanffNET Activity Index | Banff Any Diagnosis | 0.134 | (-0.033; 0.237) |
| Histological Activity Index | Banff Any Diagnosis | 0.142 | (0.068; 0.222) |

**Supplementary Table 3. Differences in explained variability ($R^2$) of multi-observer aggregated histological indices, by Banff diagnoses, histological indices, BanffNET Indices and the combination of all three in the multi-reader cohort (n = 36 biopsies scored by 67 pathologists).**

| **Model 1** | **Model 2** | **$R^2$ difference** | **95% CI** |
|---|---|---|---|
| Combined TCMR/TI diagnostics | BanffNET TCMR/TI Index | 0.110 | (0.052; 0.189) |
| Combined TCMR/TI diagnostics | Histological TCMR/TI Index | 0.121 | (0.064; 0.179) |
| Combined TCMR/TI diagnostics | Banff TCMR diagnosis | 0.286 | (0.181; 0.372) |
| BanffNET TCMR/TI Index | Histological TCMR/TI Index | 0.011 | (-0.121; 0.125) |
| BanffNET TCMR/TI Index | Banff TCMR diagnosis | 0.176 | (0.020; 0.293) |
| Histological TCMR/TI Index | Banff TCMR diagnosis | 0.165 | (0.101; 0.236) |
| Combined AMR/MVI diagnostics | BanffNET AMR/MVI Index | 0.344 | (0.175; 0.568) |
| Combined AMR/MVI diagnostics | Histological AMR/MVI Index | 0.055 | (0.032; 0.081) |
| Combined AMR/MVI diagnostics | Banff MVI diagnosis | 0.224 | (0.157; 0.283) |
| BanffNET AMR/MVI Index | Histological AMR/MVI Index | -0.289 | (-0.514; -0.108) |
| BanffNET AMR/MVI Index | Banff MVI diagnosis | -0.120 | (-0.366; 0.078) |
| Histological AMR/MVI Index | Banff MVI diagnosis | 0.169 | (0.121; 0.225) |
| Combined Activity diagnostics | BanffNET Activity Index | 0.201 | (0.121; 0.351) |
| Combined Activity diagnostics | Histological Activity Index | 0.083 | (0.051; 0.109) |
| Combined Activity diagnostics | Banff Any Diagnosis | 0.368 | (0.293; 0.447) |
| BanffNET Activity Index | Histological Activity Index | -0.118 | (-0.287; -0.021) |
| BanffNET Activity Index | Banff Any Diagnosis | 0.167 | (0.009; 0.277) |
| Histological Activity Index | Banff Any Diagnosis | 0.285 | (0.228; 0.365) |
| Combined Chronicity diagnostics | BanffNET Chronicity Index | 0.103 | (0.048; 0.192) |
| Combined Chronicity diagnostics | Histological Chronicity Index | 0.182 | (0.118; 0.258) |
| BanffNET Chronicity Index | Histological Chronicity Index | 0.079 | (-0.061; 0.201) |

**Supplementary Table 4. Differences in explained variability ($R^2$) of C4d deposition and HLA-DSA presence, by Banff AMR diagnosis, histological AMR/MVI index and BanffNET AMR/MVI Index in the training and validation cohort (training n = 2544 biopsies, validation n = 3863 biopsies).**

| Cohort | Outcome | Variable 1 | Variable 1 $R^2$ | Variable 2 | Variable 2 $R^2$ | $R^2$ difference | 95% CI |
|---|---|---|---|---|---|---|---|
| Training | C4d | BanffNET AMR/MVI Index | 0.168 | Histological AMR/MVI Index | 0.611 | -0.443 | (-0.510; -0.368) |
| Training | C4d | BanffNET AMR/MVI Index | 0.168 | Banff AMR diagnosis | 0.289 | -0.121 | (-0.197; -0.041) |
| Training | C4d | Histological AMR/MVI Index | 0.611 | Banff AMR diagnosis | 0.289 | 0.322 | (0.250; 0.392) |
| Training | HLA-DSA | BanffNET AMR/MVI Index | 0.168 | Histological AMR/MVI Index | 0.207 | -0.040 | (-0.110; 0.035) |
| Training | HLA-DSA | BanffNET AMR/MVI Index | 0.168 | Banff AMR diagnosis | 0.159 | 0.009 | (-0.058; 0.078) |
| Training | HLA-DSA | Histological AMR/MVI Index | 0.207 | Banff AMR diagnosis | 0.159 | 0.048 | (-0.021; 0.116) |
| Validation | C4d | BanffNET AMR/MVI Index | 0.100 | Histological AMR/MVI Index | 0.619 | -0.519 | (-0.573; -0.461) |
| Validation | C4d | BanffNET AMR/MVI Index | 0.100 | Banff AMR diagnosis | 0.249 | -0.149 | (-0.212; -0.092) |
| Validation | C4d | Histological AMR/MVI Index | 0.619 | Banff AMR diagnosis | 0.249 | 0.370 | (0.300; 0.435) |
| Validation | HLA-DSA | BanffNET AMR/MVI Index | 0.106 | Histological AMR/MVI Index | 0.225 | -0.119 | (-0.165; -0.075) |
| Validation | HLA-DSA | BanffNET AMR/MVI Index | 0.106 | Banff AMR diagnosis | 0.189 | -0.082 | (-0.129; -0.039) |
| Validation | HLA-DSA | Histological AMR/MVI Index | 0.225 | Banff AMR diagnosis | 0.189 | 0.037 | (-0.014; 0.091) |

**Supplementary Table 5. Differences in explained variability ($R^2$) of graft survival by Banff diagnoses, histological indices, BanffNET Indices and the combination of the three in the training cohort and within mutually exclusive Banff categories (n = 1874).**

| Category | Median N (min – max) | Model 1 | Pseudo-$R^2$ | Model 2 | Pseudo-$R^2$ | $R^2$ difference | 95% CI |
|---|---|---|---|---|---|---|---|
| All biopsies | 1874 | Combined TCMR/TI diagnostics | 0.166 | BanffNET TCMR/TI Index | 0.155 | 0.010 | (-0.005; 0.029) |
| All biopsies | 1874 | Combined TCMR/TI diagnostics | 0.166 | Histological TCMR/TI Index | 0.112 | 0.054 | (0.018; 0.092) |
| All biopsies | 1874 | Combined TCMR/TI diagnostics | 0.166 | Banff TCMR | 0.077 | 0.089 | (0.044; 0.140) |
| All biopsies | 1874 | BanffNET TCMR/TI Index | 0.155 | Histological TCMR/TI Index | 0.112 | 0.044 | (0.020; 0.072) |
| All biopsies | 1874 | BanffNET TCMR/TI Index | 0.155 | Banff TCMR | 0.077 | 0.079 | (0.043; 0.116) |
| All biopsies | 1874 | Histological TCMR/TI Index | 0.112 | Banff TCMR | 0.077 | 0.035 | (0.003; 0.066) |
| All biopsies | 1874 | Combined AMR/MVI diagnostics | 0.215 | BanffNET AMR/MVI Index | 0.225 | -0.010 | (-0.030; 0.005) |
| All biopsies | 1874 | Combined AMR/MVI diagnostics | 0.215 | Histological AMR/MVI Index | 0.123 | 0.092 | (0.048; 0.141) |
| All biopsies | 1874 | Combined AMR/MVI diagnostics | 0.215 | Banff AMR/MVI | 0.138 | 0.076 | (0.024; 0.128) |
| All biopsies | 1874 | BanffNET AMR/MVI Index | 0.225 | Histological AMR/MVI Index | 0.123 | 0.102 | (0.062; 0.137) |
| All biopsies | 1874 | BanffNET AMR/MVI Index | 0.225 | Banff AMR/MVI | 0.138 | 0.086 | (0.038; 0.129) |
| All biopsies | 1874 | Histological AMR/MVI Index | 0.123 | Banff AMR/MVI | 0.138 | -0.016 | (-0.049; 0.017) |
| All biopsies | 1874 | Combined Activity diagnostics | 0.198 | BanffNET Activity Index | 0.195 | 0.003 | (-0.016; 0.025) |
| All biopsies | 1874 | Combined Activity diagnostics | 0.198 | Histological Activity Index | 0.111 | 0.087 | (0.041; 0.140) |
| All biopsies | 1874 | Combined Activity diagnostics | 0.198 | Banff Any Diagnosis | 0.113 | 0.085 | (0.026; 0.149) |
| All biopsies | 1874 | BanffNET Activity Index | 0.195 | Histological Activity Index | 0.111 | 0.084 | (0.052; 0.124) |
| All biopsies | 1874 | BanffNET Activity Index | 0.195 | Banff Any Diagnosis | 0.113 | 0.082 | (0.029; 0.130) |
| All biopsies | 1874 | Histological Activity Index | 0.111 | Banff Any Diagnosis | 0.113 | -0.002 | (-0.041; 0.034) |
| All biopsies | 1874 | Combined Chronicity diagnostics | 0.317 | BanffNET Chronicity Index | 0.311 | 0.006 | (0.000; 0.020) |
| All biopsies | 1874 | Combined Chronicity diagnostics | 0.317 | Histological Chronicity Index | 0.254 | 0.062 | (0.029; 0.102) |
| All biopsies | 1874 | BanffNET Chronicity Index | 0.311 | Histological Chronicity Index | 0.254 | 0.056 | (0.016; 0.101) |
| Exclusive TCMR | 191 (184 – 194) | Combined TCMR/TI diagnostics | 0.071 | BanffNET TCMR/TI Index | 0.062 | 0.009 | (-0.019; 0.079) |

| | | | | | | | |
|---|---|---|---|---|---|---|---|
| Exclusive TCMR | 191 (184 – 194) | Combined TCMR/TI diagnostics | 0.071 | Histological TCMR/TI Index | 0.021 | 0.050 | (-0.018; 0.185) |
| Exclusive TCMR | 191 (184 – 194) | BanffNET TCMR/TI Index | 0.062 | Histological TCMR/TI Index | 0.021 | 0.041 | (-0.018; 0.148) |
| Exclusive TCMR | 191 (184 – 194) | Combined AMR/MVI diagnostics | 0.082 | BanffNET AMR/MVI Index | 0.003 | 0.078 | (0.005; 0.213) |
| Exclusive TCMR | 191 (184 – 194) | Combined AMR/MVI diagnostics | 0.082 | Histological AMR/MVI Index | 0.099 | -0.017 | (-0.078; 0.016) |
| Exclusive TCMR | 191 (184 – 194) | BanffNET AMR/MVI Index | 0.003 | Histological AMR/MVI Index | 0.099 | -0.095 | (-0.270; 0.000) |
| Exclusive TCMR | 191 (184 – 194) | Combined Activity diagnostics | 0.062 | BanffNET Activity Index | 0.014 | 0.048 | (0.000; 0.176) |
| Exclusive TCMR | 191 (184 – 194) | Combined Activity diagnostics | 0.062 | Histological Activity Index | 0.064 | -0.002 | (-0.031; 0.025) |
| Exclusive TCMR | 191 (184 – 194) | BanffNET Activity Index | 0.014 | Histological Activity Index | 0.064 | -0.050 | (-0.164; 0.009) |
| Exclusive TCMR | 191 (184 – 194) | Combined Chronicity diagnostics | 0.189 | BanffNET Chronicity Index | 0.173 | 0.016 | (-0.019; 0.110) |
| Exclusive TCMR | 191 (184 – 194) | Combined Chronicity diagnostics | 0.189 | Histological Chronicity Index | 0.175 | 0.014 | (-0.025; 0.107) |
| Exclusive TCMR | 191 (184 – 194) | BanffNET Chronicity Index | 0.173 | Histological Chronicity Index | 0.175 | -0.002 | (-0.098; 0.112) |
| Exclusive AMR | 146 (141 – 154) | Combined TCMR/TI diagnostics | 0.152 | BanffNET TCMR/TI Index | 0.142 | 0.010 | (-0.026; 0.115) |
| Exclusive AMR | 146 (141 – 154) | Combined TCMR/TI diagnostics | 0.152 | Histological TCMR/TI Index | 0.098 | 0.054 | (-0.024; 0.194) |
| Exclusive AMR | 146 (141 – 154) | BanffNET TCMR/TI Index | 0.142 | Histological TCMR/TI Index | 0.098 | 0.044 | (-0.073; 0.181) |
| Exclusive AMR | 146 (141 – 154) | Combined AMR/MVI diagnostics | 0.120 | BanffNET AMR/MVI Index | 0.098 | 0.022 | (-0.029; 0.140) |
| Exclusive AMR | 146 (141 – 154) | Combined AMR/MVI diagnostics | 0.120 | Histological AMR/MVI Index | 0.003 | 0.117 | (0.013; 0.248) |
| Exclusive AMR | 146 (141 – 154) | BanffNET AMR/MVI Index | 0.098 | Histological AMR/MVI Index | 0.003 | 0.095 | (-0.004; 0.200) |
| Exclusive AMR | 146 (141 – 154) | Combined Activity diagnostics | 0.110 | BanffNET Activity Index | 0.089 | 0.021 | (-0.049; 0.126) |
| Exclusive AMR | 146 (141 – 154) | Combined Activity diagnostics | 0.110 | Histological Activity Index | 0.009 | 0.101 | (-0.004; 0.276) |
| Exclusive AMR | 146 (141 – 154) | BanffNET Activity Index | 0.089 | Histological Activity Index | 0.009 | 0.080 | (-0.003; 0.250) |
| Exclusive AMR | 146 (141 – 154) | Combined Chronicity diagnostics | 0.286 | BanffNET Chronicity Index | 0.267 | 0.019 | (-0.011; 0.132) |
| Exclusive AMR | 146 (141 – 154) | Combined Chronicity diagnostics | 0.286 | Histological Chronicity Index | 0.221 | 0.065 | (-0.008; 0.192) |
| Exclusive AMR | 146 (141 – 154) | BanffNET Chronicity Index | 0.267 | Histological Chronicity Index | 0.221 | 0.046 | (-0.102; 0.180) |
| Exclusive Borderline Changes | 182 (180 – 184) | Combined TCMR/TI diagnostics | 0.123 | BanffNET TCMR/TI Index | 0.116 | 0.007 | (-0.031; 0.074) |
| Exclusive Borderline Changes | 182 (180 – 184) | Combined TCMR/TI diagnostics | 0.123 | Histological TCMR/TI Index | 0.069 | 0.054 | (-0.038; 0.244) |
| Exclusive Borderline Changes | 182 (180 – 184) | BanffNET TCMR/TI Index | 0.116 | Histological TCMR/TI Index | 0.069 | 0.047 | (-0.087; 0.202) |
| Exclusive Borderline Changes | 182 (180 – 184) | Combined AMR/MVI diagnostics | 0.070 | BanffNET AMR/MVI Index | 0.062 | 0.008 | (-0.015; 0.096) |

| | | | | | | | |
|---|---|---|---|---|---|---|---|
| Exclusive Borderline Changes | 182 (180 – 184) | Combined AMR/MVI diagnostics | 0.070 | Histological AMR/MVI Index | 0.031 | 0.039 | (-0.032; 0.232) |
| Exclusive Borderline Changes | 182 (180 – 184) | BanffNET AMR/MVI Index | 0.062 | Histological AMR/MVI Index | 0.031 | 0.031 | (-0.068; 0.233) |
| Exclusive Borderline Changes | 182 (180 – 184) | Combined Activity diagnostics | 0.057 | BanffNET Activity Index | 0.050 | 0.007 | (-0.047; 0.101) |
| Exclusive Borderline Changes | 182 (180 – 184) | Combined Activity diagnostics | 0.057 | Histological Activity Index | 0.042 | 0.015 | (-0.041; 0.091) |
| Exclusive Borderline Changes | 182 (180 – 184) | BanffNET Activity Index | 0.050 | Histological Activity Index | 0.042 | 0.008 | (-0.102; 0.151) |
| Exclusive Borderline Changes | 182 (180 – 184) | Combined Chronicity diagnostics | 0.314 | BanffNET Chronicity Index | 0.300 | 0.013 | (-0.032; 0.105) |
| Exclusive Borderline Changes | 182 (180 – 184) | Combined Chronicity diagnostics | 0.314 | Histological Chronicity Index | 0.198 | 0.116 | (-0.027; 0.317) |
| Exclusive Borderline Changes | 182 (180 – 184) | BanffNET Chronicity Index | 0.300 | Histological Chronicity Index | 0.198 | 0.103 | (-0.049; 0.256) |
| Exclusive $MVI_{DSA-/C4d-}$ | 93 (85 – 99) | Combined TCMR/TI diagnostics | 0.088 | BanffNET TCMR/TI Index | 0.066 | 0.022 | (-0.064; 0.214) |
| Exclusive $MVI_{DSA-/C4d-}$ | 93 (85 – 99) | Combined TCMR/TI diagnostics | 0.088 | Histological TCMR/TI Index | 0.008 | 0.080 | (-0.022; 0.324) |
| Exclusive $MVI_{DSA-/C4d-}$ | 93 (85 – 99) | BanffNET TCMR/TI Index | 0.066 | Histological TCMR/TI Index | 0.008 | 0.058 | (-0.034; 0.316) |
| Exclusive $MVI_{DSA-/C4d-}$ | 93 (85 – 99) | Combined AMR/MVI diagnostics | 0.331 | BanffNET AMR/MVI Index | 0.324 | 0.008 | (-0.063; 0.123) |
| Exclusive $MVI_{DSA-/C4d-}$ | 93 (85 – 99) | Combined AMR/MVI diagnostics | 0.331 | Histological AMR/MVI Index | 0.204 | 0.127 | (-0.009; 0.441) |
| Exclusive $MVI_{DSA-/C4d-}$ | 93 (85 – 99) | BanffNET AMR/MVI Index | 0.324 | Histological AMR/MVI Index | 0.204 | 0.120 | (-0.074; 0.423) |
| Exclusive $MVI_{DSA-/C4d-}$ | 93 (85 – 99) | Combined Activity diagnostics | 0.192 | BanffNET Activity Index | 0.179 | 0.013 | (-0.051; 0.116) |
| Exclusive $MVI_{DSA-/C4d-}$ | 93 (85 – 99) | Combined Activity diagnostics | 0.192 | Histological Activity Index | 0.037 | 0.154 | (-0.004; 0.445) |
| Exclusive $MVI_{DSA-/C4d-}$ | 93 (85 – 99) | BanffNET Activity Index | 0.179 | Histological Activity Index | 0.037 | 0.141 | (-0.024; 0.441) |
| Exclusive $MVI_{DSA-/C4d-}$ | 93 (85 – 99) | Combined Chronicity diagnostics | 0.277 | BanffNET Chronicity Index | 0.275 | 0.002 | (-0.072; 0.155) |
| Exclusive $MVI_{DSA-/C4d-}$ | 93 (85 – 99) | Combined Chronicity diagnostics | 0.277 | Histological Chronicity Index | 0.196 | 0.081 | (-0.047; 0.267) |
| Exclusive $MVI_{DSA-/C4d-}$ | 93 (85 – 99) | BanffNET Chronicity Index | 0.275 | Histological Chronicity Index | 0.196 | 0.079 | (-0.146; 0.282) |
| Mixed Rejection | 187 (185 – 192) | Combined TCMR/TI diagnostics | 0.093 | BanffNET TCMR/TI Index | 0.074 | 0.019 | (-0.011; 0.090) |
| Mixed Rejection | 187 (185 – 192) | Combined TCMR/TI diagnostics | 0.093 | Histological TCMR/TI Index | 0.086 | 0.007 | (-0.014; 0.076) |
| Mixed Rejection | 187 (185 – 192) | BanffNET TCMR/TI Index | 0.074 | Histological TCMR/TI Index | 0.086 | -0.012 | (-0.091; 0.075) |
| Mixed Rejection | 187 (185 – 192) | Combined AMR/MVI diagnostics | 0.029 | BanffNET AMR/MVI Index | 0.031 | -0.002 | (-0.042; 0.034) |
| Mixed Rejection | 187 (185 – 192) | Combined AMR/MVI diagnostics | 0.029 | Histological AMR/MVI Index | 0.001 | 0.028 | (-0.001; 0.157) |
| Mixed Rejection | 187 (185 – 192) | BanffNET AMR/MVI Index | 0.031 | Histological AMR/MVI Index | 0.001 | 0.030 | (-0.014; 0.159) |
| Mixed Rejection | 187 (185 – 192) | Combined Activity diagnostics | 0.062 | BanffNET Activity Index | 0.066 | -0.004 | (-0.042; 0.016) |

| | | | | | | | |
|---|---|---|---|---|---|---|---|
| Mixed Rejection | 187 (185 – 192) | Combined Activity diagnostics | 0.062 | Histological Activity Index | 0.034 | 0.028 | (-0.010; 0.130) |
| Mixed Rejection | 187 (185 – 192) | BanffNET Activity Index | 0.066 | Histological Activity Index | 0.034 | 0.032 | (-0.040; 0.158) |
| Mixed Rejection | 187 (185 – 192) | Combined Chronicity diagnostics | 0.199 | BanffNET Chronicity Index | 0.179 | 0.021 | (-0.010; 0.109) |
| Mixed Rejection | 187 (185 – 192) | Combined Chronicity diagnostics | 0.199 | Histological Chronicity Index | 0.164 | 0.035 | (-0.015; 0.143) |
| Mixed Rejection | 187 (185 – 192) | BanffNET Chronicity Index | 0.179 | Histological Chronicity Index | 0.164 | 0.015 | (-0.073; 0.113) |
| No Diagnosis | 904 (897 – 909) | Combined TCMR/TI diagnostics | 0.184 | BanffNET TCMR/TI Index | 0.182 | 0.003 | (-0.004; 0.021) |
| No Diagnosis | 904 (897 – 909) | Combined TCMR/TI diagnostics | 0.184 | Histological TCMR/TI Index | 0.104 | 0.081 | (0.015; 0.162) |
| No Diagnosis | 904 (897 – 909) | BanffNET TCMR/TI Index | 0.182 | Histological TCMR/TI Index | 0.104 | 0.078 | (0.014; 0.153) |
| No Diagnosis | 904 (897 – 909) | Combined AMR/MVI diagnostics | 0.275 | BanffNET AMR/MVI Index | 0.255 | 0.020 | (-0.001; 0.060) |
| No Diagnosis | 904 (897 – 909) | Combined AMR/MVI diagnostics | 0.275 | Histological AMR/MVI Index | 0.007 | 0.268 | (0.179; 0.377) |
| No Diagnosis | 904 (897 – 909) | BanffNET AMR/MVI Index | 0.255 | Histological AMR/MVI Index | 0.007 | 0.248 | (0.165; 0.355) |
| No Diagnosis | 904 (897 – 909) | Combined Activity diagnostics | 0.211 | BanffNET Activity Index | 0.189 | 0.021 | (-0.007; 0.072) |
| No Diagnosis | 904 (897 – 909) | Combined Activity diagnostics | 0.211 | Histological Activity Index | 0.001 | 0.209 | (0.123; 0.318) |
| No Diagnosis | 904 (897 – 909) | BanffNET Activity Index | 0.189 | Histological Activity Index | 0.001 | 0.188 | (0.097; 0.292) |
| No Diagnosis | 904 (897 – 909) | Combined Chronicity diagnostics | 0.338 | BanffNET Chronicity Index | 0.339 | -0.002 | (-0.025; 0.005) |
| No Diagnosis | 904 (897 – 909) | Combined Chronicity diagnostics | 0.338 | Histological Chronicity Index | 0.230 | 0.107 | (0.053; 0.195) |
| No Diagnosis | 904 (897 – 909) | BanffNET Chronicity Index | 0.339 | Histological Chronicity Index | 0.230 | 0.109 | (0.046; 0.191) |

The sample size of the diagnostic subgroups varied per imputed dataset and was reported as median with minimum and maximum. The categories Exclusive BKPyVAN and Exclusive Probable AMR had insufficient events for survival analysis.

**Supplementary Table 6. Differences in explained variability ($R^2$) of graft survival by Banff diagnoses, histological indices, BanffNET Indices and the combination of the three in the validation cohort and within mutually exclusive Banff categories (n = 3863).**

| Category | N | Model 1 | Pseudo-$R^2$ | Model 2 | Pseudo-$R^2$ | $R^2$ difference | 95% CI |
|---|---|---|---|---|---|---|---|
| All biopsies | 3863 | Combined TCMR/TI diagnostics | 0.280 | BanffNET TCMR/TI Index | 0.263 | 0.018 | (0.000; 0.047) |
| All biopsies | 3863 | Combined TCMR/TI diagnostics | 0.280 | Histological TCMR/TI Index | 0.242 | 0.038 | (0.000; 0.093) |
| All biopsies | 3863 | Combined TCMR/TI diagnostics | 0.280 | Banff TCMR | 0.193 | 0.087 | (0.001; 0.177) |
| All biopsies | 3863 | BanffNET TCMR/TI Index | 0.263 | Histological TCMR/TI Index | 0.242 | 0.020 | (-0.044; 0.089) |
| All biopsies | 3863 | BanffNET TCMR/TI Index | 0.263 | Banff TCMR | 0.193 | 0.070 | (-0.027; 0.168) |
| All biopsies | 3863 | Histological TCMR/TI Index | 0.242 | Banff TCMR | 0.193 | 0.049 | (-0.029; 0.132) |
| All biopsies | 3863 | Combined AMR/MVI diagnostics | 0.339 | BanffNET AMR/MVI Index | 0.331 | 0.007 | (-0.010; 0.030) |
| All biopsies | 3863 | Combined AMR/MVI diagnostics | 0.339 | Histological AMR/MVI Index | 0.209 | 0.129 | (0.065; 0.193) |
| All biopsies | 3863 | Combined AMR/MVI diagnostics | 0.339 | Banff AMR/MVI | 0.224 | 0.114 | (0.040; 0.194) |
| All biopsies | 3863 | BanffNET AMR/MVI Index | 0.331 | Histological AMR/MVI Index | 0.209 | 0.122 | (0.043; 0.201) |
| All biopsies | 3863 | BanffNET AMR/MVI Index | 0.331 | Banff AMR/MVI | 0.224 | 0.107 | (0.017; 0.198) |
| All biopsies | 3863 | Histological AMR/MVI Index | 0.209 | Banff AMR/MVI | 0.224 | -0.015 | (-0.070; 0.050) |
| All biopsies | 3863 | Combined Activity diagnostics | 0.305 | BanffNET Activity Index | 0.304 | 0.001 | (-0.011; 0.020) |
| All biopsies | 3863 | Combined Activity diagnostics | 0.305 | Histological Activity Index | 0.207 | 0.099 | (0.042; 0.164) |
| All biopsies | 3863 | Combined Activity diagnostics | 0.305 | Banff Any Diagnosis | 0.204 | 0.101 | (0.035; 0.181) |
| All biopsies | 3863 | BanffNET Activity Index | 0.304 | Histological Activity Index | 0.207 | 0.098 | (0.031; 0.168) |
| All biopsies | 3863 | BanffNET Activity Index | 0.304 | Banff Any Diagnosis | 0.204 | 0.100 | (0.022; 0.186) |
| All biopsies | 3863 | Histological Activity Index | 0.207 | Banff Any Diagnosis | 0.204 | 0.002 | (-0.059; 0.063) |
| All biopsies | 3863 | Combined Chronicity diagnostics | 0.437 | BanffNET Chronicity Index | 0.391 | 0.046 | (0.020; 0.099) |
| All biopsies | 3863 | Combined Chronicity diagnostics | 0.437 | Histological Chronicity Index | 0.350 | 0.087 | (0.045; 0.134) |
| All biopsies | 3863 | BanffNET Chronicity Index | 0.391 | Histological Chronicity Index | 0.350 | 0.041 | (-0.043; 0.110) |

| | | | | | | | |
|---|---|---|---|---|---|---|---|
| Exclusive TCMR | 219 | Combined TCMR/TI diagnostics | 0.105 | BanffNET TCMR/TI Index | 0.070 | 0.035 | (-0.033; 0.311) |
| Exclusive TCMR | 219 | Combined TCMR/TI diagnostics | 0.105 | Histological TCMR/TI Index | 0.138 | -0.033 | (-0.104; -0.013) |
| Exclusive TCMR | 219 | BanffNET TCMR/TI Index | 0.070 | Histological TCMR/TI Index | 0.138 | -0.067 | (-0.392; 0.113) |
| Exclusive TCMR | 219 | Combined AMR/MVI diagnostics | 0.018 | BanffNET AMR/MVI Index | 0.015 | 0.003 | (-0.052; 0.071) |
| Exclusive TCMR | 219 | Combined AMR/MVI diagnostics | 0.018 | Histological AMR/MVI Index | 0.000 | 0.017 | (-0.158; 0.195) |
| Exclusive TCMR | 219 | BanffNET AMR/MVI Index | 0.015 | Histological AMR/MVI Index | 0.000 | 0.015 | (-0.075; 0.245) |
| Exclusive TCMR | 219 | Combined Activity diagnostics | 0.084 | BanffNET Activity Index | 0.056 | 0.028 | (-0.022; 0.204) |
| Exclusive TCMR | 219 | Combined Activity diagnostics | 0.084 | Histological Activity Index | 0.083 | 0.002 | (-0.045; 0.077) |
| Exclusive TCMR | 219 | BanffNET Activity Index | 0.056 | Histological Activity Index | 0.083 | -0.027 | (-0.234; 0.117) |
| Exclusive TCMR | 219 | Combined Chronicity diagnostics | 0.356 | BanffNET Chronicity Index | 0.284 | 0.072 | (-0.027; 0.299) |
| Exclusive TCMR | 219 | Combined Chronicity diagnostics | 0.356 | Histological Chronicity Index | 0.322 | 0.034 | (-0.048; 0.218) |
| Exclusive TCMR | 219 | BanffNET Chronicity Index | 0.284 | Histological Chronicity Index | 0.322 | -0.038 | (-0.309; 0.246) |
| Exclusive AMR | 180 | Combined TCMR/TI diagnostics | 0.101 | BanffNET TCMR/TI Index | 0.029 | 0.072 | (-0.019; 0.405) |
| Exclusive AMR | 180 | Combined TCMR/TI diagnostics | 0.101 | Histological TCMR/TI Index | 0.121 | -0.020 | (-0.104; 0.021) |
| Exclusive AMR | 180 | BanffNET TCMR/TI Index | 0.029 | Histological TCMR/TI Index | 0.121 | -0.092 | (-0.458; 0.070) |
| Exclusive AMR | 180 | Combined AMR/MVI diagnostics | 0.146 | BanffNET AMR/MVI Index | 0.053 | 0.093 | (0.002; 0.289) |
| Exclusive AMR | 180 | Combined AMR/MVI diagnostics | 0.146 | Histological AMR/MVI Index | 0.027 | 0.119 | (0.000; 0.420) |
| Exclusive AMR | 180 | BanffNET AMR/MVI Index | 0.053 | Histological AMR/MVI Index | 0.027 | 0.026 | (-0.161; 0.318) |
| Exclusive AMR | 180 | Combined Activity diagnostics | 0.157 | BanffNET Activity Index | 0.032 | 0.125 | (0.000; 0.477) |
| Exclusive AMR | 180 | Combined Activity diagnostics | 0.157 | Histological Activity Index | 0.123 | 0.034 | (-0.033; 0.272) |
| Exclusive AMR | 180 | BanffNET Activity Index | 0.032 | Histological Activity Index | 0.123 | -0.091 | (-0.458; 0.163) |
| Exclusive AMR | 180 | Combined Chronicity diagnostics | 0.273 | BanffNET Chronicity Index | 0.291 | -0.018 | (-0.119; 0.032) |
| Exclusive AMR | 180 | Combined Chronicity diagnostics | 0.273 | Histological Chronicity Index | 0.241 | 0.032 | (-0.032; 0.182) |
| Exclusive AMR | 180 | BanffNET Chronicity Index | 0.291 | Histological Chronicity Index | 0.241 | 0.050 | (-0.107; 0.231) |

| | | | | | | | |
|---|---|---|---|---|---|---|---|
| Exclusive Borderline Changes | 271 | Combined TCMR/TI diagnostics | 0.067 | BanffNET TCMR/TI Index | 0.052 | 0.015 | (-0.022; 0.118) |
| Exclusive Borderline Changes | 271 | Combined TCMR/TI diagnostics | 0.067 | Histological TCMR/TI Index | 0.062 | 0.005 | (-0.118; 0.055) |
| Exclusive Borderline Changes | 271 | BanffNET TCMR/TI Index | 0.052 | Histological TCMR/TI Index | 0.062 | -0.010 | (-0.220; 0.100) |
| Exclusive Borderline Changes | 271 | Combined AMR/MVI diagnostics | 0.275 | BanffNET AMR/MVI Index | 0.125 | 0.151 | (0.001; 0.264) |
| Exclusive Borderline Changes | 271 | Combined AMR/MVI diagnostics | 0.275 | Histological AMR/MVI Index | 0.324 | -0.049 | (-0.566; 0.173) |
| Exclusive Borderline Changes | 271 | BanffNET AMR/MVI Index | 0.125 | Histological AMR/MVI Index | 0.324 | -0.200 | (-0.830; 0.118) |
| Exclusive Borderline Changes | 271 | Combined Activity diagnostics | 0.244 | BanffNET Activity Index | 0.036 | 0.208 | (0.031; 0.400) |
| Exclusive Borderline Changes | 271 | Combined Activity diagnostics | 0.244 | Histological Activity Index | 0.256 | -0.012 | (-0.289; 0.279) |
| Exclusive Borderline Changes | 271 | BanffNET Activity Index | 0.036 | Histological Activity Index | 0.256 | -0.219 | (-0.656; 0.110) |
| Exclusive Borderline Changes | 271 | Combined Chronicity diagnostics | 0.714 | BanffNET Chronicity Index | 0.644 | 0.071 | (0.002; 0.198) |
| Exclusive Borderline Changes | 271 | Combined Chronicity diagnostics | 0.714 | Histological Chronicity Index | 0.667 | 0.047 | (-0.017; 0.148) |
| Exclusive Borderline Changes | 271 | BanffNET Chronicity Index | 0.644 | Histological Chronicity Index | 0.667 | -0.023 | (-0.174; 0.107) |
| Exclusive $MVI_{DSA-/C4d-}$ | 136 | Combined TCMR/TI diagnostics | 0.110 | BanffNET TCMR/TI Index | 0.113 | -0.003 | (-0.110; 0.047) |
| Exclusive $MVI_{DSA-/C4d-}$ | 136 | Combined TCMR/TI diagnostics | 0.110 | Histological TCMR/TI Index | 0.066 | 0.044 | (-0.083; 0.438) |
| Exclusive $MVI_{DSA-/C4d-}$ | 136 | BanffNET TCMR/TI Index | 0.113 | Histological TCMR/TI Index | 0.066 | 0.047 | (-0.193; 0.615) |
| Exclusive $MVI_{DSA-/C4d-}$ | 136 | Combined AMR/MVI diagnostics | 0.465 | BanffNET AMR/MVI Index | 0.439 | 0.027 | (-0.057; 0.341) |
| Exclusive $MVI_{DSA-/C4d-}$ | 136 | Combined AMR/MVI diagnostics | 0.465 | Histological AMR/MVI Index | 0.434 | 0.031 | (-0.131; 0.248) |
| Exclusive $MVI_{DSA-/C4d-}$ | 136 | BanffNET AMR/MVI Index | 0.439 | Histological AMR/MVI Index | 0.434 | 0.004 | (-0.275; 0.298) |
| Exclusive $MVI_{DSA-/C4d-}$ | 136 | Combined Activity diagnostics | 0.335 | BanffNET Activity Index | 0.369 | -0.034 | (-0.117; -0.002) |
| Exclusive $MVI_{DSA-/C4d-}$ | 136 | Combined Activity diagnostics | 0.335 | Histological Activity Index | 0.111 | 0.224 | (-0.063; 0.574) |
| Exclusive $MVI_{DSA-/C4d-}$ | 136 | BanffNET Activity Index | 0.369 | Histological Activity Index | 0.111 | 0.258 | (-0.050; 0.623) |
| Exclusive $MVI_{DSA-/C4d-}$ | 136 | Combined Chronicity diagnostics | 0.349 | BanffNET Chronicity Index | 0.263 | 0.086 | (-0.068; 0.361) |
| Exclusive $MVI_{DSA-/C4d-}$ | 136 | Combined Chronicity diagnostics | 0.349 | Histological Chronicity Index | 0.392 | -0.043 | (-0.125; -0.030) |
| Exclusive $MVI_{DSA-/C4d-}$ | 136 | BanffNET Chronicity Index | 0.263 | Histological Chronicity Index | 0.392 | -0.129 | (-0.570; 0.090) |

| | | | | | | | |
|---|---|---|---|---|---|---|---|
| Mixed Rejection | 126 | Combined TCMR/TI diagnostics | 0.048 | BanffNET TCMR/TI Index | 0.002 | 0.047 | (-0.009; 0.278) |
| Mixed Rejection | 126 | Combined TCMR/TI diagnostics | 0.048 | Histological TCMR/TI Index | 0.051 | -0.002 | (-0.069; 0.059) |
| Mixed Rejection | 126 | BanffNET TCMR/TI Index | 0.002 | Histological TCMR/TI Index | 0.051 | -0.049 | (-0.277; 0.052) |
| Mixed Rejection | 126 | Combined AMR/MVI diagnostics | 0.018 | BanffNET AMR/MVI Index | 0.017 | 0.000 | (-0.032; 0.028) |
| Mixed Rejection | 126 | Combined AMR/MVI diagnostics | 0.018 | Histological AMR/MVI Index | 0.007 | 0.011 | (-0.112; 0.191) |
| Mixed Rejection | 126 | BanffNET AMR/MVI Index | 0.017 | Histological AMR/MVI Index | 0.007 | 0.010 | (-0.132; 0.230) |
| Mixed Rejection | 126 | Combined Activity diagnostics | 0.014 | BanffNET Activity Index | 0.008 | 0.006 | (-0.052; 0.071) |
| Mixed Rejection | 126 | Combined Activity diagnostics | 0.014 | Histological Activity Index | 0.002 | 0.012 | (-0.034; 0.120) |
| Mixed Rejection | 126 | BanffNET Activity Index | 0.008 | Histological Activity Index | 0.002 | 0.007 | (-0.089; 0.155) |
| Mixed Rejection | 126 | Combined Chronicity diagnostics | 0.544 | BanffNET Chronicity Index | 0.449 | 0.095 | (0.000; 0.331) |
| Mixed Rejection | 126 | Combined Chronicity diagnostics | 0.544 | Histological Chronicity Index | 0.354 | 0.190 | (0.045; 0.476) |
| Mixed Rejection | 126 | BanffNET Chronicity Index | 0.449 | Histological Chronicity Index | 0.354 | 0.095 | (-0.142; 0.315) |
| No Diagnosis | 2712 | Combined TCMR/TI diagnostics | 0.184 | BanffNET TCMR/TI Index | 0.166 | 0.018 | (0.001; 0.066) |
| No Diagnosis | 2712 | Combined TCMR/TI diagnostics | 0.184 | Histological TCMR/TI Index | 0.156 | 0.028 | (-0.035; 0.110) |
| No Diagnosis | 2712 | BanffNET TCMR/TI Index | 0.166 | Histological TCMR/TI Index | 0.156 | 0.011 | (-0.083; 0.108) |
| No Diagnosis | 2712 | Combined AMR/MVI diagnostics | 0.236 | BanffNET AMR/MVI Index | 0.223 | 0.013 | (-0.005; 0.047) |
| No Diagnosis | 2712 | Combined AMR/MVI diagnostics | 0.236 | Histological AMR/MVI Index | 0.087 | 0.149 | (0.048; 0.246) |
| No Diagnosis | 2712 | BanffNET AMR/MVI Index | 0.223 | Histological AMR/MVI Index | 0.087 | 0.136 | (0.017; 0.244) |
| No Diagnosis | 2712 | Combined Activity diagnostics | 0.165 | BanffNET Activity Index | 0.162 | 0.003 | (-0.003; 0.030) |
| No Diagnosis | 2712 | Combined Activity diagnostics | 0.165 | Histological Activity Index | 0.043 | 0.122 | (0.048; 0.212) |
| No Diagnosis | 2712 | BanffNET Activity Index | 0.162 | Histological Activity Index | 0.043 | 0.119 | (0.028; 0.215) |
| No Diagnosis | 2712 | Combined Chronicity diagnostics | 0.434 | BanffNET Chronicity Index | 0.378 | 0.056 | (0.016; 0.106) |
| No Diagnosis | 2712 | Combined Chronicity diagnostics | 0.434 | Histological Chronicity Index | 0.408 | 0.026 | (-0.001; 0.061) |
| No Diagnosis | 2712 | BanffNET Chronicity Index | 0.378 | Histological Chronicity Index | 0.408 | -0.030 | (-0.111; 0.042) |

The categories Exclusive BKPyVAN and Exclusive Probable AMR had insufficient events for survival analysis.

**Supplementary Table 7. Differences in explained variability ($R^2$) of BHOT molecular Indices by Banff diagnoses, histological indices, BanffNET Indices and the combination of the three in the validation cohort (n = 3863).**

| Model 1 | $R^2$ | Model 2 | $R^2$ | $R^2$ difference | 95% CI |
|---|---|---|---|---|---|
| Combined TCMR/TI diagnostics | 0.583 | BanffNET TCMR/TI Index | 0.499 | 0.084 | (0.057; 0.117) |
| Combined TCMR/TI diagnostics | 0.583 | Histological TCMR/TI Index | 0.468 | 0.115 | (0.087; 0.158) |
| Combined TCMR/TI diagnostics | 0.583 | Banff TCMR | 0.280 | 0.303 | (0.260; 0.343) |
| BanffNET TCMR/TI Index | 0.499 | Histological TCMR/TI Index | 0.468 | 0.031 | (-0.023; 0.098) |
| BanffNET TCMR/TI Index | 0.499 | Banff TCMR | 0.280 | 0.219 | (0.165; 0.280) |
| Histological TCMR/TI Index | 0.468 | Banff TCMR | 0.280 | 0.188 | (0.151; 0.231) |
| Combined AMR/MVI diagnostics | 0.429 | BanffNET AMR/MVI Index | 0.249 | 0.180 | (0.136; 0.237) |
| Combined AMR/MVI diagnostics | 0.429 | Histological AMR/MVI Index | 0.369 | 0.060 | (0.033; 0.102) |
| Combined AMR/MVI diagnostics | 0.429 | Banff AMR/MVI | 0.295 | 0.135 | (0.086; 0.179) |
| BanffNET AMR/MVI Index | 0.249 | Histological AMR/MVI Index | 0.369 | -0.120 | (-0.196; -0.049) |
| BanffNET AMR/MVI Index | 0.249 | Banff AMR/MVI | 0.295 | -0.045 | (-0.130; 0.025) |
| Histological AMR/MVI Index | 0.369 | Banff AMR/MVI | 0.295 | 0.075 | (0.043; 0.111) |
| Combined Activity diagnostics | 0.567 | BanffNET Activity Index | 0.467 | 0.100 | (0.066; 0.148) |
| Combined Activity diagnostics | 0.567 | Histological Activity Index | 0.444 | 0.124 | (0.091; 0.163) |
| Combined Activity diagnostics | 0.567 | Banff Any Diagnosis | 0.336 | 0.232 | (0.202; 0.282) |
| BanffNET Activity Index | 0.467 | Histological Activity Index | 0.444 | 0.024 | (-0.056; 0.094) |
| BanffNET Activity Index | 0.467 | Banff Any Diagnosis | 0.336 | 0.132 | (0.078; 0.201) |
| Histological Activity Index | 0.444 | Banff Any Diagnosis | 0.336 | 0.108 | (0.082; 0.145) |
| Combined Chronicity diagnostics | 0.574 | BanffNET Chronicity Index | 0.458 | 0.115 | (0.079; 0.154) |

| | | | | | |
|---|---|---|---|---|---|
| Combined Chronicity diagnostics | 0.574 | Histological Chronicity Index | 0.453 | 0.121 | (0.080; 0.171) |
| BanffNET Chronicity Index | 0.458 | Histological Chronicity Index | 0.453 | 0.006 | (-0.072; 0.093) |

**Supplementary Table 8. Overview of missing data strategies in cohorts and resulting sample sizes for each statistical model.**

| Cohort | Total number of observations | Missing data | Missing data strategy | Models | | Number of observations in models | Number of graft failure events in models |
|---|---|---|---|---|---|---|---|
| | | | | Independent variables | Dependent variables | | |
| Dutch training cohort | 2544 | 83 missing Banff TCMR due to missing v lesion, 1566 missing Banff AMR/MVI due to missing C4d (n = 1081) and/or missing DSA (n = 1367), 1595 missing Banff Any Diagnosis due to missing C4d (n = 1081), missing DSA (n = 1367) and missing v (n = 83), 670 missing graft survival time, 83 missing histological TCMR/TI index due to missing v lesion, 1081 | Banff TCMR, Banff AMR/MVI, Banff Any Diagnosis, histological TCMR/TI index, histological AMR/MVI index, histological Activity index and histological Chronicity index were imputed using multiple imputation of the missing lesion scores as well as DSA and C4d. Missing graft survival time was treated as censoring at time 0, leading to exclusion of these observations from the survival models, but IPCW | logit(t_NET), logit(i_NET), logit(ptc_NET), logit(v_NET), logit(ti_NET), logit(tifta_NET) | Banff TCMR | 2544 | NA |
| | | | | logit(g_NET), logit(ptc_NET), logit(v_NET), logit(tma_NET), logit(cg_NET), logit(mm_NET) | Banff AMR/MVI | 2544 | NA |
| | | | | logit(t_NET), logit(i_NET), logit(v_NET), | Hazard of graft failure | 1874 | 397 |

| | | | | | | | |
|---|---|---|---|---|---|---|---|
| | | missing histological AMR/MVI index due to missing C4d, 1130 missing histological Activity index due to missing C4d (n = 1081) and/or missing v (n = 83), 83 missing histological Chronicity index due to missing cv (n = 83) | was used to improve generalizability of the estimates. | logit(g_NET), logit(ptc_NET), logit(tma_NET) | | | |
| | | | | logit(ifta_NET), logit(cg_NET), logit(mm_NET), logit(gs_NET), logit(fsgs_NET) | Hazard of graft failure | 1874 | 397 |
| | | | | BanffNET TCMR/TI Index | Banff TCMR | 2544 | NA |
| | | | | BanffNET AMR/MVI Index | Banff MVI | 2544 | NA |
| | | | | BanffNET Activity Index | Banff Any Diagnosis | 2544 | NA |
| | | | | BanffNET TCMR/TI Index | Histological TCMR/TI Index | 2544 | NA |
| | | | | BanffNET AMR/MVI Index | Histological AMR/MVI Index | 2544 | NA |

| | | | | | | | |
|---|---|---|---|---|---|---|---|
| | | | | BanffNET Activity Index | Histological Activity Index | 2544 | NA |
| | | | | BanffNET Chronicity Index | Histological Chronicity Index | 2544 | NA |
| | | | | Banff TCMR | Hazard of graft failure | 1874 | 397 |
| | | | | Banff AMR/MVI | Hazard of graft failure | 1874 | 397 |
| | | | | Banff Any Diagnosis | Hazard of graft failure | 1874 | 397 |
| | | | | Histological TCMR/TI Index | Hazard of graft failure | 1874 | 397 |
| | | | | Histological AMR/MVI Index | Hazard of graft failure | 1874 | 397 |
| | | | | Histological Activity Index | Hazard of graft failure | 1874 | 397 |
| | | | | Histological Chronicity Index | Hazard of graft failure | 1874 | 397 |
| | | | | BanffNET TCMR/TI Index | Hazard of graft failure | 1874 | 397 |

| | | | | | | | |
|---|---|---|---|---|---|---|---|
| | | | | BanffNET AMR/MVI Index | Hazard of graft failure | 1874 | 397 |
| | | | | BanffNET Activity Index | Hazard of graft failure | 1874 | 397 |
| | | | | BanffNET Chronicity Index | Hazard of graft failure | 1874 | 397 |
| | | | | Banff TCMR, Histological TCMR/TI Index, BanffNET TCMR/TI Index | Hazard of graft failure | 1874 | 397 |
| | | | | Banff AMR/MVI, histological AMR/MVI index, BanffNET AMR/MVI Index | Hazard of graft failure | 1874 | 397 |
| | | | | Banff Any Diagnosis, histological Activity index, BanffNET Activity Index | Hazard of graft failure | 1874 | 397 |

| | | | | | | | |
|---|---|---|---|---|---|---|---|
| | | | | histological Chronicity index, BanffNET Chronicity Index | Hazard of graft failure | 1874 | 397 |
| Validation cohort | 4122 | 5 missing histological TCMR/TI index due to missing lesion scores, 254 missing histological AMR/MVI index due to missing lesion scores or missing C4d, 256 missing histological Activity index due to missing lesion scores or missing C4d, 6 missing histological Chronicity index due to missing lesion scores. 3758 biopsies have missing values for all BHOT molecular Indices. | Analyses not involving molecular Indices were performed on observations with complete histological data. Analyses involving molecular Indices were also performed on observations with complete histological data, with multiple imputation to account for the missing molecular Indices. | BanffNET TCMR/TI Index | Banff TCMR | 3863 | NA |
| | | | | BanffNET AMR/MVI Index | Banff MVI | 3863 | NA |
| | | | | BanffNET Activity Index | Banff Any Diagnosis | 3863 | NA |
| | | | | BanffNET TCMR/TI Index | Histological TCMR/TI Index | 3863 | NA |
| | | | | BanffNET AMR/MVI Index | Histological AMR/MVI Index | 3863 | NA |
| | | | | BanffNET Activity Index | Histological Activity Index | 3863 | NA |
| | | | | BanffNET Chronicity Index | Histological Chronicity Index | 3863 | NA |

| | | | | | | | |
|---|---|---|---|---|---|---|---|
| | | | | Banff TCMR | Hazard of graft failure | 3863 | 207 |
| | | | | Banff AMR/MVI | Hazard of graft failure | 3863 | 207 |
| | | | | Banff Any Diagnosis | Hazard of graft failure | 3863 | 207 |
| | | | | Histological TCMR/TI Index | Hazard of graft failure | 3863 | 207 |
| | | | | Histological AMR/MVI Index | Hazard of graft failure | 3863 | 207 |
| | | | | Histological Activity Index | Hazard of graft failure | 3863 | 207 |
| | | | | Histological Chronicity Index | Hazard of graft failure | 3863 | 207 |
| | | | | BanffNET TCMR/TI Index | Hazard of graft failure | 3863 | 207 |
| | | | | BanffNET AMR/MVI Index | Hazard of graft failure | 3863 | 207 |
| | | | | BanffNET Activity Index | Hazard of graft failure | 3863 | 207 |
| | | | | BanffNET Chronicity Index | Hazard of graft failure | 3863 | 207 |

| | | | | | | | |
|---|---|---|---|---|---|---|---|
| | | | | Banff TCMR, Histological TCMR/TI Index, BanffNET TCMR/TI Index | Hazard of graft failure | 3863 | 207 |
| | | | | Banff AMR/MVI, histological AMR/MVI index, BanffNET AMR/MVI Index | Hazard of graft failure | 3863 | 207 |
| | | | | Banff Any Diagnosis, histological Activity index, BanffNET Activity Index | Hazard of graft failure | 3863 | 207 |
| | | | | histological Chronicity index, BanffNET Chronicity Index | Hazard of graft failure | 3863 | 207 |

| | | | | | | | |
|---|---|---|---|---|---|---|---|
| | | | | Banff TCMR | Molecular TCMR/TI Index | 3863 | NA |
| | | | | Banff AMR/MVI | Molecular AMR/MVI Index | 3863 | NA |
| | | | | Banff Any Diagnosis | Molecular Activity Index | 3863 | NA |
| | | | | Histological TCMR/TI Index | Molecular TCMR/TI Index | 3863 | NA |
| | | | | Histological AMR/MVI Index | Molecular AMR/MVI Index | 3863 | NA |
| | | | | Histological Activity Index | Molecular Activity Index | 3863 | NA |
| | | | | Histological Chronicity Index | Molecular Chronicity Index | 3863 | NA |

| | | | | | | | |
|---|---|---|---|---|---|---|---|
| | | | | BanffNET TCMR/TI Index | Molecular TCMR/TI Index | 3863 | NA |
| | | | | BanffNET AMR/MVI Index | Molecular AMR/MVI Index | 3863 | NA |
| | | | | BanffNET Activity Index | Molecular Activity Index | 3863 | NA |
| | | | | BanffNET Chronicity Index | Molecular Chronicity Index | 3863 | NA |
| | | | | Banff TCMR, Histological TCMR/TI Index, BanffNET TCMR/TI Index | Molecular TCMR/TI Index | 3863 | NA |
| | | | | Banff AMR/MVI, histological AMR/MVI index, BanffNET AMR/MVI Index | Molecular AMR/MVI Index | 3863 | NA |

| | | | | | | | |
|---|---|---|---|---|---|---|---|
| | | | | Banff Any Diagnosis, histological Activity index, BanffNET Activity Index | Molecular Activity Index | 3863 | NA |
| | | | | histological Chronicity index, BanffNET Chronicity Index | Molecular Chronicity Index | 3863 | NA |
| Reader study cohort | 2412 pathologist-biopsy pairs | 81 pathologist-biopsy pairs with missing Banff TCMR due to missing v lesion, 58 missing Banff Any Diagnosis due to missing v lesion, 117 pathologist-biopsy pairs with missing histological TCMR/TI index due to missing v lesion, 138 pathologist-biopsy | Analyses were conducted on pathologist-biopsy pairs with complete observations. Histological variables were analyzed on the level of pathologist-biopsy pairs while BanffNET Indices were analyzed on the level of biopsies. | Banff TCMR | Multi-observer aggregated TCMR diagnosis | 2272 pathologist-biopsy pairs | NA |
| | | | | Banff MVI | Multi-observer aggregated MVI diagnosis | 2272 pathologist-biopsy pairs | NA |

| | | | | | | | |
|---|---|---|---|---|---|---|---|
| | | pairs with missing histological Chronicity index due to missing v lesion and missing cv lesion, 2 biopsies with missing BanffNET Indices. | | Banff Any Diagnosis | Multi-observer aggregated Any Diagnosis | 2272 pathologist-biopsy pairs | NA |
| | | | | Histological TCMR/TI Index | Multi-observer aggregated TCMR diagnosis | 2272 pathologist-biopsy pairs | NA |
| | | | | Histological AMR/MVI Index | Multi-observer aggregated MVI diagnosis | 2272 pathologist-biopsy pairs | NA |
| | | | | Histological Activity Index | Multi-observer aggregated Any Diagnosis | 2272 pathologist-biopsy pairs | NA |
| | | | | BanffNET TCMR/TI Index | Multi-observer | 36 biopsies | NA |

| | | | | | | | |
|---|---|---|---|---|---|---|---|
| | | | | | aggregated TCMR diagnosis | | |
| | | | | BanffNET AMR/MVI Index | Multi-observer aggregated MVI diagnosis | 36 biopsies | NA |
| | | | | BanffNET Activity Index | Multi-observer aggregated Any Diagnosis | 36 biopsies | NA |
| | | | | Banff TCMR, histological TCMR/TI index, BanffNET TCMR/TI Index | Multi-observer aggregated TCMR diagnosis | 2272 pathologist-biopsy pairs | NA |
| | | | | Banff MVI, histological AMR/MVI index, | Multi-observer aggregated | 2272 pathologist-biopsy pairs | NA |

| | | | | | | | |
|---|---|---|---|---|---|---|---|
| | | | | BanffNET AMR/MVI Index | MVI diagnosis | | |
| | | | | Banff Any Diagnosis, histological Activity index, BanffNET Activity Index | Multi-observer aggregated Any Diagnosis | 2272 pathologist-biopsy pairs | NA |
| | | | | Banff TCMR | Multi-observer aggregated histological TCMR/TI index | 2272 pathologist-biopsy pairs | NA |
| | | | | Banff AMR/MVI | Multi-observer aggregated histological AMR/MVI index | 2272 pathologist-biopsy pairs | NA |

| | | | | | | | |
|---|---|---|---|---|---|---|---|
| | | | | Banff Any Diagnosis | Multi-observer aggregated histological Activity index | 2272 pathologist-biopsy pairs | NA |
| | | | | Histological TCMR/TI Index | Multi-observer aggregated histological TCMR/TI index | 2272 pathologist-biopsy pairs | NA |
| | | | | Histological AMR/MVI Index | Multi-observer aggregated histological AMR/MVI index | 2272 pathologist-biopsy pairs | NA |
| | | | | Histological Activity Index | Multi-observer aggregated histological | 2272 pathologist-biopsy pairs | NA |

| | | | | | | | |
|---|---|---|---|---|---|---|---|
| | | | | | Activity index | | |
| | | | | Histological Chronicity Index | Multi-observer aggregated histological Chronicity index | 2272 pathologist-biopsy pairs | NA |
| | | | | BanffNET TCMR/TI Index | Multi-observer aggregated histological TCMR/TI index | 36 biopsies | NA |
| | | | | BanffNET AMR/MVI Index | Multi-observer aggregated histological AMR/MVI index | 36 biopsies | NA |
| | | | | BanffNET Activity Index | Multi-observer | 36 biopsies | NA |

| | | | | | | | |
|---|---|---|---|---|---|---|---|
| | | | | | aggregated histological Activity index | | |
| | | | | BanffNET Chronicity Index | Multi-observer aggregated histological Chronicity index | 36 biopsies | NA |
| | | | | Banff TCMR, histological TCMR/TI index, BanffNET TCMR/TI Index | Multi-observer aggregated histological TCMR/TI index | 2272 pathologist-biopsy pairs | NA |
| | | | | Banff AMR/MVI, histological AMR/MVI index, BanffNET AMR/MVI Index | Multi-observer aggregated histological AMR/MVI index | 2272 pathologist-biopsy pairs | NA |

| | | | | | | | |
|---|---|---|---|---|---|---|---|
| | | | | Banff Any Diagnosis, histological Activity index, BanffNET Activity Index | Multi-observer aggregated histological Activity index | 2272 pathologist-biopsy pairs | NA |
| | | | | histological Chronicity index, BanffNET Chronicity Index | Multi-observer aggregated histological Chronicity index | 2272 pathologist-biopsy pairs | NA |

### Supplementary figures

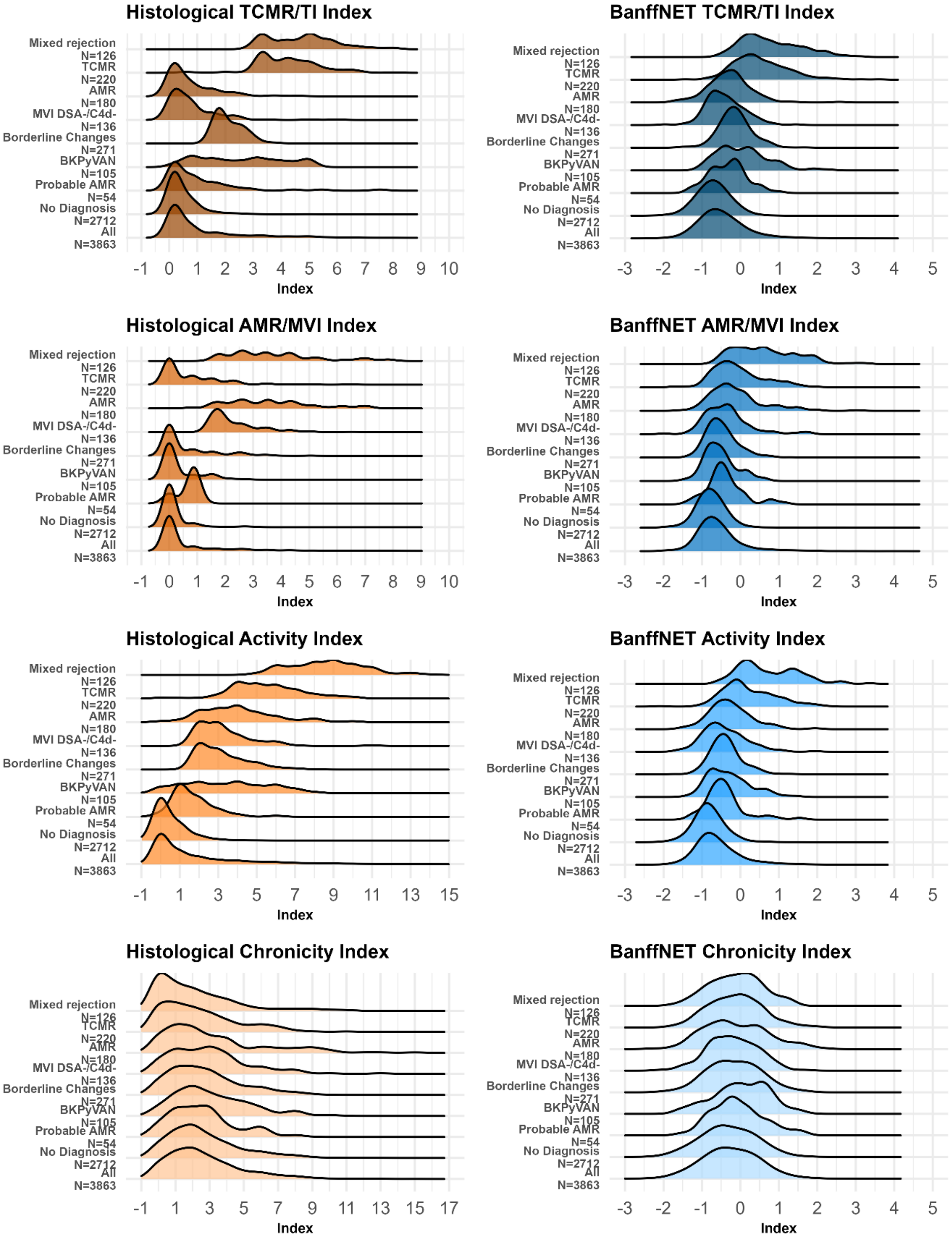

**Supplementary Figure 1. Distributions of histological indices and BanffNET Indices within Banff categories in the validation cohort (n = 3863).** Only exclusive diagnoses were included in the categories to prevent overlap.

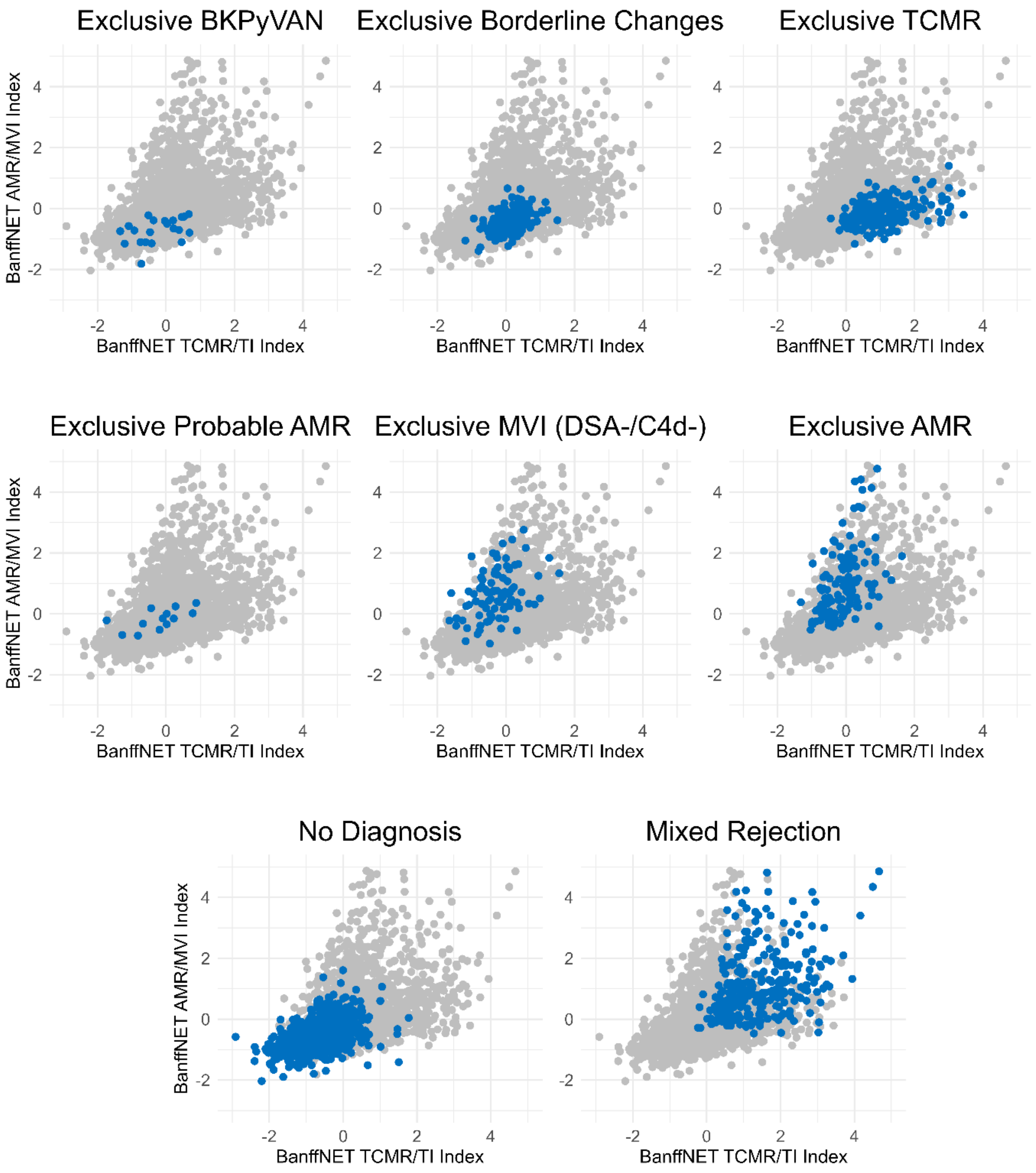

**Supplementary Figure 2. Banff diagnoses in the continuous 2-dimensional diagnostic space in the training cohort (n = 2544).** The 2-dimensional diagnostic space is defined by the BanffNET TCMR/TI Index and the BanffNET AMR/MVI Index.

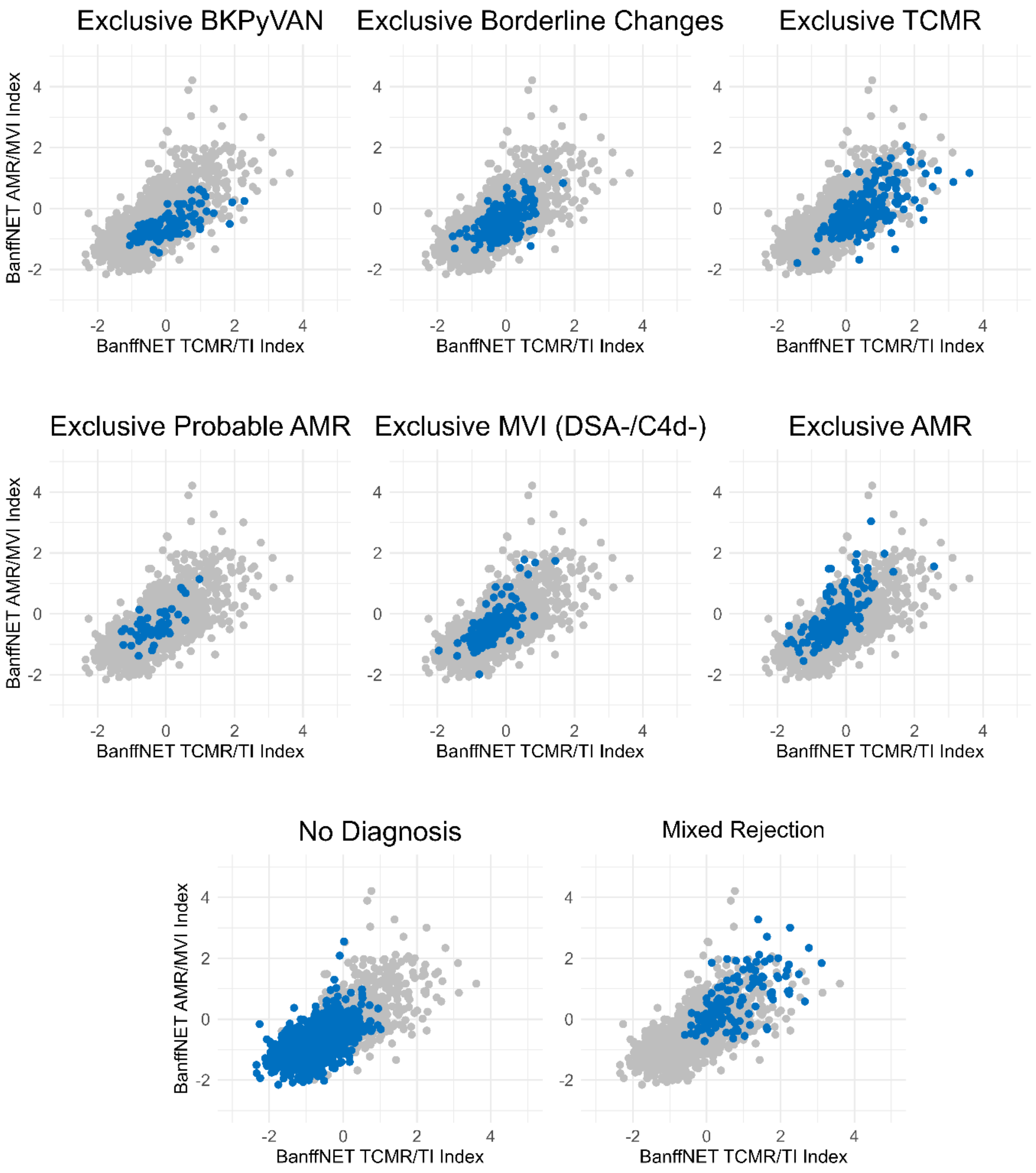

**Supplementary Figure 3. Banff diagnoses in the continuous 2-dimensional diagnostic space in the validation cohort (n = 3863).** The 2-dimensional diagnostic space is defined by the BanffNET TCMR/TI Index and the BanffNET AMR/MVI Index.

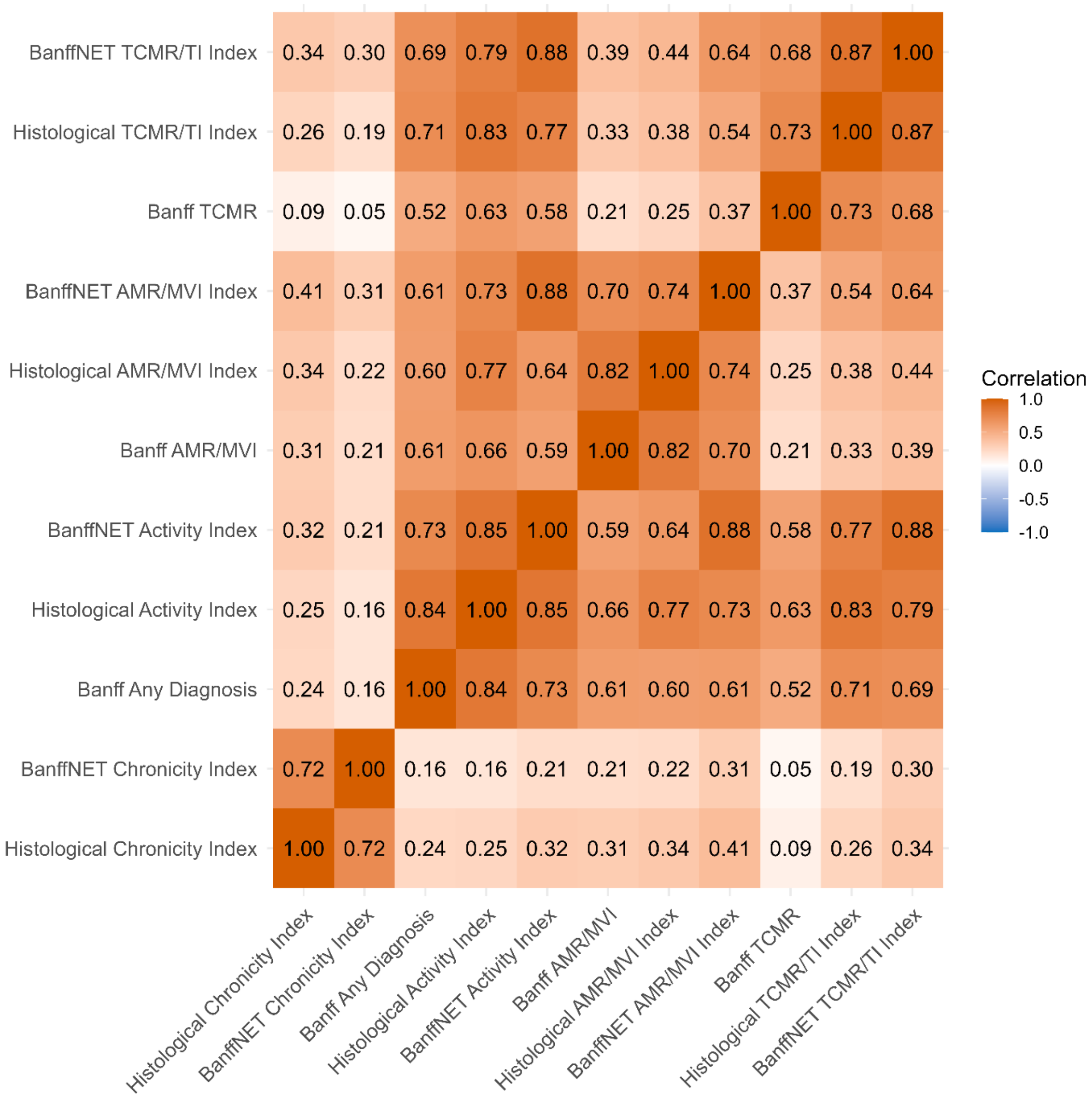

**Supplementary Figure 4. Spearman correlations between diagnostics in the training cohort (n = 2544).** A comparison of Banff diagnoses, BanffNET Indices and histological indices.

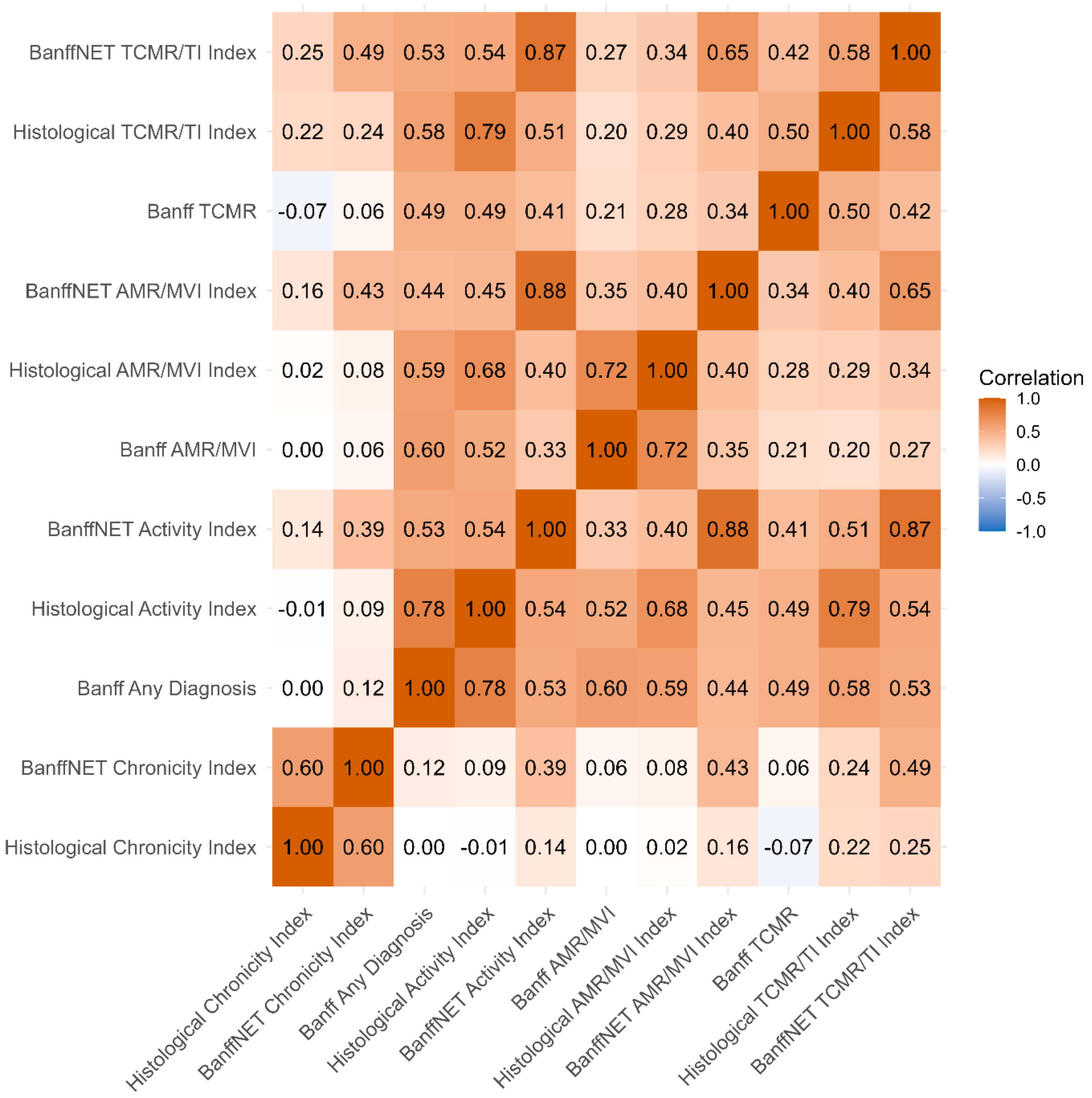

**Supplementary Figure 5. Spearman correlations between diagnostics in the validation cohort (n = 3863).** A comparison of Banff diagnoses, BanffNET Indices and histological indices.

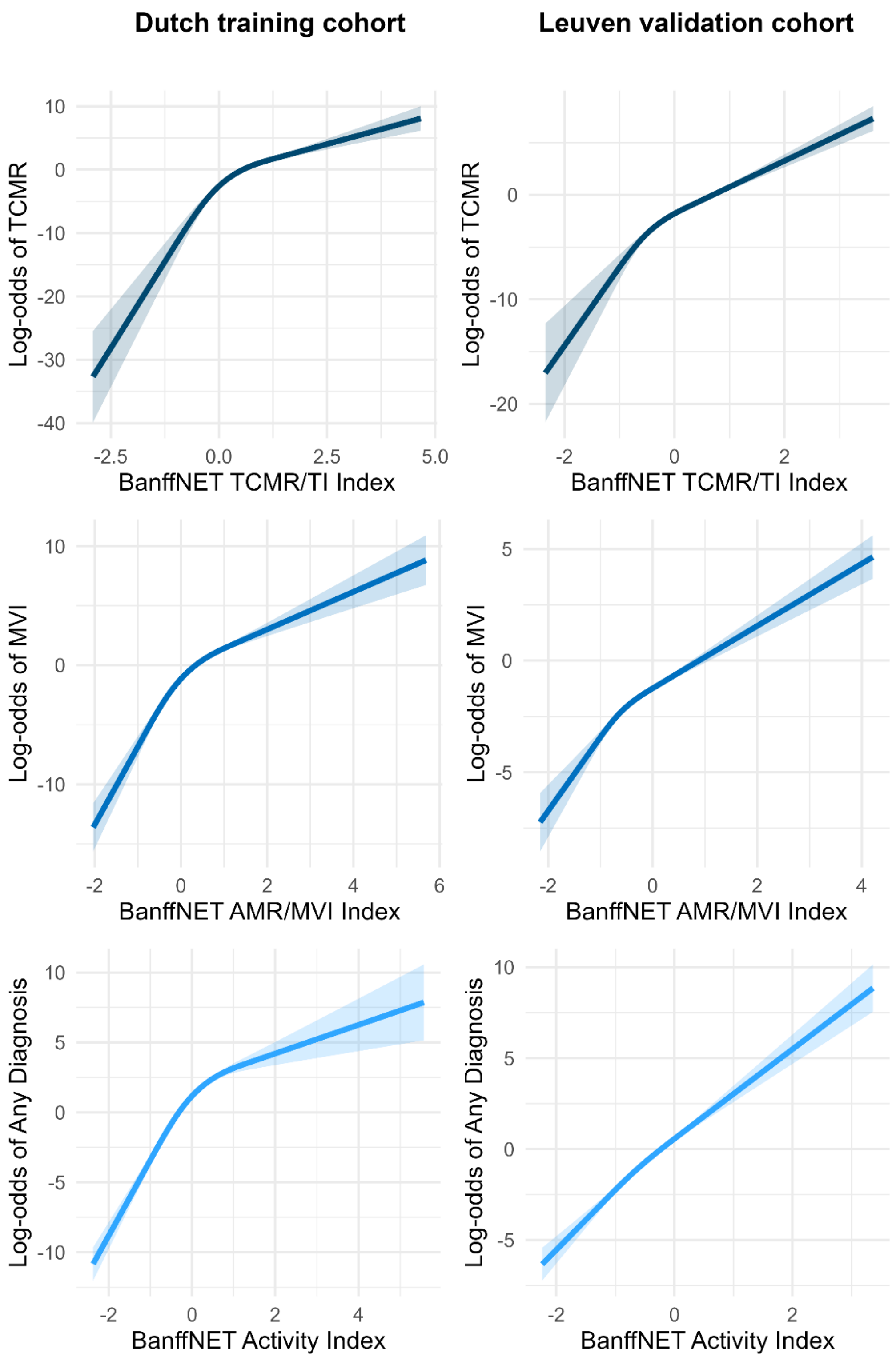

**Supplementary Figure 6. Linearity of associations with Banff diagnoses in the training cohort (n = 2544) and the validation cohort (n = 3863).** Effect plots with pointwise 95% confidence intervals to evaluate linearity in the associations of BanffNET Indices with log-odds of diagnoses in the Dutch training cohort (n = 2544) and validation cohort (n = 3863).

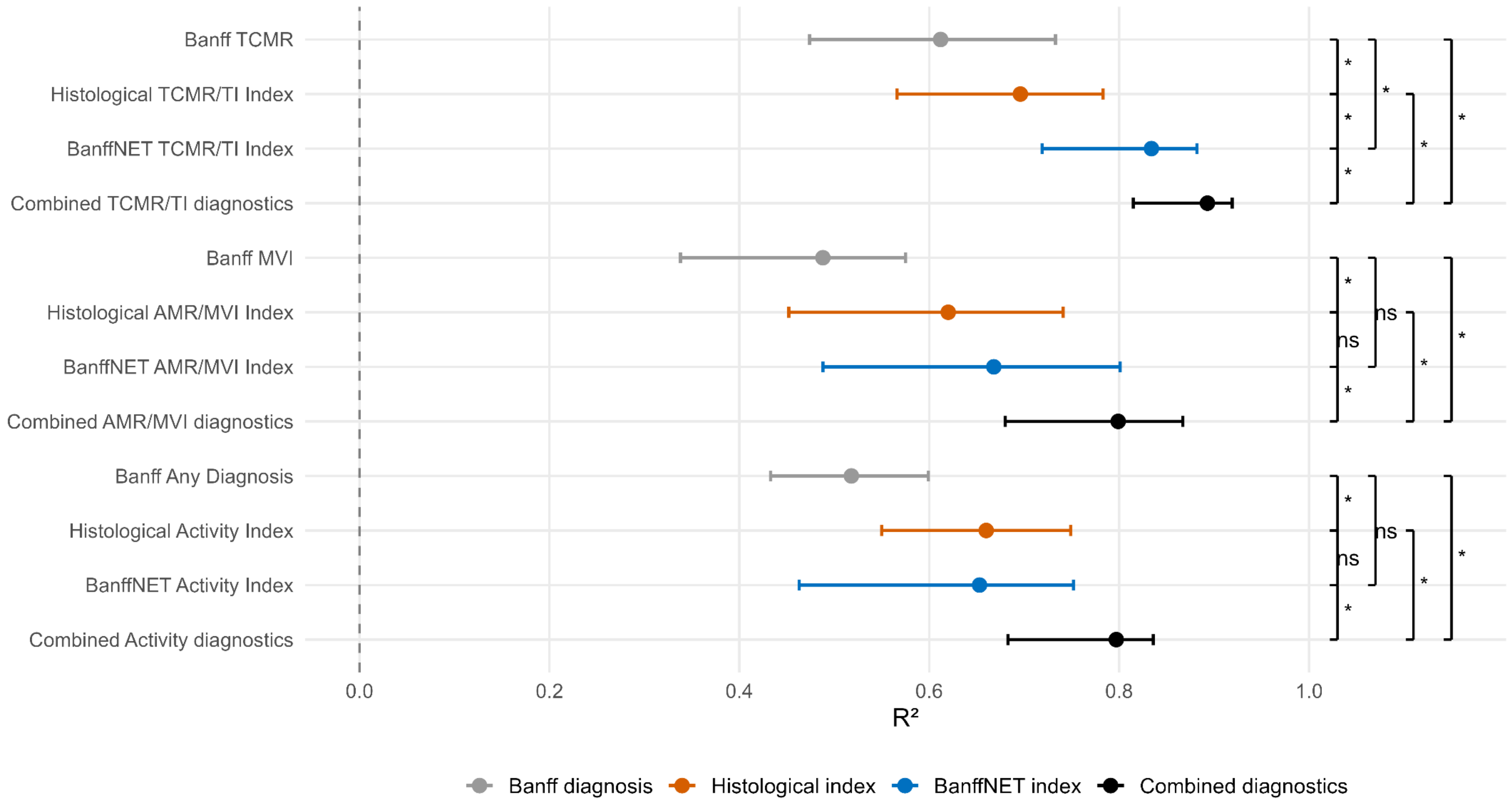

**Supplementary Figure 7. $R^2$ values for aggregated Banff diagnoses in the multi-reader cohort (n = 36 biopsies scored by 67 pathologists).** 95% Confidence bounds were calculated for the $R^2$ values as well as for differences between $R^2$ values. Differences were considered significant if zero was not included in the confidence bounds. Significant differences were indicated with an asterisk, while non-significant differences were indicated with 'ns'.

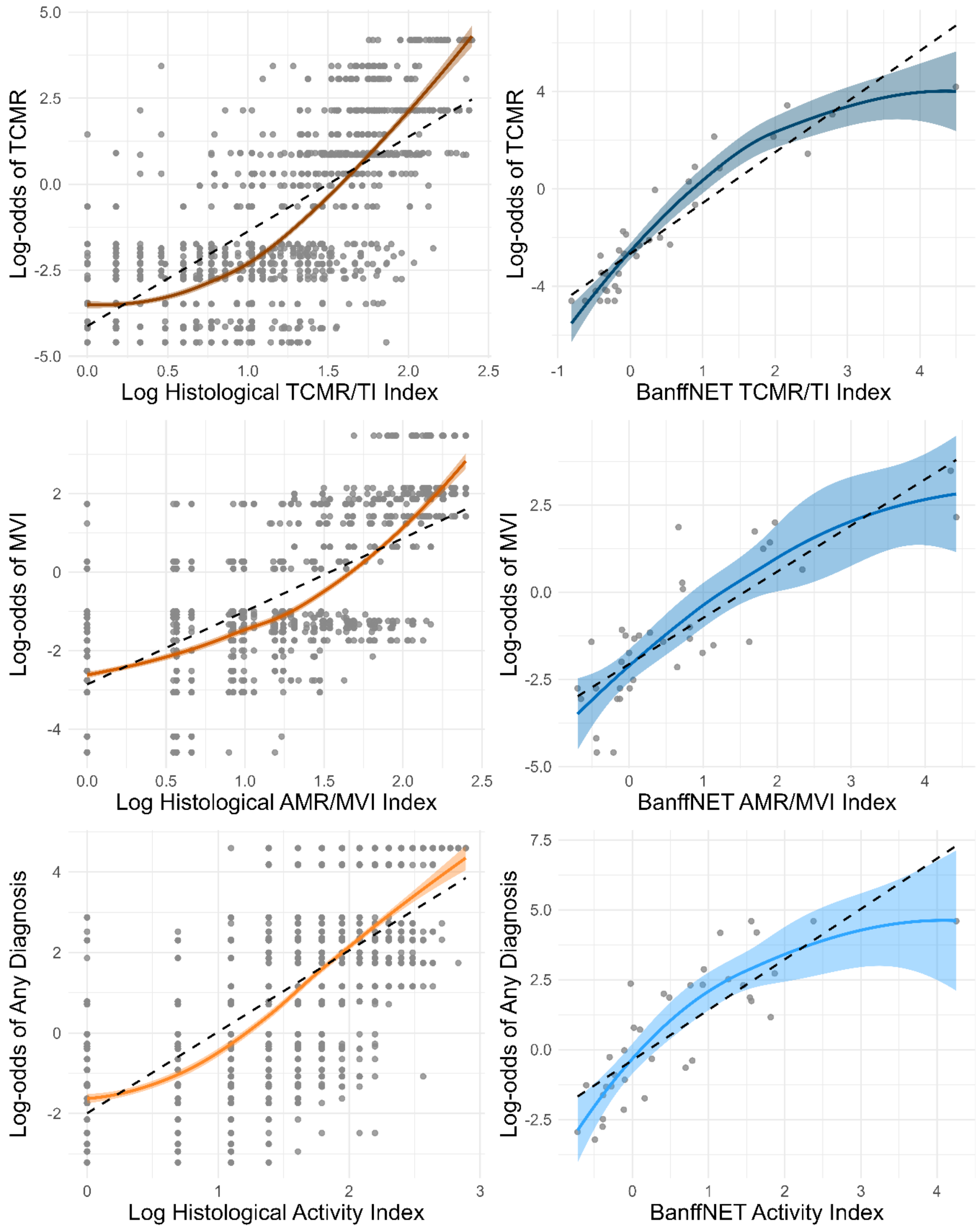

**Supplementary Figure 8. Linearity of associations with multi-observer reference standards in the multi-reader cohort (n = 36 biopsies scored by 67 pathologists).** Loess-smoothed scatter plots with pointwise 95% confidence intervals to evaluate linearity in the associations of log-transformed histological indices and BanffNET Indices with log-odds of aggregated diagnoses.

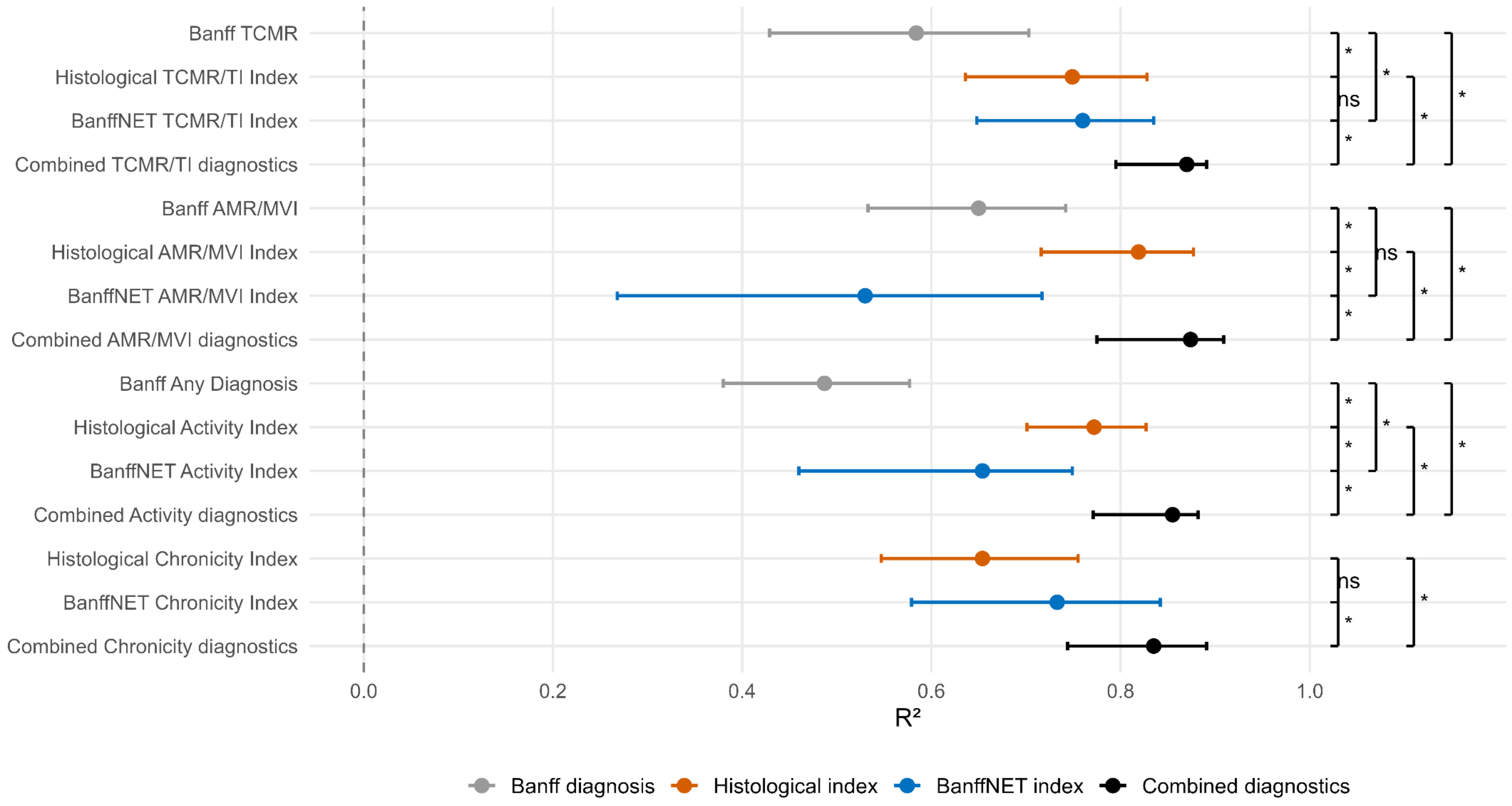

**Supplementary Figure 9. $R^2$ values for aggregated histological indices in the multi-reader cohort (n = 36 biopsies scored by 67 pathologists).** 95% Confidence bounds were calculated for the $R^2$ values as well as for differences between $R^2$ values. Differences were considered significant if zero was not included in the confidence bounds. Significant differences were indicated with an asterisk, while non-significant differences were indicated with 'ns'.

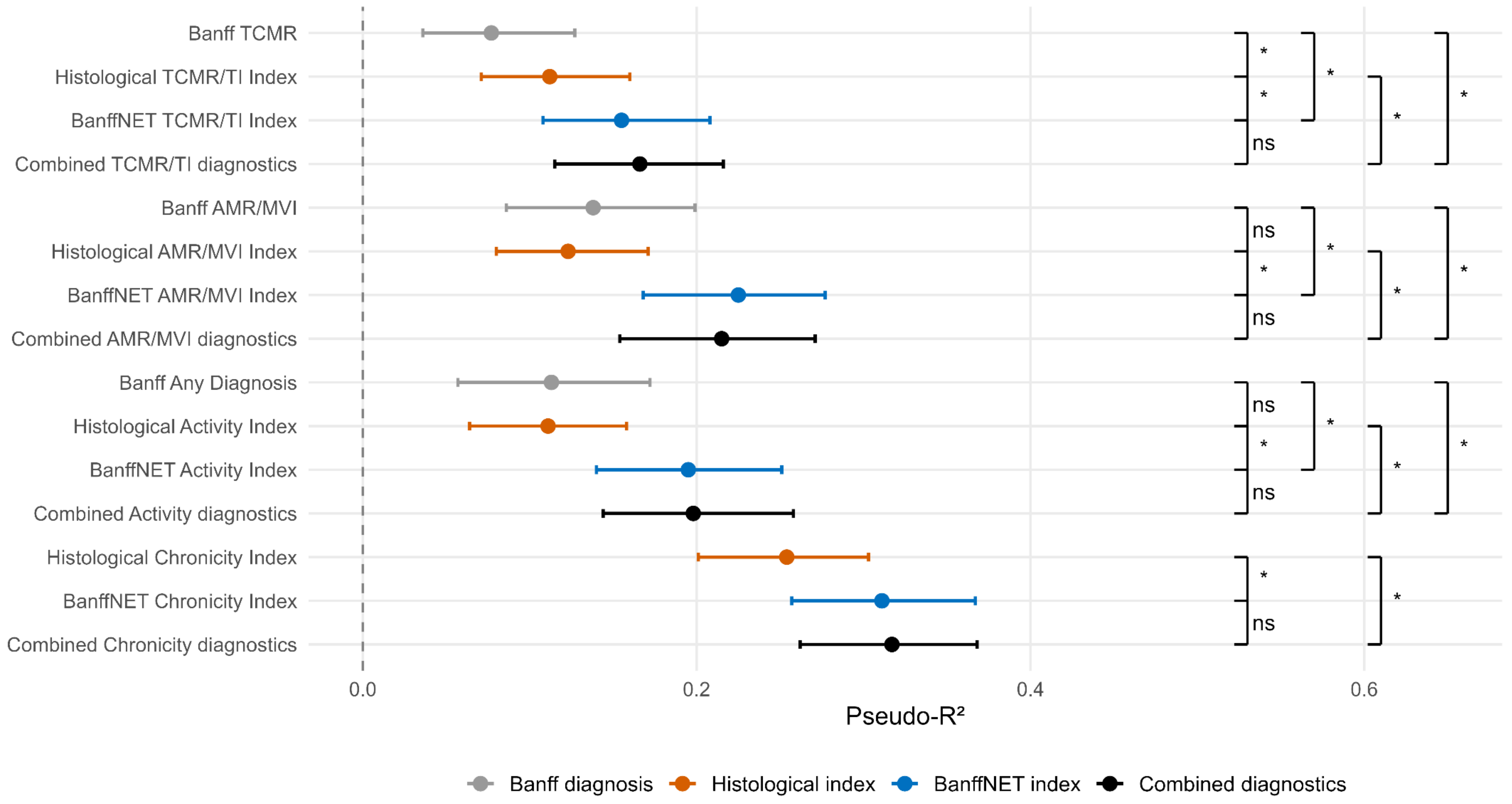

**Supplementary Figure 10. Pseudo-$R^2$ values for the hazard of kidney graft failure in the training cohort (n = 1874).** 95% Confidence bounds were calculated for the $R^2$ values as well as for differences between $R^2$ values. Differences were considered significant if zero was not included in the confidence bounds. Significant differences were indicated with an asterisk, while non-significant differences were indicated with 'ns'.

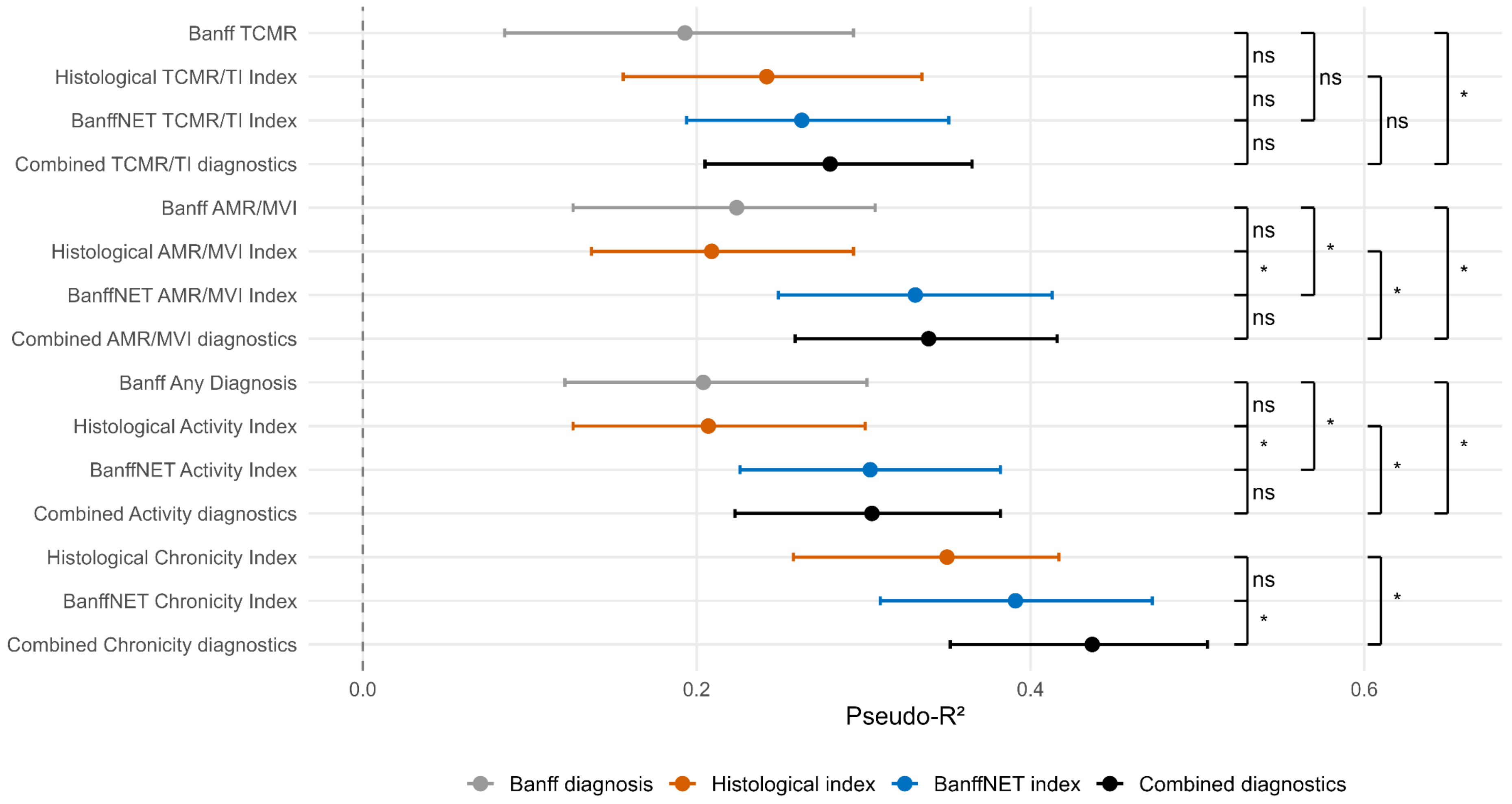

**Supplementary Figure 11. Pseudo-$R^2$ values for the hazard of kidney graft failure in the validation cohort (n = 3863).** 95% Confidence bounds were calculated for the $R^2$ values as well as for differences between $R^2$ values. Differences were considered significant if zero was not included in the confidence bounds. Significant differences were indicated with an asterisk, while non-significant differences were indicated with 'ns'.

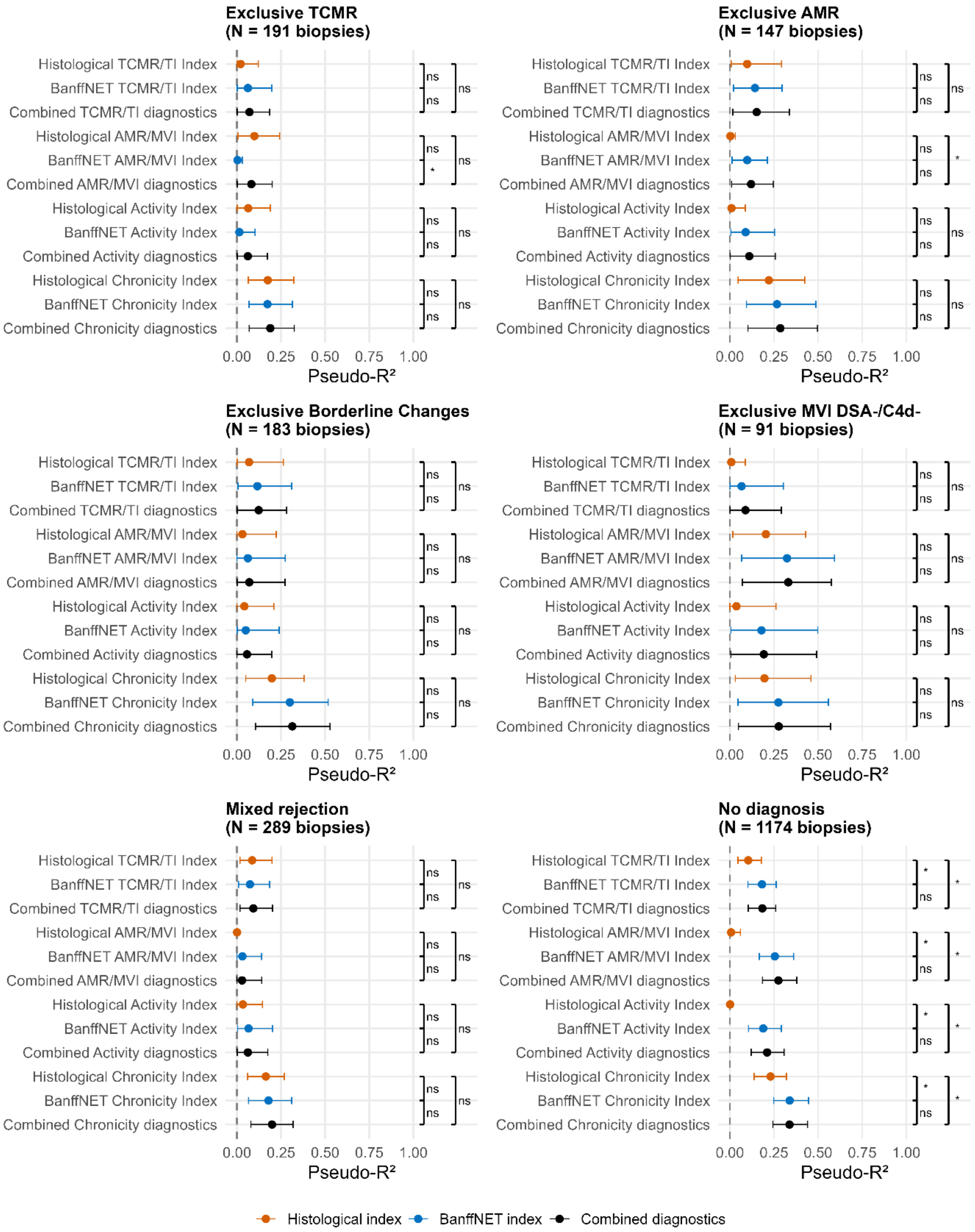

**Supplementary Figure 12. Explaining residual graft survival variability within mutually exclusive Banff categories in the training cohort (n = 1874).** Estimates denote Royston-Sauerbrei pseudo-$R^2$ values for the hazard of kidney graft failure. 95% Confidence bounds were calculated for the pseudo-$R^2$ values as well as for differences between pseudo-$R^2$ values. Differences were considered significant if zero was not included in the confidence bounds. Significant differences were indicated with an asterisk, while non-significant differences were indicated with 'ns'.

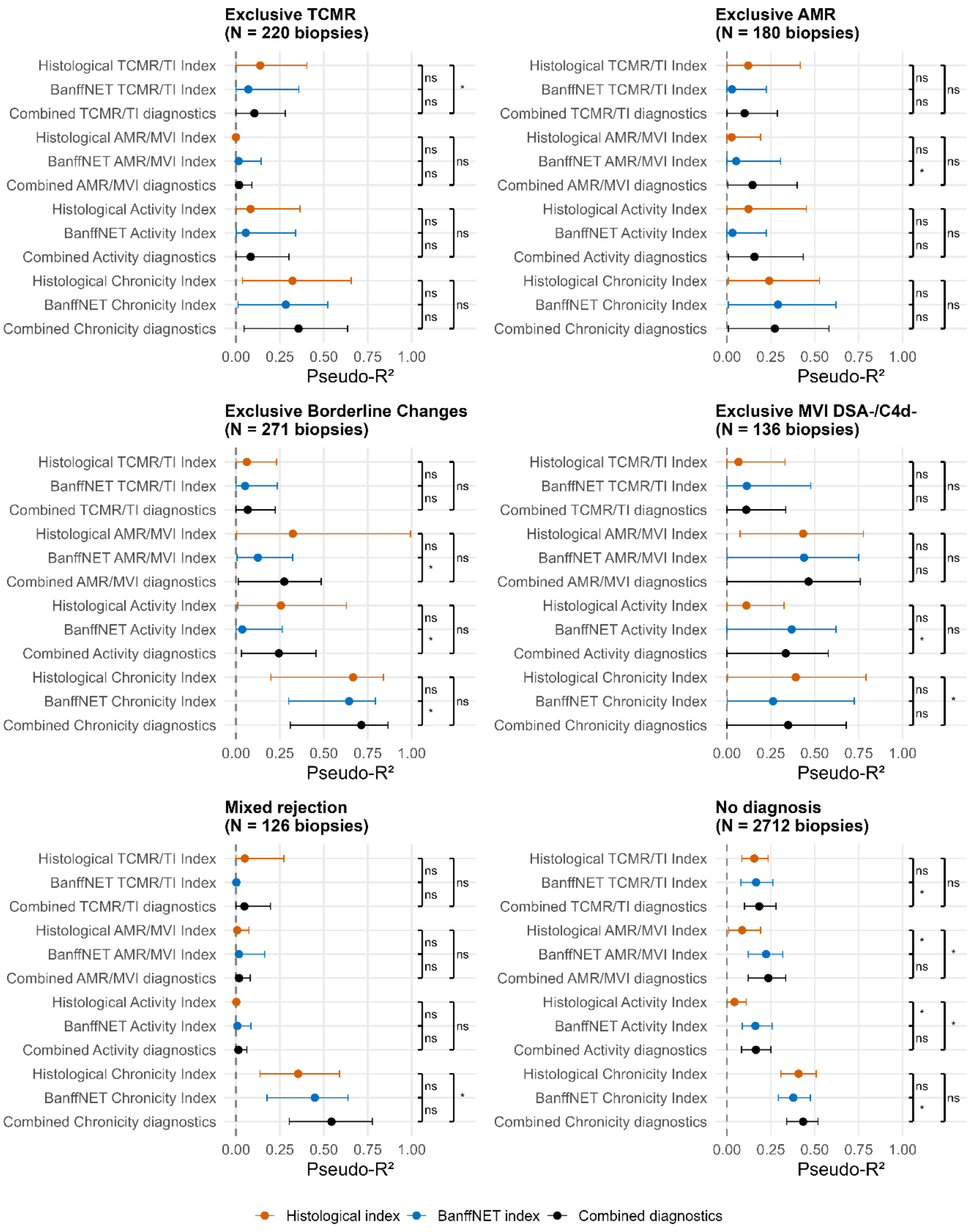

**Supplementary Figure 13. Explaining residual graft survival variability within mutually exclusive Banff categories in the validation cohort (n = 3863).** Estimates denote Royston-Sauerbrei pseudo-$R^2$ values for the hazard of kidney graft failure. 95% Confidence bounds were calculated for the pseudo-$R^2$ values as well as for differences between pseudo-$R^2$ values. Differences were considered significant if zero was not included in the confidence bounds. Significant differences were indicated with an asterisk, while non-significant differences were indicated with ‘ns’.

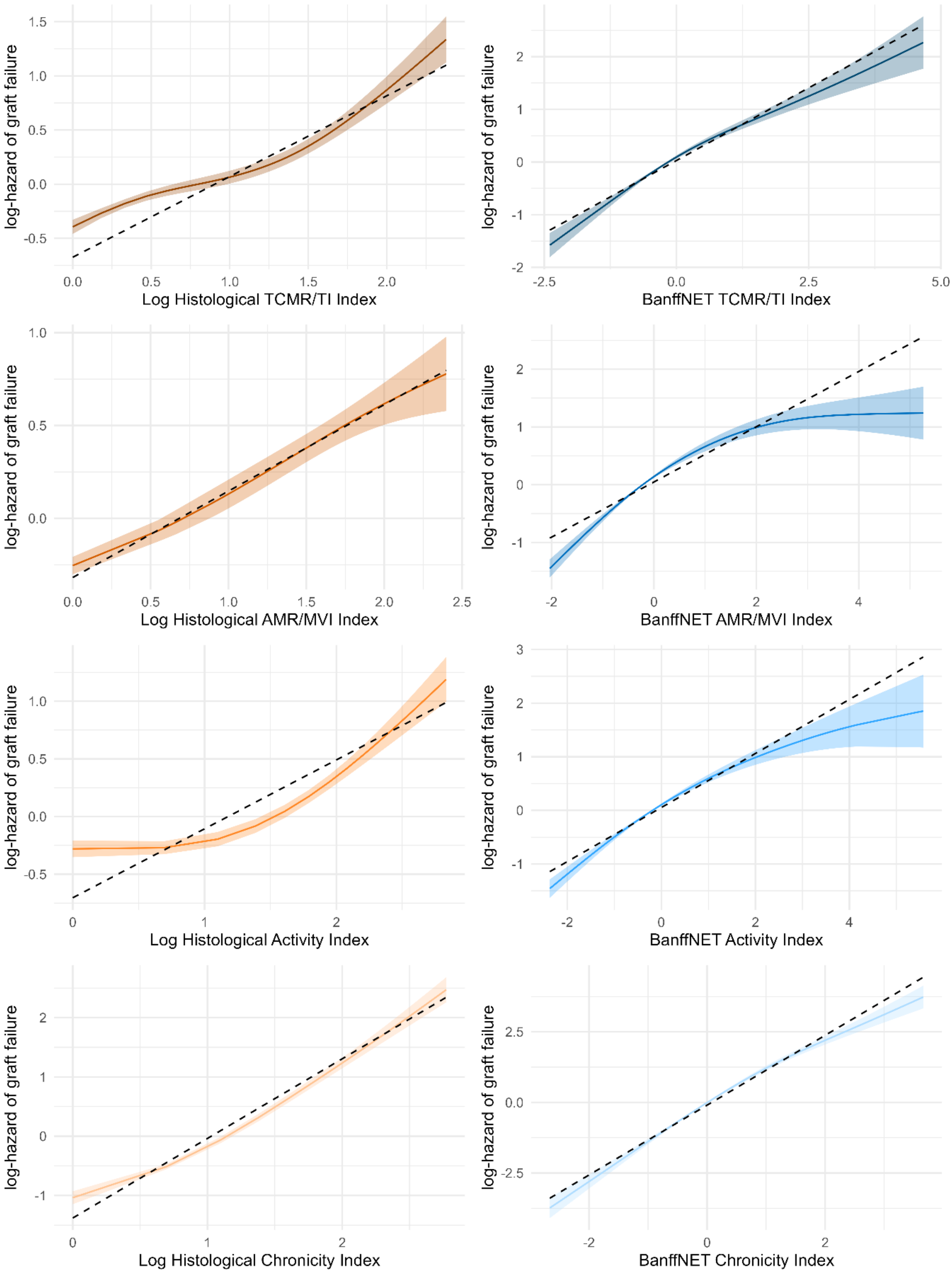

**Supplementary Figure 14. Linearity of associations with graft survival in the training cohort (n = 1874).** Effect plots with pointwise 95% confidence intervals to evaluate linearity in the associations of log-transformed histological indices and BanffNET Indices with the log-hazard of kidney graft failure.

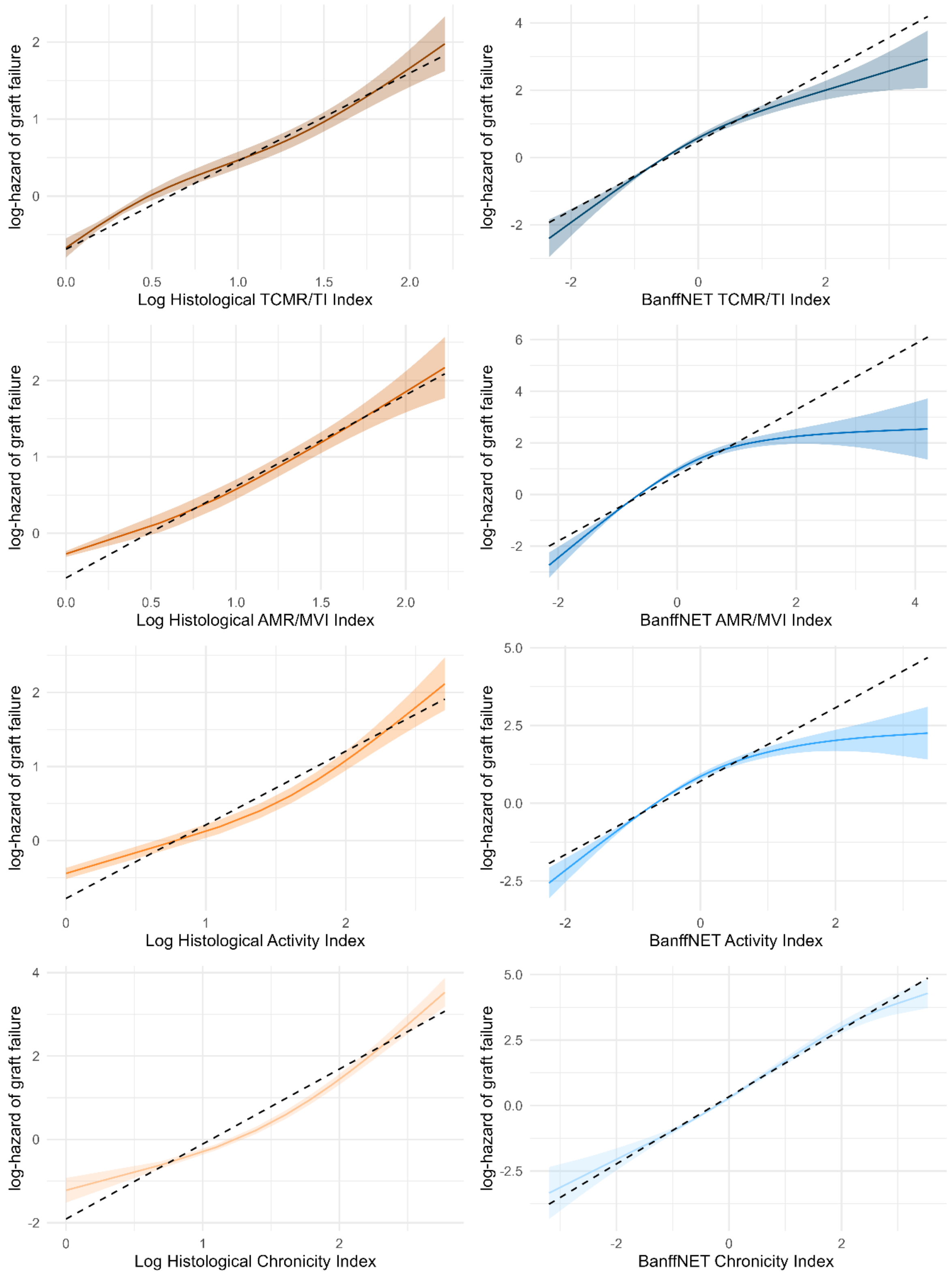

**Supplementary Figure 15. Linearity of associations with graft survival in the validation cohort (n = 3863).** Effect plots with pointwise 95% confidence intervals to evaluate linearity in the associations of log-transformed histological indices and BanffNET Indices with the log-hazard of kidney graft failure.

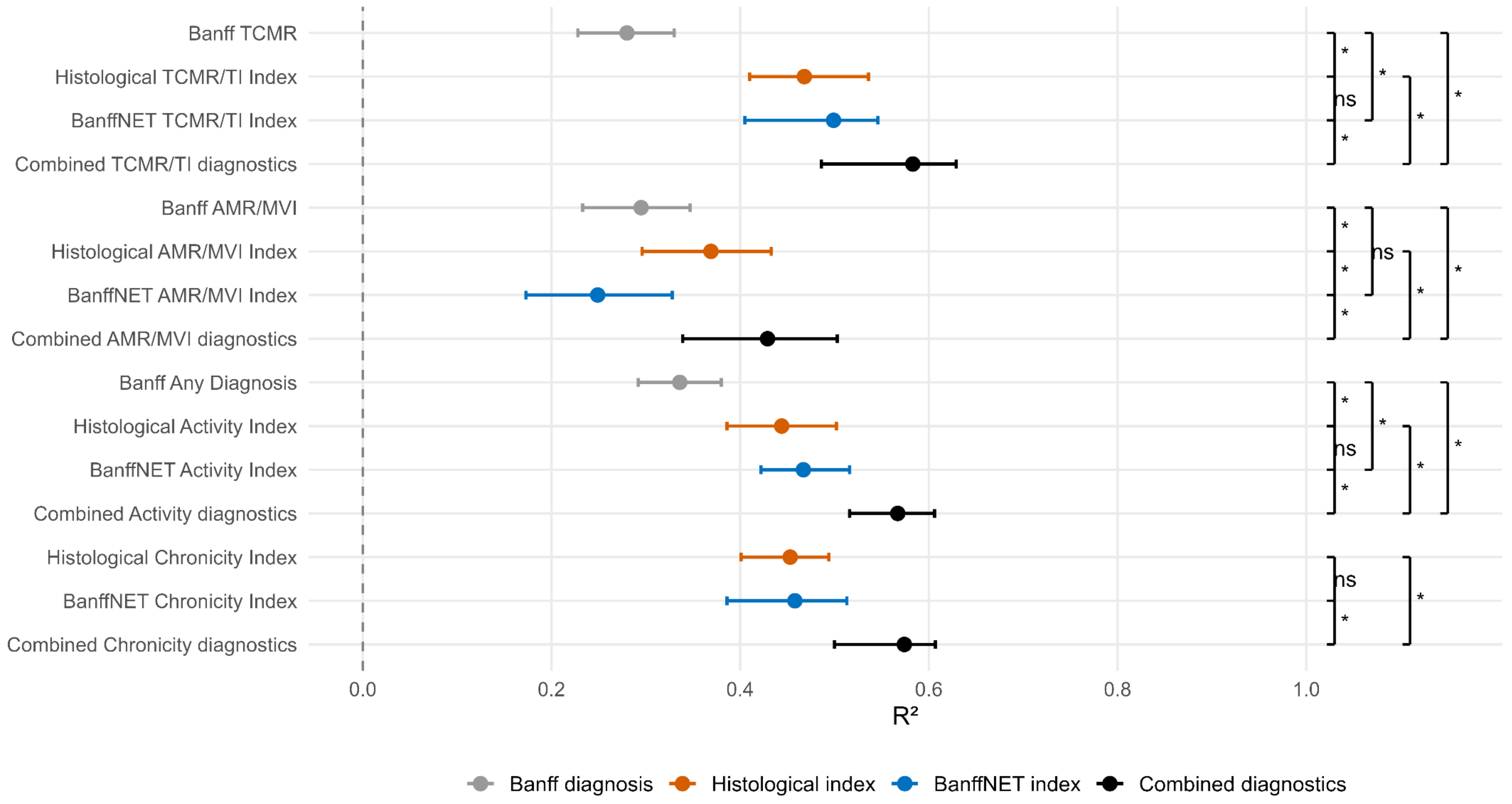

**Supplementary Figure 16. $R^2$ values for BHOT molecular Indices in the validation cohort (n = 3863).** 95% Confidence bounds were calculated for the $R^2$ values as well as for differences between $R^2$ values. Differences were considered significant if zero was not included in the confidence bounds. Significant differences were indicated with an asterisk, while non-significant differences were indicated with 'ns'.

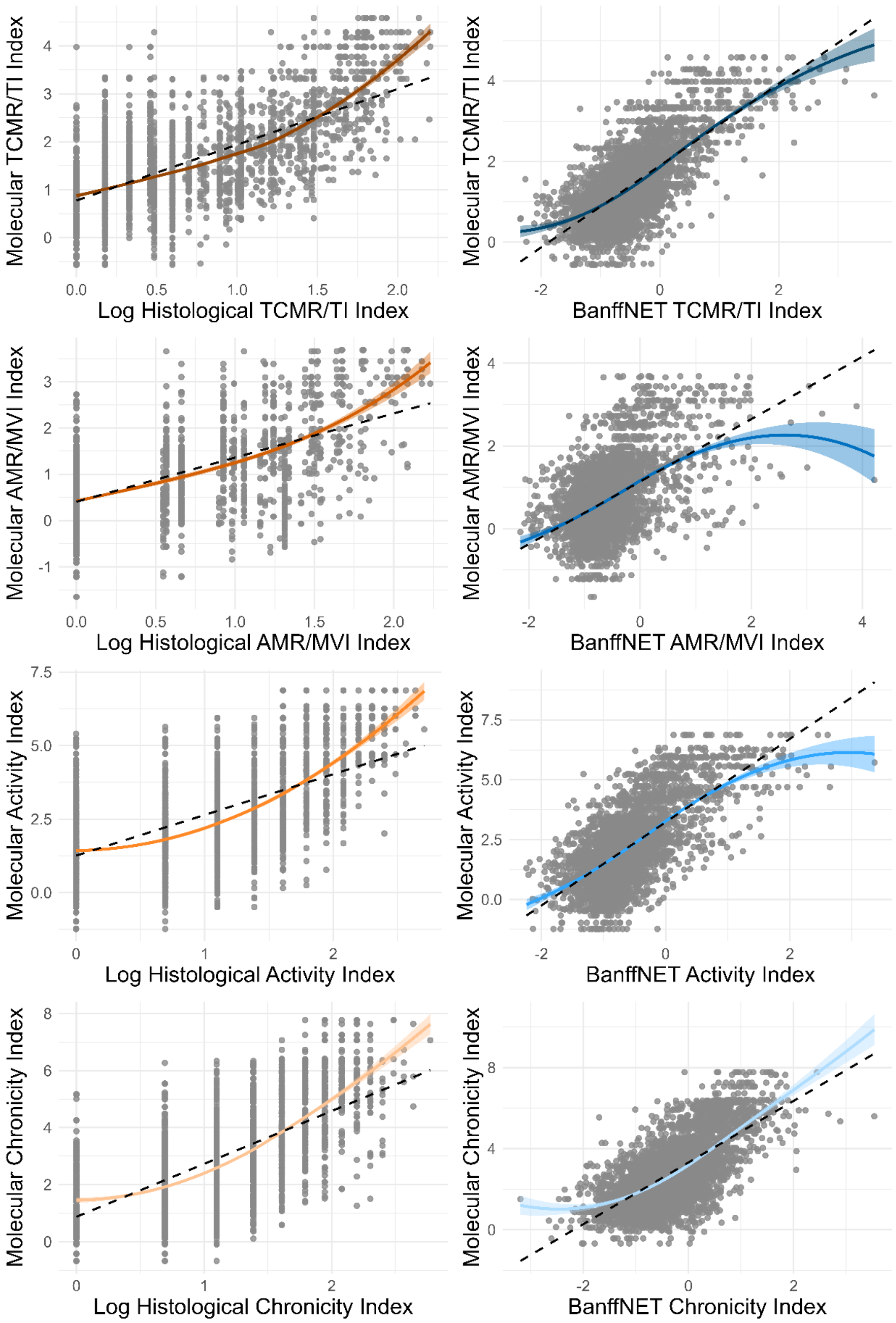

**Supplementary Figure 17. Linearity of associations with BHOT molecular Indices in the validation cohort (n = 3863).** Loess-smoothed scatter plots with pointwise 95% confidence intervals to evaluate linearity in the associations of log-transformed histological indices and BanffNET Indices with BHOT molecular Indices.
